# Discovery of Novel Digital Biomarkers for Type 2 Diabetic Nephropathy Classification via Integration of Urinary Proteomics and Pathology

**DOI:** 10.1101/2023.04.28.23289272

**Authors:** Nicholas Lucarelli, Donghwan Yun, Dohyun Han, Brandon Ginley, Kyung Chul Moon, Avi Z. Rosenberg, John E. Tomaszewski, Jarcy Zee, Kuang-Yu Jen, Seung Seok Han, Pinaki Sarder

**Author notes:** Correspondence: Seung Seok Han, Department of Internal Medicine, Seoul National University College of Medicine 103 Daehak-ro, Jongno-gu, Seoul, 03080, Republic of Korea; Pinaki Sarder, Department of Medicine-Quantitative Health, University of Florida College of Medicine, 1600 SW Archer Rd, Communicore CG-98, Gainesville, FL, USA. All authors have seen and approved the manuscript. indicates equal contributions (Nicholas Lucarelli, Donghwan Yun, Dohyun Han).

## Abstract

**Background:** The heterogeneous phenotype of diabetic nephropathy (DN) from type 2 diabetes complicates appropriate treatment approaches and outcome prediction. Kidney histology helps diagnose DN and predict its outcomes, and an artificial intelligence (AI)- based approach will maximize clinical utility of histopathological evaluation. Herein, we addressed whether AI-based integration of urine proteomics and image features improves DN classification and its outcome prediction, altogether augmenting and advancing pathology practice.

**Methods:** We studied whole slide images (WSIs) of periodic acid-Schiff-stained kidney biopsies from 56 DN patients with associated urinary proteomics data. We identified urinary proteins differentially expressed in patients who developed end-stage kidney disease (ESKD) within two years of biopsy. Extending our previously published human-AI-loop pipeline, six renal sub-compartments were computationally segmented from each WSI. Hand-engineered image features for glomeruli and tubules, and urinary protein measurements, were used as inputs to deep-learning frameworks to predict ESKD outcome. Differential expression was correlated with digital image features using the Spearman rank sum coefficient.

**Results:** A total of 45 urinary proteins were differentially detected in progressors, which was most predictive of ESKD (*AUC*=0.95), while tubular and glomerular features were less predictive (*AUC*=0.71 and *AUC*=0.63, respectively). Accordingly, a correlation map between canonical cell-type proteins, such as epidermal growth factor and secreted phosphoprotein 1, and AI-based image features was obtained, which supports previous pathobiological results. **Conclusions:** Computational method-based integration of urinary and image biomarkers may improve the pathophysiological understanding of DN progression as well as carry clinical implications in histopathological evaluation.

**Significance Statement:** The complex phenotype of diabetic nephropathy from type 2 diabetes complicates diagnosis and prognosis of patients. Kidney histology may help overcome this difficult situation, particularly if it further suggests molecular profiles. This study describes a method using panoptic segmentation and deep learning to interrogate both urinary proteomics and histomorphometric image features to predict whether patients progress to end-stage kidney disease since biopsy date. A subset of urinary proteomics had the most predictive power in identifying progressors, which could annotate significant tubular and glomerular features related to outcomes. This computational method, which aligns molecular profiles and histology, may improve our understanding of pathophysiological progression of diabetic nephropathy as well as carry clinical implications in histopathological evaluation.

## Introduction

Diabetic nephropathy (DN), in the setting of type 2 diabetes, is the leading driver of chronic kidney disease (CKD) and end-stage kidney disease (ESKD) worldwide.^1^ The incidence of DN is increasing due to high prevalence of diabetes, with subsequent kidney complications such as proteinuria and a decline in kidney function affecting more than 40% of patients.^2–4^ The result is an increased burden of cardiovascular morbidity and mortality,^5^ but the current therapeutic tools including lifestyle modification, blockers of renin-angiotensin-aldosterone system and SGLT2 inhibitors may not be sometimes effective to manage the rising incidence of DN and comorbidities. The heterogeneous phenotype of DN complicates a comprehensive approach, with a subset of patients having an abrupt decline in kidney function requiring dialysis within a few years of diabetes diagnosis.^6^

Kidney biopsy is the gold standard diagnosis of kidney disease, and histology aids in predicting prognosis and/or response to therapy.^7^ The utility of a renal biopsy in the setting of diabetes, beyond staging of DN, is critical to identifying non-diabetic processes (up to 30% of cases)^8, 9^ with a different etiology and disease trajectory compared with DN.^10, 11^ For improved classification and staging of DN, the Renal Pathology Society generated a DN classification schema.^12^ This system reflects the course of progressive DN well,^13, 14^ but has been hindered by the lack of practical application and implementation.^15^ Recent approaches have interrogated molecular profiles with classical grading systems of tissue histology such as T cell- and antibody-mediated rejection, glomerulonephritis, and glomerular nephropathy,^16–18^ but the resulting integrated metric has not yet been reported for the RPS DN classification schema.

Artificial intelligence (AI) has the potential to classify kidney disease histology in an automated fashion, which reduces the burden on pathologists and promises to improve reproducibility and robustness.^19^ Our previous work demonstrated digital RPS classification of diabetic glomerulopathy in renal biopsies^20, 21^ where our models achieved high levels of agreement with pathologist classification. To demonstrate the value added by these computational techniques, AI-based supervision for morphological changes related with molecular profiles or patient outcomes, but imperceptible to the human eye, are a promising next step.^22–24^ Herein, we integrated urinary proteomic profiles with AI-based histology images from biopsy-confirmed DN cases. The urinary proteomic model performed better than the AI-based image feature models in predicting early deterioration of kidney function. Several significant proteins and/or relevant pathways were associated with histological changes at the pixel and morphological levels. These results suggest that the urinary proteomic dataset could improve annotation of meaningful AI image features. Based on our results, AI will enable the extraction of molecular data encoded in the DN histology or in CKD histology in general. The resulting framework will stimulate the next revolution in histology-guided care of DN and generate new hypotheses for discovering novel pathobiology with diagnostic and prognostic implications.

## Results

### Renal Tissue Multicompartment Segmentation

A PAS-stained renal tissue biopsy whole slide image (WSI) from each subject was segmented into six classes: image background, interstitium (excluding perivascular stroma), non-sclerotic glomeruli, globally sclerotic glomeruli, tubules, and arteries/arterioles. We used features quantified from the glomerular and tubular image segments for our computational DN prediction study in this work. The segmentation was first performed by our trained panoptic segmentation network, and each slide was then manually corrected for any errors or missed segmentations.^25^ Approximately 30% of glomeruli and 10% of tubules required at least some manual correction. Examples of corrected network predictions are shown in ***Fig. 1*.** For this study, only cortical regions of the biopsies were used for analyses. Therefore, medullary regions were manually annotated and excluded from further analyses. Panoptic Quality (PQ),^25, 26^ was measured for segmentation of glomeruli, globally sclerotic glomeruli, and tubules in cortical regions. PQ ranges from 0, indicating no object recognition or completely incorrect object segmentation, to 1, indicating perfect segmentation of a given class when compared to ground truth. On average, *PQ*=0.76, 0.79, and 0.89 for segmenting glomeruli, globally sclerotic glomeruli, and tubules, respectively.

**Fig. 1.**
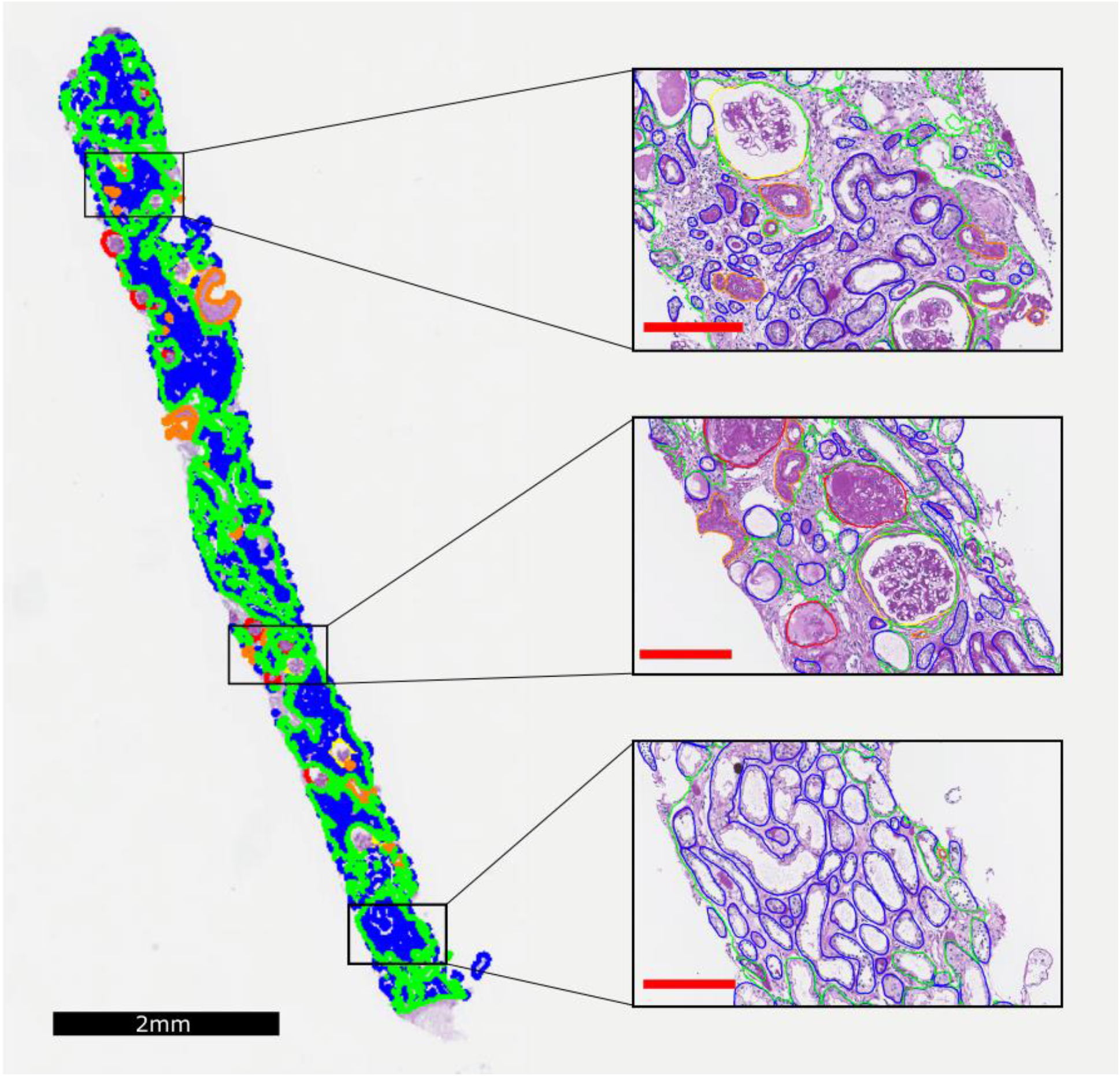
Whole slide image segmentation of renal Biopsy. Biopsy level predictions mapped back on a whole slide image (WSI) viewer Aperio ImageScope. Background (none), interstitium (green), glomeruli (yellow), globally sclerotic glomeruli (red), tubules (blue), arteries and arterioles (orange). Black scale bar = 2 mm. Red scale bars = 200 µm.

### Renal Tissue Sub-Segmentation

Glomerular sub-segmentation was carried out similarly to the method described by Ginley et al.^20^ in our earlier work and a similar methodology was employed for tubular sub-segmentation. Each glomerular and tubular instance was further segmented into three components: (1) nuclear, (2) PAS-positive, and (3) white space. Examples of the tubular sub-compartmentalization are shown in ***Fig. 2*.** This method of sub-segmentation has been shown previously to be useful in classifying patients into stages of DN, by using features extracted from the compartmentalized glomeruli.^20^ In this work, we studied the validity of such sub-segmentation in the context of prediction of DN progression. We did not analyze artery and arteriolar morphometries for our studies since, in a single WSI, the typical sparse appearance of fewer arteries/arterioles does not capture all possible 2D projections of these structures to determine feature information that would otherwise be invariant of feature variation as expected in 3D structures. Thus, feature quantification from arteries/arterioles for the subsequent analysis task would be prone to noise compared to glomeruli and tubules.

**Fig. 2.**
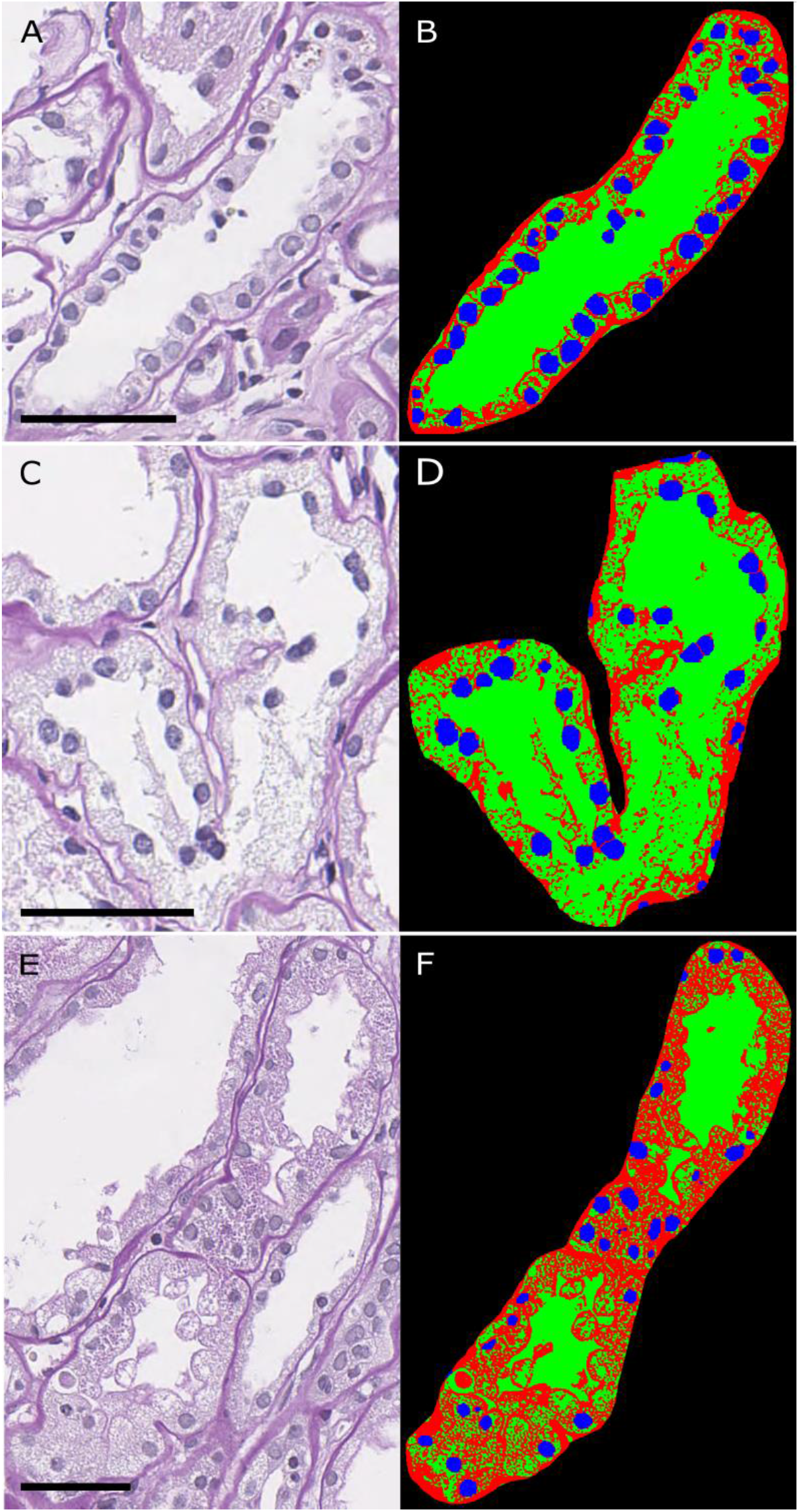
Component segmentation of renal tubules. Tubules segmented into nuclear component (blue), periodic-acid Shiff (PAS)+ component (red), and luminal/white space component (green). Scale bars = 25 µm.

### Renal Tissue Image Feature Quantification

Beofre feature quantification, crops of glomeruli and tubules were color-normalized using the Reinhard method,^27^ described above. Hand-engineered digital image features were measured from each segmented instance of glomeruli and tubules. The features measured per glomerulus have been described in our earlier work^20^ and new features were added, including component-specific distance metrics, such as quantiles of pixel distance from the glomerular border. In total, there were 315 digital image features measured for each glomerulus (***Supp. Table 3)***.

Tubular digital image features from the three components segmented of each tubule include pixel colors, textures, morphology, containment of one tubular sub-segment in another, and inter- and intra-structural distances. Examples of these feature extractions are shown in ***Fig. 3*.** One unique aspect of this set was the features added to measure more specific tubular morphology. While glomeruli typically appear circular in histological sections, tubules can have a highly variable shape. To account for this variance, we measured tubular curl,^28^ major axis length, perimeter, and area. These features quantify the tortuosity and size of each tubule. Also, features were defined to quantify tubular basement membrane (TBM) morphology. During feature extraction, the TBM was segmented from the rest of the PAS-positive area, and color, textural, and size features were measured for the resulting segmentation. Some examples of TBM thickness quantifications are shown in ***Fig. 3*.** TBM thickening is a feature of tubular atrophy and in particular has been shown to be of significant in diabetic kidney disease.^29^ A total of 207 digital image features were measured for each segmented tubule (***Supp. Table 4)***.

**Fig. 3.**
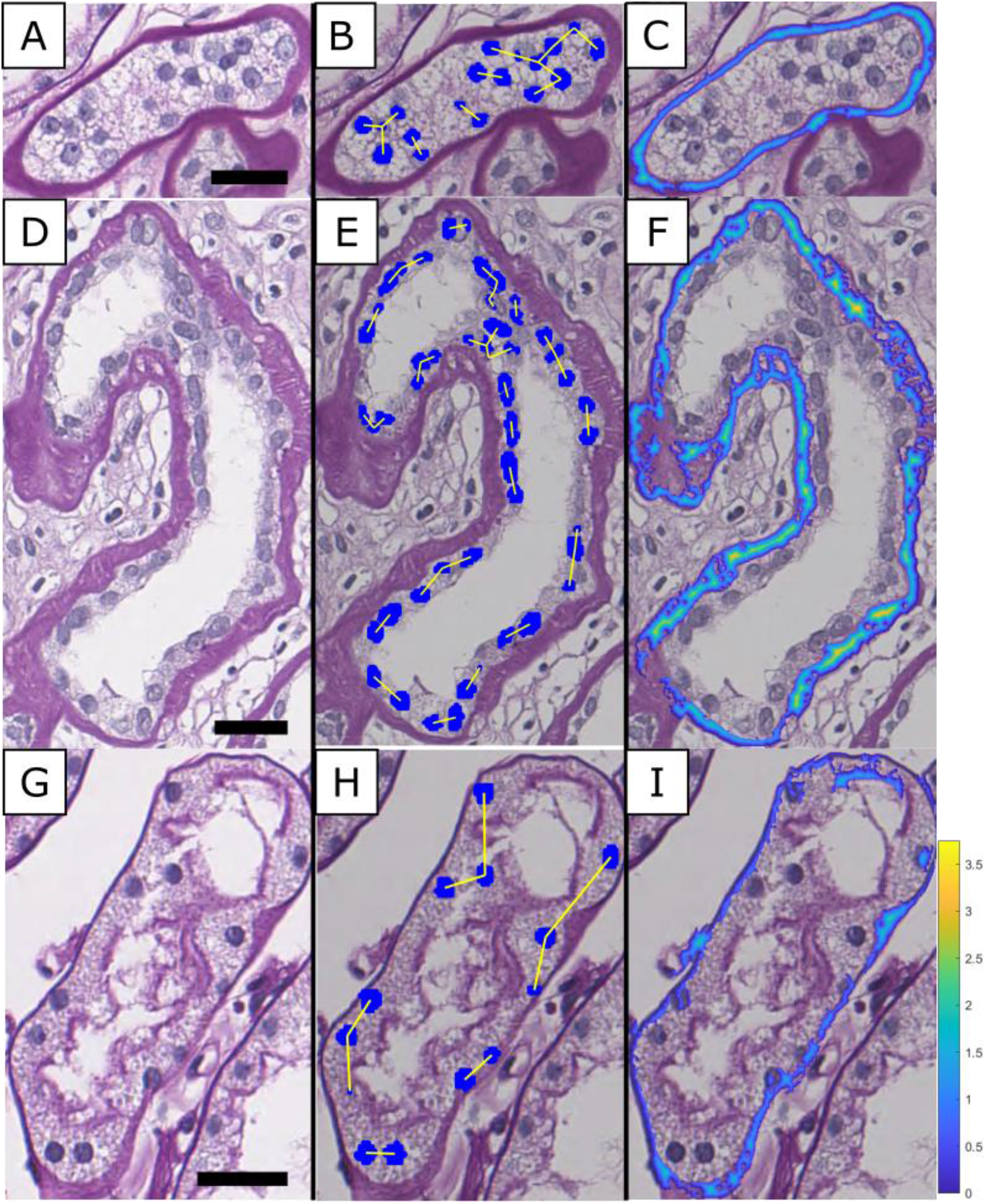
Feature extraction from renal tubules. (A, D, G) Original images from whole slide image (WSI). (B, E, H) Representation of closest internuclear distance. (C, F, I) Representation of tubular basement membrane thickness. Scale bars = 25 µm.

### Urinary Proteomics Data Feature Manifold Classification

The feature sets used in this study were initially analyzed using Seurat.^30, 31^ Each feature set (urinary proteins, glomerular image features, tubular image features) was visualized separately using Uniform Manifold Approximation and Projection (UMAP) to project and reduce the high-dimensional feature-space to a 2-dimensional plot.^32^ For subject-wise visualization of image feature sets in UMAP space, each feature was averaged, and the standard deviations were calculated across all object instances (i.e., for all the segmented glomeruli or tubules) in a single biopsy. An SVM was trained on each feature set to classify patients on whether they progressed to ESKD within 2 years of biopsy. To account for the imbalance in outcomes, the class weights were adjusted to be inversely proportional to class frequencies.^33^ For aggregated glomerular image features, the SVM achieved Matthews Correlation Coefficient (*MCC*), *MCC*=0.35. For tubular image features, we achieved *MCC*=0.58, and for the full urine protein set, *MCC*=0.60. MCC is used for measuring differences between predicted and actual values, ranging from -1 to 1, where -1 represents complete anti-correlation, 0 represents no correlation, and 1 represents perfect correlation, and is more robust when dealing with class imbalances. The simplified feature spaces and SVM hyperplanes are shown in ***Fig. 4***.

**Fig. 4.**
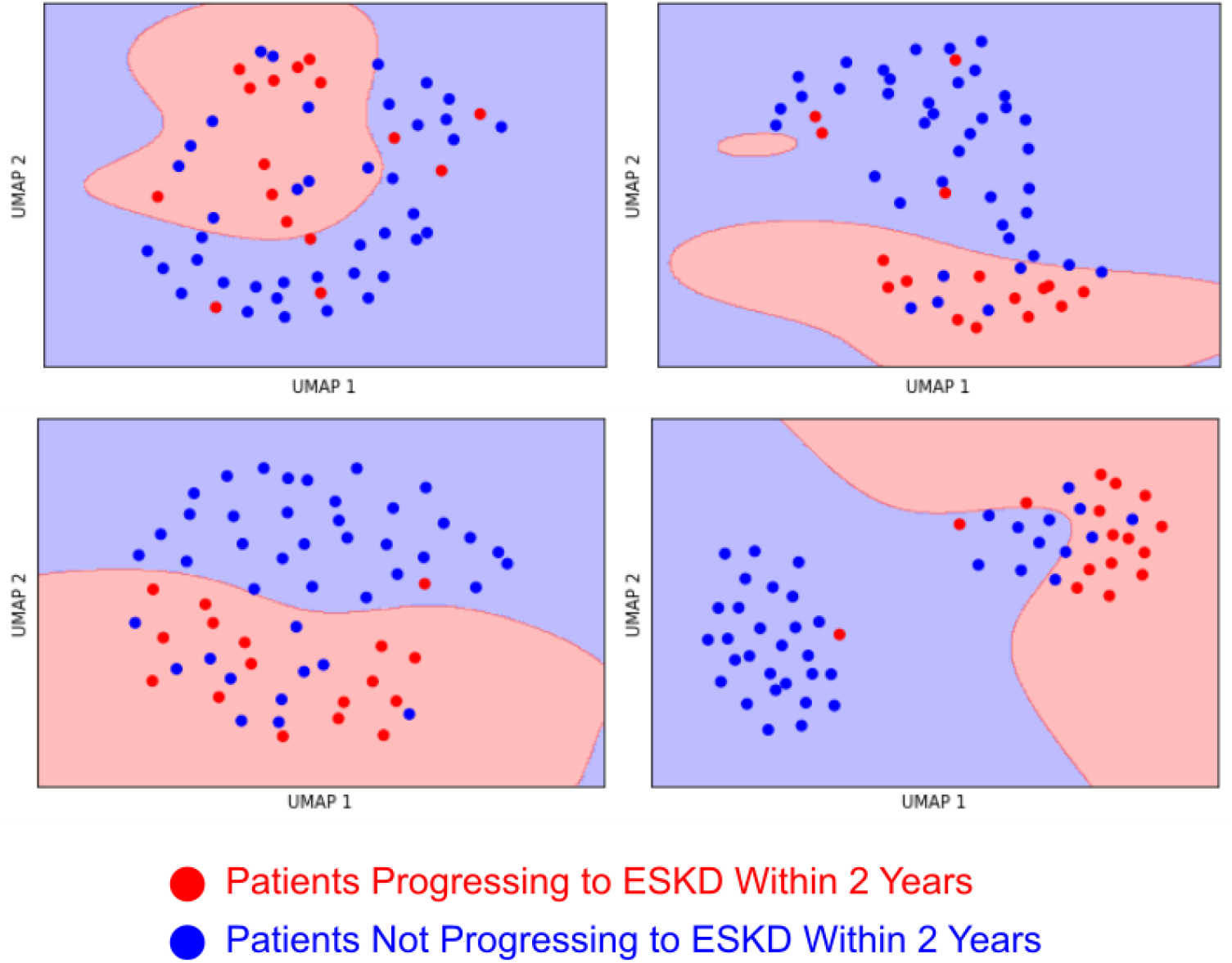
Uniform Manifold Approximation and Projection (UMAP) dimensionality reduction for various feature types. (A) Aggregated glomerular features, (B) Aggregated tubular features, (C) Full urine proteomics set, (D) Protein biomarkers related to end-stage kidney disease (ESKD) progression. Red dots represent patients progressing to ESKD within two years of biopsy/urine collection. Blue dots represent patients that did not. Red background represents support vector machine (SVM) prediction of progressing to ESKD within 2 years. Blue background represents SVM prediction of not progressing to ESKD within 2 years. Urinary proteins are more robust in delineating progressors from non- progressors than image features, and a reduced set of significant proteins increases predictive strength.

Using Seurat, we were also able to identify 45 proteins that were differentially measured (Bonferroni adjusted *p*<0.05) in subjects that progressed to ESKD. Cluster biomarkers were found using the *FindMarkers* function. Of these proteins, 25 were significantly higher in patients progressing to ESKD (***Table 1***), while 20 were significantly lower (***Table 2***). Using this limited set of differentially identified proteins, we reduced the number of features in the urine protein set and retrained the SVM, achieving an improved *MCC*=0.72.

### Mapping Urinary Proteomic Features to Pertinent Renal Cells using Single-Nuclei RNA Sequencing

The percentage of cells and average expression for each of the 45 differentially measured urinary proteins were assessed in single-nuclei RNA (snRNA) sequencing data for both non-diabetic control and diabetic kidney tissues (*n*=3 per each).^34^ Canonical cell-type candidate biomarkers, and protein expression for the differentially identified urinary proteins are shown in ***Fig. 5***. A total of 11 clusters including proximal convoluted tubule, parietal epithelial cell, thick ascending loop, distal convoluted tubule, collecting duct (principal cell, intercalated cell type A and B), podocyte, endothelial cell, mesangial cell, fibroblast, and immune cells were identified by cell marker genes. Further sub-clustering of the above mesangial cell and fibroblast clusters yielded additional three sub-clusters, namely, fibroblast, myofibroblast, and mesangial cell, see ***Fig. 5A-B***. The single cell gene expression of differentially measured urine proteins in DN subjects with progressive disease is summarized in ***Fig. 5C*.** This result provides insights linking likely cell-type of origin for differentially identified urinary proteins. Additionally, we quantified the changes in protein expression between non-diabetic and diabetic cases (***Fig. 5D***). As an example, there was a significant increase in the number of fibroblasts expressing the *C7* gene (gene name of Complement component 7) in diabetic kidneys, subclustered with mesangial cells and fibroblasts (***Fig. 5E***). We explored this relationship further in the context of our data (urinary proteomics and digital biopsy image features in DN), correlating it with differential image features. To summarize our findings thus far, we learned from our data that urinary proteomics data may be more informative than digital pixel-level image features of glomeruli and tubules, and certain urinary proteomics measurements may be originating from renal cells with prognostic significance for diabetic versus non-diabetic status.

**Fig. 5.**
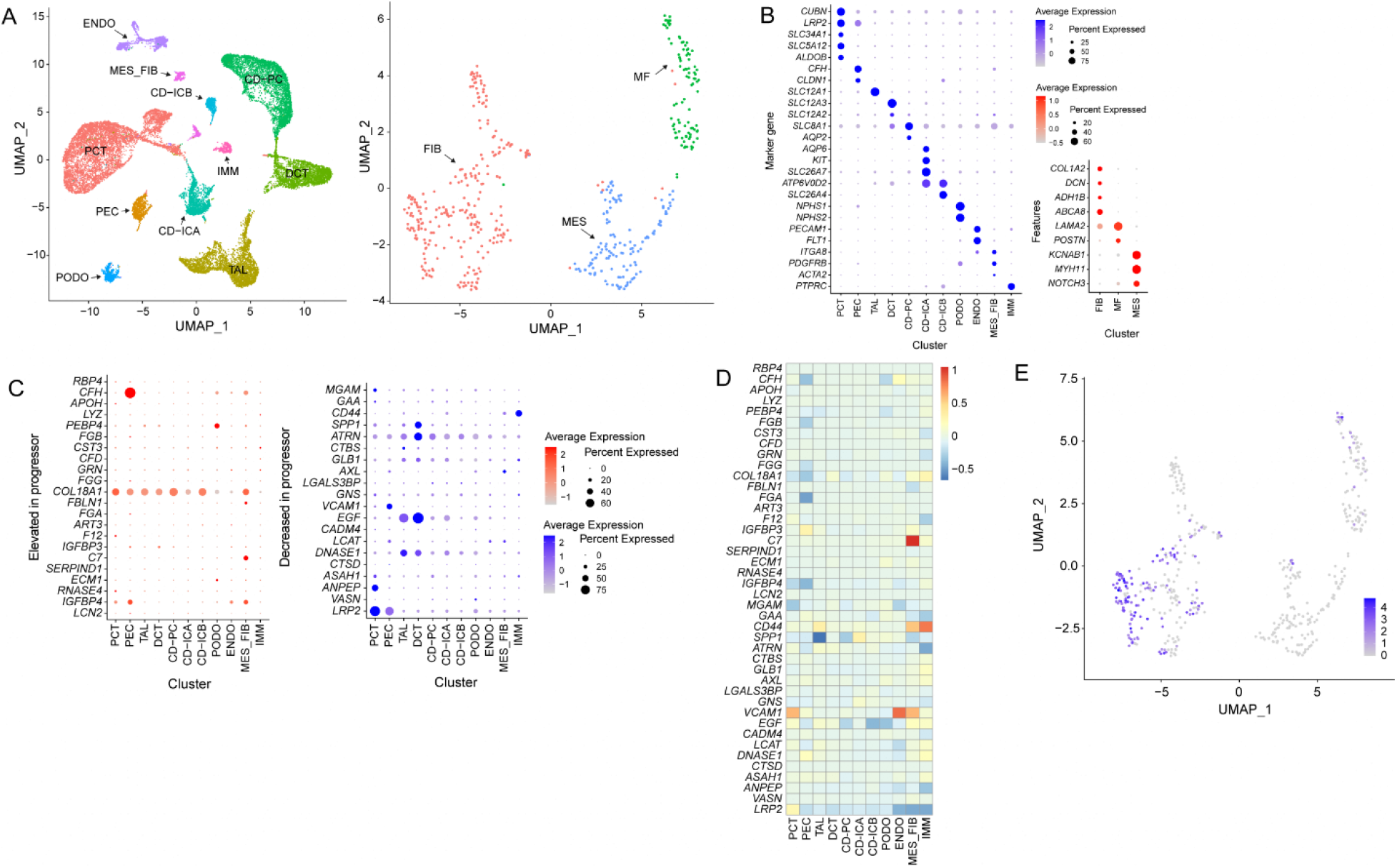
Mapping urinary proteomics data on an existing single cell RNA (scRNA) sequencing data to decipher cell locale of the identified urinary proteins. (A) UMAP (Uniform Manifold Approximation and Projection) plot of 23,987 cells pooled from a source with human kidneys (left) and a plot subclustered from a MES_FIB cluster (right). (B) Dot plots to identify clusters (left) and a MES_FIB cluster (right). (C) Dot plots for the gene expression of increased (left) and decreased (right) urine proteins related with progression of diabetic nephropathy. The genes corresponding to CFHR1, CFHR2, FGG, FGA, and GKN1 were not expressed in all the clusters. (D) Heatmap of logarithmic fold changes of genes in diabetic kidneys compared with non-diabetic kidneys. (E) Expression of the C7 gene in the UMAP plot of a MES_FIB cluster. PCT, proximal convoluted tubule; TAL, thick ascending limb of loop of Henle; DCT, distal convoluted tubule; CD-PC, principal cell of the collecting duct; CD-ICA, type A intercalated cell of the collecting duct; CD-ICB, type B intercalated cell of the collecting duct; PEC, parietal epithelial cell; PODO, podocyte; ENDO, endothelial cell; MES, mesangial cell; FIB, fibroblast; IMM, immune cell; MF, myofibroblast. Expression of genes encoding differentially identified proteins in snRNA sequencing data from diabetic and non-diabetic kidneys showed key differences in specific genes.

### Disease Progression Prediction using AI

AI was implemented to predict disease progression in patients with DN because of its unique ability to detect and model nonlinear relationships,^35^ which is important for studying nonlinear disease progression such as that of CKD.^36^ We used 2 neural network architectures to predict progression in DN patients. Urinary proteins are subject-level measurements, and therefore these features can be used directly to produce a classification through a series of dense layers, in a Fully Connected Neural Network (FCNN). However, the digital image features measured in this study are measured from multiple instances (namely, multiple segmented glomeruli or tubules) per patient, and must be aggregated to form a single prediction for a patient. We therefore used a Recurrent Neural Network (RNN) to incorporate all glomeruli or tubules to form predictions following recipes discussed in our previous publication.^20^ Since some of our data is unlabeled, we trained our networks using a self-training semi-supervised scheme.^37^ The results are aggregated from each subject when applied in the holdout set during 10-fold cross-validation. We compared the SoftMax predictions of each network to the patient’s ground truth label, and assessed the performance with the area under the curve (AUC) of the receiver operating characteristic curve. These results are shown in ***Fig. 6***.

**Fig. 6.**
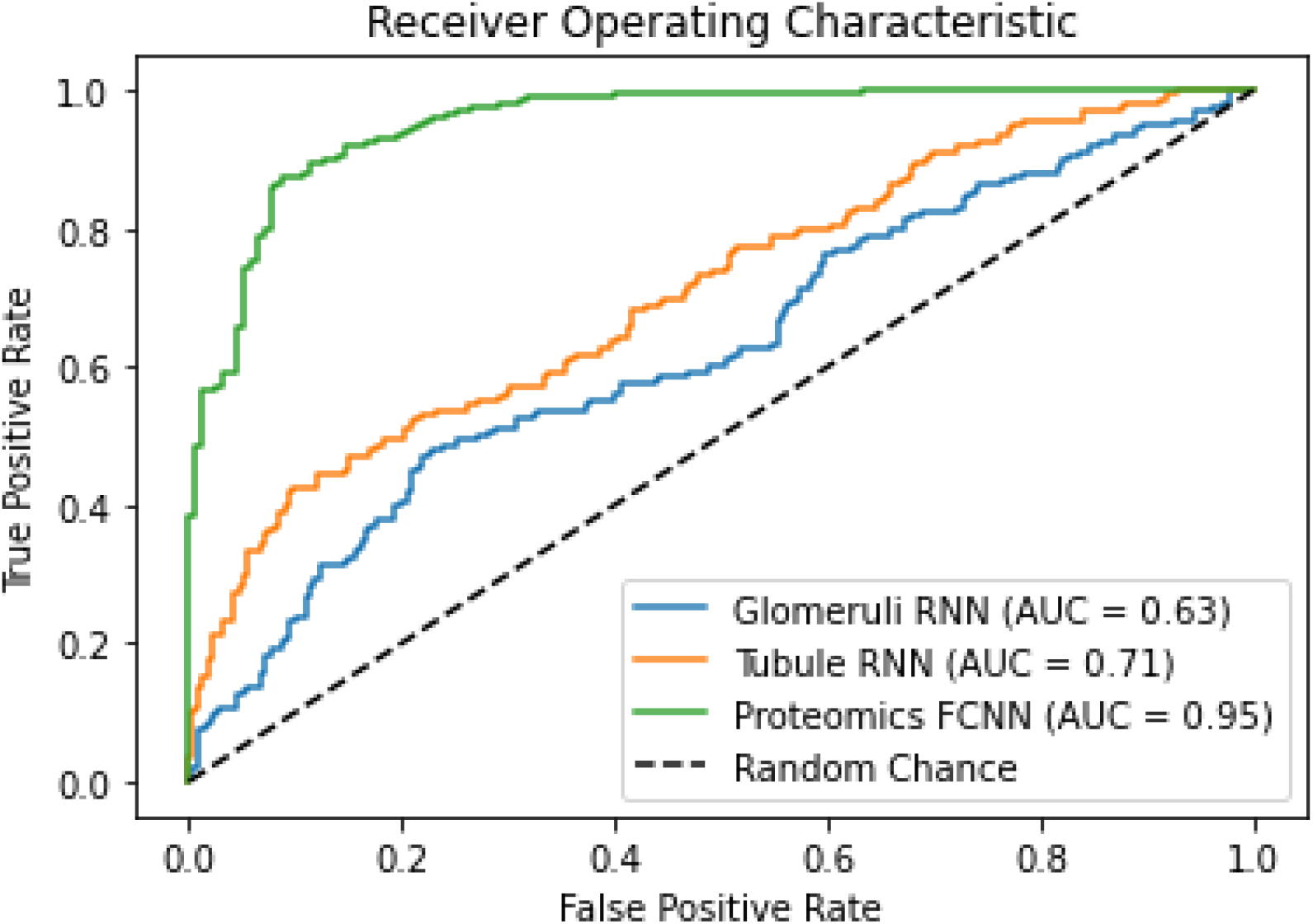
AI Network end stage kidney disease (ESKD) prediction performance. Results shown from fully connected neural network (FCNN) using ESKD marker proteins (green), recurrent neural network (RNN) using tubular digital image features (orange), and RNN using glomerular digital image features (blue). AI predictions compared to ground truth labels for *n* = 56. FCNN results shown for 100 trials of 10-fold cross validation. RNN results shown for 10 trials of 10-fold cross-validation. Urinary proteomics continue to show the greatest predictive power over image features.

We achieved the best performance when training on differentially identified urinary proteins in an FCNN, with *AUC*=0.95. This score indicates nearly perfect agreement between the classifier network and the ground truth label. Amongst image feature types, those measured for tubules outperformed glomeruli. Tubular image features achieved *AUC*=0.71, while glomerular image features achieved *AUC*=0.63. The difference in performance for tubular and glomerular image features may be a characteristic of the present cohort, or the difference in numbers of glomeruli compared to tubules in a typical biopsy sample that we analyzed. Nevertheless, the outperformance of urinary proteomic features may guide the significant annotation of image features in correlation to DN progression.

### Correlation of Renal Tissue Morphometry and Urinary Proteomics Data

Using Spearman’s rank correlation coefficient, we measured the degree of correlation between digital image features and urinary protein measurements, while controlling for confounding variables. We were interested in understanding how changes in the urinary proteome may be reflected in tissue sections and analyzing the significance of these relationships. Each digital image feature across all structures in each subject was matched with their respective differentially identified urinary proteomic profile, and with their associated molecular pathway scores. The measured correlation coefficients are displayed on heatmaps reflecting the degree of correlation between an image feature and a urinary protein or pathway (***Fig. 7***). On average, correlation coefficients with glomerular image features were greater than those with tubules, but *p*-values were lower for tubules, likely due to differences in sample size.

**Fig. 7.**
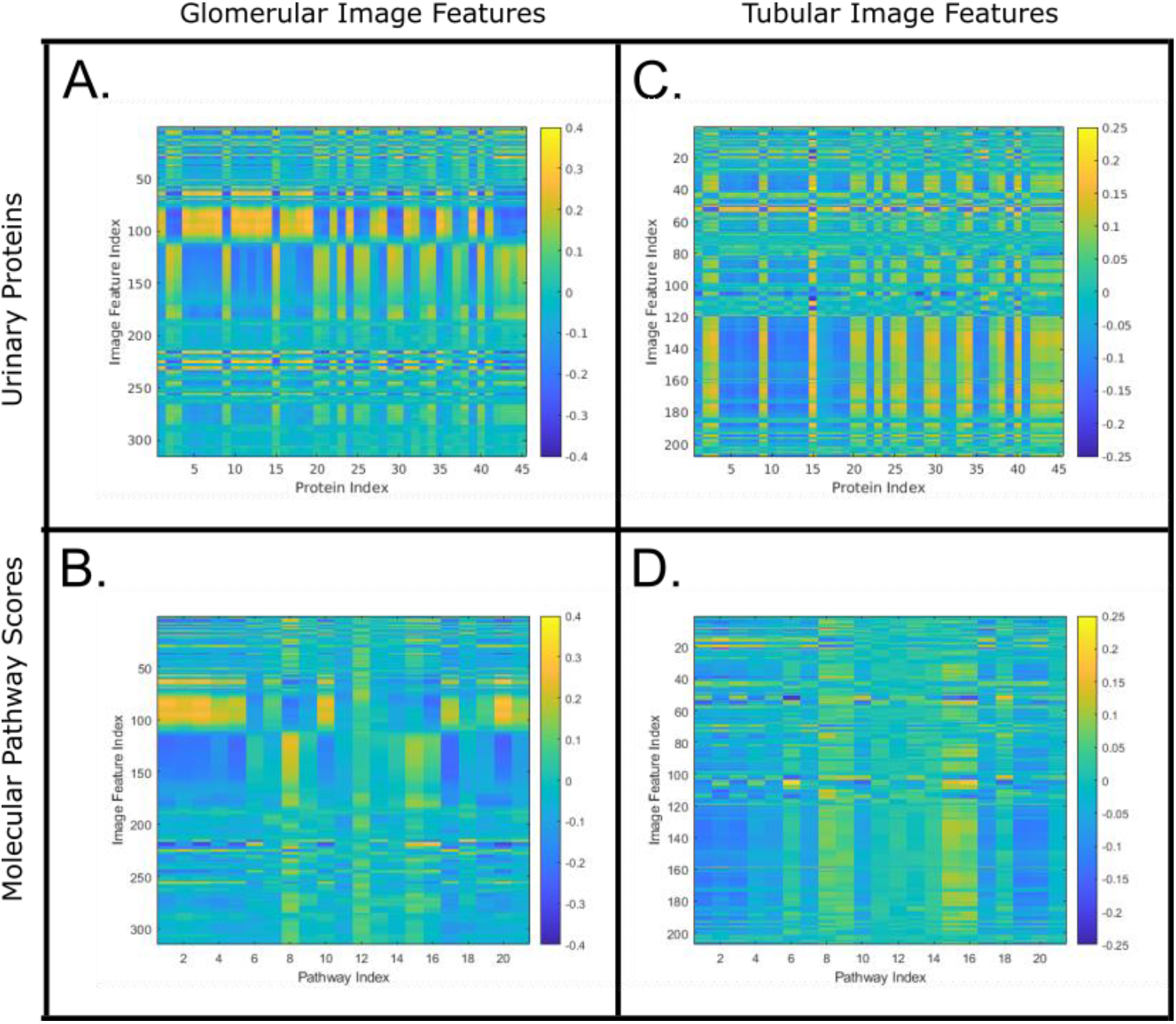
Spearman Rank Correlation Coefficients between digital image features and urinary protein measurements or molecular pathway scores. Color at intersection represents single spearman coefficient between pairs. Glomerular correlations were generally higher in magnitude, but tubular correlations were more statistically significant, with both of these relationships likely coming from differences in sample size.

### Discovery via Renal Tissue Image Pixel Parsing

As a select example among the highest Spearman rank correlation coefficients, when looking at the correlation coefficients calculated between non-sclerotic glomerular image features and urinary proteins, one of the most highly correlated pair consisted of the protein Complement component 7 (C7) and the mean value of red pixel values in PAS-positive region, with a coefficient value of 0.30 (FDR adjusted *p* <0.05). These pixel values are directly related to the perceived brightness in an image. However, of the three color channels, the green channel has the most significant effect.^38^ Furthermore, the standard deviation of the pixel values in an image is related to the image contrast,^39^ specifically within the PAS-positive region of the segmented glomerulus. Since this feature and protein have a direct correlation, as the amount of C7 present in the urine increases, one should expect to see an increase in this image feature. As demonstrated from snRNA sequencing data, C7 was expressed in renal fibroblasts. Furthermore, one can link this relationship to outcome, since C7 is also a urinary protein that is differentially elevated in patients developing ESKD. Some image feature examples are shown in ***Fig. 8***.

**Fig. 8.**
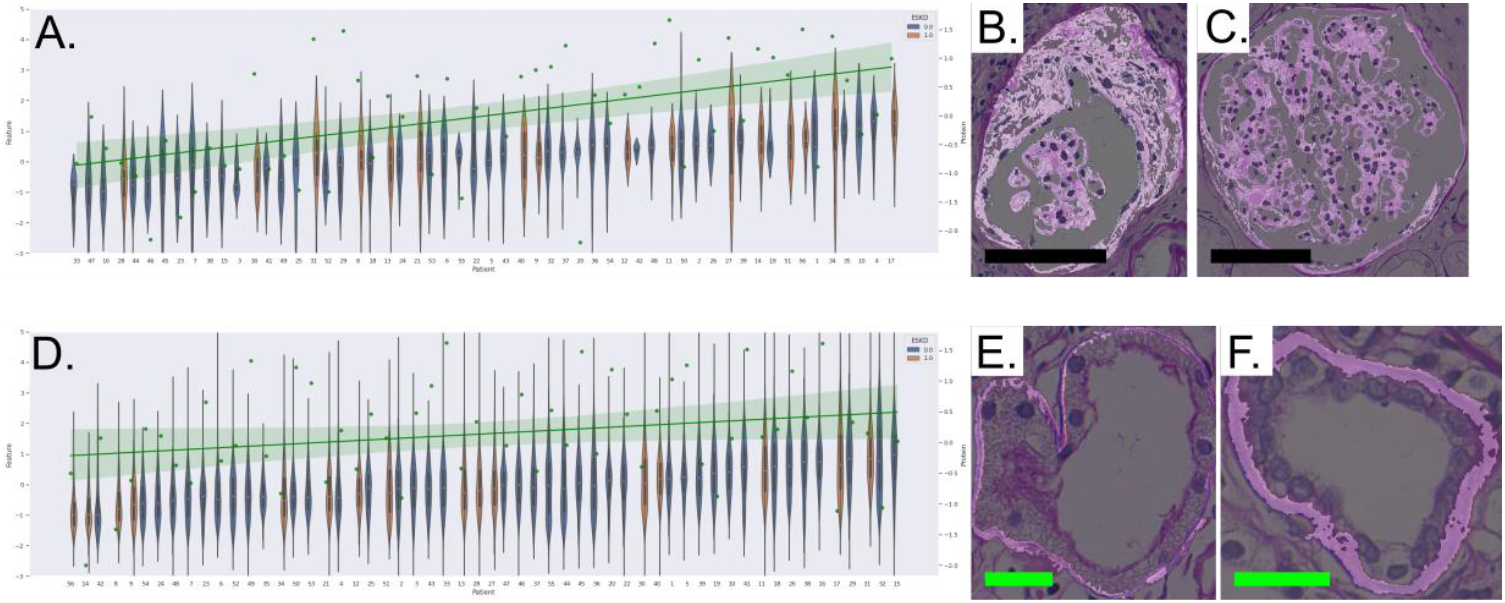
Pixel parsing discovery result for significant correlations between urinary proteomics and digital image features. (A) Violin plot of the average value of red pixel values in glomerular PAS+ regions, plotted for each patient in order of increasing mean feature value. Scatter plot of urinary proteomic measurements for Complement C7. Correlation between image feature and protein value is given by Spearman Rank Correlation Coefficient 0.30 (p<<0.0001). (B) Image of candidate glomerulus with low feature value, and (C) image of candidate glomerulus with high feature value. Black scale bars = 100 µm. (D) Violin plot of the standard deviation of green pixel values in the tubular basement membrane, plotted for each patient in order of increasing mean feature value. Scatter plot of urinary proteomic measurements for Epidermal Growth Factor. Correlation between image feature and protein value is given by Spearman Rank Correlation Coefficient 0.15 (p<<0.0001). (E) Image of candidate tubule with low feature value, and (F) image of candidate tubule with high feature value. First image highlights PAS+ segmented area, while second image shows z scores of green pixel values in PAS+ areas. Green scale bars = 25 µm.

When looking at correlation coefficients between tubular image features and urinary proteins, a few interesting cases arose. One of the most highly correlated pairs consisted of the epidermal growth factor (EGF), and the standard deviation of green pixel values in the TBM regions, with a coefficient value of 0.15 (*p* <0.05). Similar to the glomerular example, EGF was also relatively highly correlated with the standard deviations of the red and blue pixels in the TBM regions. As the amount of EGF present in the urine decreases, one should also expect to see a decrease in these features in the image data. EGF has previously been implicated in the development of acute kidney injury and CKD,^40^ and is expressed in renal tubular cells,^41^ as supported by our snRNA sequencing results.

Overall, the most highly correlated feature-protein pairs involved the secreted phosphoprotein 1 (SPP1). SPP1 is another protein that is found in renal tubular cells, as shown in both our snRNA sequencing results, and previous studies,^42^ while the receptor is found in immune cells.^43^ These urinary proteins were elucidated by this study for their relations to renal morphology in histological images, but their specific roles in kidney diseases are currently not fully understood.^44^

## Discussion

AI utilization has achieved remarkable advances particularly in segmenting microscopic structures of kidney. The next revolution may include the guidance of AI annotation to kidney histology while providing implicative information, such as patient outcome and relevant molecular profile, in addition to histology itself. As the first challenge, we correlated image features segmented by AI with urinary proteomic profiles in biopsy-proven DN with type 2 diabetes. Signatures in urinary proteins could discover novel histologic features related to DN progression; herein, various tubular image features, rather than glomerular, were selected to be associated with worse outcome. Furthermore, we applied various approaches, such as pathway analysis and pixel-based histologies, which may inspire researchers to consider how to expand the AI use in several kidney diseases as well as DN.

Biopsy is the current gold standard for diagnosis and outcome prediction of kidney disease, and predicting response to therapy. In this work, we examined the digital morphology of glomeruli and tubules as seen in renal biopsies. It is much more manageable for a pathologist to analyze individual glomeruli, since these structures are significantly less present in the biopsy as compared to tubules. The large number of tubules present in a biopsy makes it difficult for pathologists to integrate data over the entire WSI and make an objective prognosis. In this work, we are able to use the power of AI and computation to study image features from a large number of tubules, and found that the tubular image features from a standard needle biopsy are more predictive of disease progression than glomerular features. Therefore, we also suggest that computational methods like those we have demonstrated provide an advantage over current diagnosis standards, since our methods can provide a reproducible and comprehensive view of the tubular morphology.

We have also demonstrated the ability to link urinary proteomics data to quantitative image features. Qualitative image analysis has long been the standard for assessing pathology, but our results show that proteomics are robust in assessing progression in DN. Therefore, discovering and investigating links between image features and molecular profiles can aid in further research to increase our understanding of the nonlinear nature of DN. This methodology is also not limited to DN, or specific to renal pathology. Investigating these links further could lead to discoveries in the area of nondiabetic kidney disease. Integration of other data types (e.g., spatial tissue or blood omics) are also applicable in this case, depending on investigative goals and resources.

As discussed in Ginley et al.,^20^ sub-segmental distinction of both glomeruli and tubules in accordance to underlying pathobiology is complex. Our sub-segmentation of glomeruli and tubules does not fully reflect actual components of the glomerulus and tubule. The simplification of each structure into three basic components (nuclear, PAS-positive, and white space) based on their colorimetric appearance is a limitation to our performance in quantifying disease progression. Also, we largely analyzed each data source (image and molecular) individually, and there is a potential value in integrating image and molecular data to predict disease progression, which can be an area of future investigation. Lastly, our snRNA sequencing data come from non-diabetics and early diabetic, while the urinary proteomics and histology are sampled from various stages of DN. Therefore, the sequencing data may not be fully reflective of the conclusions we have made. To mitigate this shortcoming, our future study will focus on generating image, urinary proteomics, and snRNA sequencing data from same subject to tighten the conclusions made in this work.

The similarity in microscopic structure may not always guarantee the same pathophysiology, which is a cause of heterogenous outcomes within the same histological grade of kidney diseases. The current pathology reports made by human eyes would miss undistinguishable image features at the pixel level and not be linked to overall molecular signatures. The present study addresses this issue using urinary proteomics in the DN kidneys, and integrated results between urinary and histology signatures would provide better understanding of pathophysiology and the histological implication related to outcomes. The present study will be a basis of future studies using AI-based histological segmentation as well as multimodal molecular data to make a breakthrough in overcoming current clinical difficulties in understanding holistically a patient’s health state.

## Methods

### Human Samples

Study subjects were from Seoul National University Hospital, Seoul, Korea. Human data collection followed protocols approved by the Institutional Review Board at the Seoul National University Hospital (H-1812-159-998). All experiments were performed according to federal guidelines and regulations. Individual subject data contain patient demographic and medical history information, including height, weight, age, history of diabetes, stroke, and presence of hypertension. Blood tests were administered to the subjects at the time of biopsy. Serum creatinine was used to measure the estimated glomerular filtration rate (eGFR) at the time of biopsy using the CKD-EPI equation.^45^ Patients were re-evaluated and new blood samples were collected at one- and two-years following the initial biopsy and urine collection, and eGFR was recalculated. For each timepoint, it was determined whether serum creatinine had doubled compared with baseline, whether eGFR had reached 50% of the baseline value, and whether the subject had reached end stage kidney disease (ESKD). A full description of subject baseline characteristics can be found in ***Supp. Tables 1 and 2***.

### Whole Slide Image Data

Image data for this study consisted of brightfield microscopy WSIs of periodic acid-Schiff (PAS)-stained renal biopsies (2011-2017) from *n*=56 human DN subjects as a testing set. An additional *n*=30 WSIs from human DN patients from 2018-2021 were used as a validation set, but outcome data was not available for these patients. Whole slide images were collected on a digital slide scanner (Aperio AT2, Leica Biosystems, Wetzlar, Germany) with a 0.25 µm per pixel resolution. Following the Renal Pathology Society (RPS) DN scoring system,^12^ glomerular lesions, interstitial fibrosis and tubular atrophy (IFTA), interstitial inflammation, arteriolar hyalinosis, and arteriosclerosis were scored.

### Urine Sample Preparation

Urine sample of 1-2 ml from each subject was concentrated to 250 ul using a spin filter with a molecular weight cut-off of 3 kDa (Millipore, Billerica, MA). The protein concentration was measured according to the Bradford assay protocol (Bio-Rad protein assay kit, Bio-Rad, Hercules, CA). Protein of 50 µg was precipitated by adding a 5-fold volume of ice-cold acetone. The precipitated samples were reconstituted in 50 µl of SDT buffer (2% SDS, 0.1 M dithiothreitol in 0.1 M Tris HCl; pH, 8.0) and heated to 95°C. The denatured proteins were digested by a filter-aided sample preparation method as previously described^46^ with some modifications. Briefly, protein samples were loaded onto an Amicon 30K filter (Millipore, Billerica, MA), and buffer was exchanged with UA solution (8 M urea in 0.1M Tris-HCl; pH, 8.5) via centrifugation. After three buffer exchanges with UA solution, the reduced cysteines were alkylated with 0.05 M iodoacetamide in UA solution for 30 min at room temperature in the dark. Thereafter, UA buffer was exchanged for 40 mM ammonium bicarbonate twice. The protein samples were digested with trypsin/LysC (enzyme to substrate ratio of 1:100) at 37°C for 16 h. The resulting peptides were collected in new tubes via centrifugation, and an additional elution step was performed using 40 mM ammonium bicarbonate and 0.5 M sodium chloride. Peptides were desalted and fractionated on a homemade styrene divinylbenzene reversed-phase sulfonate-StageTips by basic reverse-phase using a stepwise gradient of acetonitrile (40%, 60%, and 80%) in 1% ammonium hydroxide. ^47, 48^ Fractionated peptides were completely dried with a vacuum dryer and stored at -80°C.

### Establishment of a Matching Peptide Library

To construct a matching peptide library for matching between runs, we made a pool of sub-aliquots of the same protein amount (5 ug) from urines. The pooled urine proteins (100 ug) were digested using the two-step filter-aided sample preparation as described previously.^48, 49^

Digested peptides were desalted using Oasis HLB solid-phase extraction. For constructing the matching peptide library, 100 µg of purified peptides were fractionated using an Agilent 1260 bioinert HPLC (Agilent, Santa Clara, CA) equipped with an analytical column (4.6 × 250 mm; 5-µm particle). High-pH reversed-phase peptide fractionation was performed at a flow rate of 0.8 ml/min over a 60-min gradient using solvent A (15 mm ammonium hydroxide in water) and solvent B (15 mM ammonium hydroxide in 90% acetonitrile). A total of 96 fractions was collected at 30-second intervals over a 48-minute gradient. The collected fractions were noncontiguously concatenated into 24 fractions. In detail, early-, middle-, and late-eluting peptides were combined by mixing every 24^th^ original fraction (e.g., combining fractions 1, 25, 49, and so on). The pooled fractions were dried in a vacuum centrifuge and stored at - 80°C until liquid chromatography-tandem mass spectrometry (LC-MS/MS).

### LC-MS/MS Analysis

LC-MS/MS analysis was performed using Quadrupole Orbitrap mass spectrometers, Q-exactive plus (Thermo Fisher Scientific, Waltham, MA) coupled to an Ultimate 3000 RSLC systems (Dionex, Sunnyvale, CA) with a nanoelectrospray source as previously described with some modifications.^46, 47^ Namely, peptide fractions of individual urine samples were separated on a two-column setup with a trap column (300 µm I.D. × 0.5 cm, C18 3 µm, 100 Å) and analytical column (50 µm I.D. × 50 cm, C18 1.9 µm, 100 Å). Before sample injection, the dried peptide samples were reconstituted in solvent A (2% acetonitrile and 0.1% formic acid). After the samples were loaded onto the nano LC, a 90-min gradient from 8% to 30% solvent B (100% acetonitrile and 0.1% formic acid) was applied to all samples. The spray voltage was 2.0 kV in positive ion mode and the temperature of the heated capillary was set to 320°C. Mass spectra were acquired in data-dependent mode using a top 15 method. The Orbitrap analyser scanned precursor ions with a mass range of 300–1650 m/z and resolution of 70,000 at m/z 200. Higher-energy collisional dissociation (HCD) scans were acquired at a resolution of 17,500 at m/z 200. HCD peptide fragments were acquired at a normalized collision energy of 28. The maximum ion injection times for the survey and MS/MS scans were 20 ms and 120 ms, respectively.

### Data Processing for Label-Free Quantification

Mass spectra were processed with MaxQuant (version 1.6.1.0).^50^ MS/MS spectra were searched against the Human Uniprot protein sequence database (December 2014, 88,657 entries) using the Andromeda search engine.^51^ Primary searches were performed using a six- ppm precursor ion tolerance for total protein level analysis. The MS/MS ion tolerance was set to 20 ppm. Cysteine carbamido-methylation was set as a fixed modification and *N*- Acetylation of proteins and oxidation of methionine were set as variable modifications. Enzyme specificity was set to full tryptic digestion. Peptides with a minimum length of 6 amino acids and up to 2 missed cleavages were considered. The required false discovery rate (FDR) was set to 1% at the peptide, protein, and modification levels. To maximize the number of quantification events across samples, matching between runs was performed using the pooled urine sample as a matching library. For label-free quantification, the Intensity Based Absolute Quantification (iBAQ) algorithm^52^ was used as a part of the MaxQuant software. Briefly, iBAQ values calculated by MaxQuant were raw intensities divided by the number of theoretical peptides. Thus, iBAQ values were proportional to the molar quantities of the proteins. The proteomic data have been deposited in the ProteomeXchange Consortium (http://proteomecentral.proteomexchange.org)^53^ via the PRIDE partner repository (accession no. PXD037505).

### Human AI Loop (HAIL) Pipeline

To conduct computational segmentation of renal micro-compartments, we used our previously published, publicly available HAIL pipeline.^54^ This pipeline allows users to train segmentation CNNs directly from annotations produced in the WSI viewer Aperio ImageScope®. HAIL allows for an iterative process of network training, where the network’s own predictions produced after each training iteration can be manually corrected and used to bootstrap further annotation data.

### Panoptic Segmentation

Panoptic segmentation^25^ was performed on PAS stained WSIs to classify pixels into six categories, and to resolve separate instances (e.g., individual tubules) of the same class. These six categories include image background, interstitium (excluding perivascular stroma), non-sclerotic glomeruli, globally sclerotic glomeruli, renal tubules, and arteries/arterioles. To train the model, 126 WSIs of kidney biopsy from native diabetic, lupus nephritis, and transplant patients were selected due to the presence of minimal cortex (<10%) in the slide, and fully annotated for all six categories. The network architecture was modelled closely after Google’s Panoptic-Deeplab model.^55^ There are three primary differences between our network model, and DeepLab-Panoptic. First, to reduce the memory overhead, the feature encoding backbone used is ResNet50.^56^ Second, the output stride of our encoder is only 8 compared with 16 as in the DeepLab-Panoptic model. Finally, for the output of our instance decoder branch, rather than predicting a Hough transform, our branch predicts the distance transform of each object. Then, instance segmentation is performed by computing a watershed transform^57^ on the predicted output distance transform map. Classification IDs of the predicted objects are settled with the majority voting rule used in Panoptic-DeepLab. The network was trained on cropped image patches of 560 × 560-pixel size and batch size 2 for approximately 3.8M steps. The trained network was then used for prediction where medullary regions were manually annotated and excluded from the analysis. Finally, glomeruli and tubules with artefact were manually removed from the final segmentations to eliminate erroneous quantification. Approximately 30% of glomeruli and 10% of tubules needed some manual correction. Segmentation accuracy was assessed using the Panoptic Quality (PQ) metric.^26^ This metric joins segmentation quality and recognition quality to assess both semantic and instance segmentation performance. PQ was only measured for segmentation of glomeruli, globally sclerotic glomeruli, and tubules, since these were the classes used in later analyses. We selected five random WSIs from patients that progressed to ESKD and 5 random WSIs from patients that did not, and these images were used for performance analysis. Our manually corrected annotations for these slides were further corrected by a pathologist, and used as a ground truth.

### Glomerular and Tubular Sub-Segmentation

To simplify sub-segmentation of the annotated regions, glomerular pixels are assigned to one of three components based on their appearance in the PAS-stained biopsies, according to Ginley *et. al*^20^: (1) nuclei, (2) PAS-positive areas, including the glomerular basement membrane, mesangium, and Bowman’s capsule, and (3) luminal areas, including capillary lumina and the Bowman’s space. For tubular segmentation, the three compartments were similar, where PAS-positive areas included tubular basement membrane and “intra-tubular objects” consisting of PAS-positive areas of the tubule not contained within the basement membrane, such as the proximal tubule brush border.

Nuclei were detected in the tissue by a custom-coded Panoptic segmentation network. This network was trained on previously annotated data described by Ginley *et. al*.^20^ In both glomerular and tubular images, nuclei were segmented by the same network. In glomeruli, PAS+ components were segmented by a several step process. First, the original RGB image was color-normalized using the Reinhard method,^58^ then was transformed to the hue-saturation-value color space,^59^ the saturation channel was isolated, and the dark regions were brightened using a gamma transform.^60^ For each glomerulus, this gamma was set at a constant of 0.7. Otsu’s thresholding method^61^ was then applied to segment the PAS+ area, excluding the already segmented nuclei. Once the nuclear and PAS+ components were segmented, the remaining pixels not contained in these components were classified as luminal pixels. PAS-positive components of the tubules were segmented in a slightly different fashion. First, the RGB image was color-normalized, and then transformed to the LAB color space.^62^ Second, the lightness color channel (L) was isolated and a constant threshold of 80 was applied to the 8-bit pixels. All pixels, not already classified as nuclei, above this threshold were classified as luminal, or white-space components. All other pixels not contained in the nuclear or luminal segmentation were classified as PAS-positive components.

### Quality Control and Color Normalization

To detect batch effects in the histology samples, we used the open-source softwares HistoQC and Cohort Finder^63, 64^ for quality control. The “first” configuration was used on the *n*=56 testing samples with determined outcome status, and 2 batches were detected by the software. Each batch contained exactly 28 samples, with exactly 8 patients progressing to ESKD partitioned into each cohort. To control for stain variation between the 2 batches, the Reinhard method of color normalization^58^ was used. Before normalization, tissue masks were generated for the thumbnail images using a saturation threshold, RGB values for the tissue sections were converted to the LAB color space, and the averages and standard deviations for L, A, and B values were determined across the entire dataset (*n*=56). These values were then used to normalize image crops before feature extraction.

### Feature Engineering

For glomerular images, digital image feature types are defined according to the methods described by Ginley *et al*.^20^ These feature types included color, texture, morphology, containment of one compartment within another, interstructural distances, and intrastructural distances. A total 315 image features were measured for each glomerulus. This feature set was expanded from the original paper for this cohort. For tubular images, similar types of digital image features (color, texture, morphology, containment, intra- and interstructural distances) are quantified for the same respective structures. Additional features are defined for the tubular basement membrane, and to measure tubular tortuosity. A total 207 image features per tubule were quantified. A full list of digital image feature sets for glomeruli and tubules can be found in ***Supp. Tables 3 and 4***.

### Image/ Proteomic Feature Manifold Classification

Image/proteomic feature expression manifolds were studied in low dimensional space using a state-of-the-art software Seurat.^30, 31^ Seurat is typically used for studying single-cell RNA sequencing data, but can be extended to other high-dimensional data, such as digital image features and urinary proteomics without loss of generality. For image feature analysis, each datapoint is modelled as a computationally segmented individual microcompartment (e.g., tubule, glomerulus), and corresponding feature values are quantified engineered feature data. For urinary proteomics analysis, each datapoint is modelled as a single subject, and corresponding feature values are measured urinary proteomics feature expressions. As a first step, the data are normalized by a global-scaling normalization method. This method normalizes each feature by min-max scaling values between 0-1, multiplying by a scale factor, and then log-transforming the result. The data is then scaled to zero mean and unit variance. Principal component analysis was then performed on the scaled data. Non-linear dimensionality reduction using UMAP^32^ is performed using the first 20 principal components per datapoint. Class labels (e.g., subjects with ESKD vs. non-ESKD) of interest are applied to these points following dimensionality reduction.

We analyzed differentially measured proteomics and image features between various data classes to characterize these classes. Features that are differentially expressed between classes are discovered using the *FindMarkers* function in Seurat, which uses the Wilcoxon rank sum test^65^ to determine statistical significance. This function also automatically performs *p*-value adjustment using Bonferroni correction, based on the number of features in the dataset. Features with an adjusted *p* <0.05 were identified as differentially expressed.

To classify subjects based on ESKD binary outcome measure, we performed binary classification of the UMAP feature data using an SVM classifier.^66^ One-dimensional hyperplanes were optimized using a radial basis function kernel, with balanced class weight, to classify the subjects. The performance of the classifier was quantified using Matthews correlation coefficient.^67^

### Mapping Urinary Proteins to Source Renal Parenchymal Cells using Single-Nuclei RNA Sequencing Data

While fusing urinary proteomics data and digital renal tissue image features is an important goal of our study, it is also important to map the urinary proteomics data to pertinent renal cell types to investigate biological relevance of our findings. For this mapping, pooled snRNA sequencing data of early diabetic (*n*=3) and nondiabetic (*n*=3) patients from a previous study^34^ were used. Genes expressed in ≥3 cells, cells with ≥500 unique molecular identifier counts were used. The R^68^ and Seurat^30, 31^ software packages as well as *pheatmap* package (version 1.0.12; an R package to draw heatmap) embedded in R were used for data analyses and visualization. For mapping the following functionalities from these software packages were used: *SCTransform* for merging data with adjusting batch-effects; *FindNeighbors* and *FindClusters* for finding clusters; *RunUMAP, FeaturePlot, DotPlot,* and *pheatmap* for data visualization; and *Findmarkers* for identifying differentially expressed genes.

### Disease Progression Prediction using Urinary Proteins by Semi-Supervised Fully Connected Neural Network

For each patient in the test cohort of *n*=56 subjects, 2,038 unique urinary proteins were identified across all subjects at the time of the biopsy, and for the additional validation cohort (*n=*30), 2,313 urinary proteins were identified across all the subjects (***Supp. Table 5***). Proteins identified as contaminant were removed. To predict patient progression to ESKD, we designed a fully connected neural network (FCNN). For all the AI networks, a binary outcome for ESKD progression within 2 years of biopsy was used as ground truth using differentially detected proteins as input. The network architecture consists of an input layer for protein data, then a series of 2 dense layers, each with 30 hidden nodes. These dense layers both contain leaky rectified linear unit (ReLU) activation functions, and 50% dropout during training. The last dense layer connects to a two-node prediction layer, with a SoftMax activation function. Cross-entropy was used to compute the loss for this network. The network was trained with an Adam optimizer, with the learning rate set at 0.001.The network was trained in a self-supervised fashion^37^ with *n*=56 labelled cases and *n*=30 unlabelled cases. First, the labelled dataset was split into 90% training and 10% testing, and the network was trained using just this training set for 500 iterations with batch size 50. Then, predictions were produced for the additional dataset of *n*=30 that did not have any outcome label. The SoftMax output was clipped at a threshold over 0.90, so any network predictions with more than 90% confidence were kept, and assigned the label produced by the network. Next, the newly labelled cases were added to the original set, and a single training step of batch 50 was run. Then, predictions are reproduced for the second dataset of *n*=30, and the process is repeated for a total of 100 iterations. Lastly, predictions are run on the 10% holdout labelled set, and compiled into the final predictions. The network is then reset, and a different 10% holdout is taken for testing. This was repeated for 10 folds and the results reported are compiled from 100 trials of 10-fold cross validation. Performance was calculated using the area under the receiver operator characteristic curve. The generalized network architecture is shown in ***Supp. Doc. 1*.**

### Disease Progression Prediction using Image features by Semi-Supervised Recurrent Neural Network

To predict DN progression in our cohort using quantified image features from glomerular or tubular compartments, we employed the recurrent neural network (RNN) architecture designed by Ginley *et al*.^20^ The goal is to predict an outcome label for a given WSI by incorporating at the network input the ensemble of quantified image features per compartment basis serially with respect to all the glomeruli or tubules in a biopsy. The main modification of the network architecture with that described in Ginley et al.^20^ is the final prediction layer, where we use 2 prediction nodes for our binary outcome (ESKD vs. no- ESKD in 2 years after biopsy), with a SoftMax activation. All training parameters, including learning rate, batch size, and training steps were kept the same. Semi-supervised training was coded similarly to the FCNN above. First, 500 training steps of batch size 256 and 10-time steps were completed with 90% of the labelled dataset. Predictions were run on the unlabelled patients, and those with prediction confidence >75% were kept, assigned the predicted label, and added back to the training set. Then, 100 additional training steps are run with the expanded training set, and predictions on all the unlabelled data are rerun. This process is repeated 5 times for 500 additional training steps. Final predictions are compiled from 10 trials of 10-fold cross-validation. The generalized network architecture is shown in ***Supp. Doc. 1*.**

### Correlation of Renal Tissue Morphometry and Molecular Data

Our previous experiments investigate how urine proteins and digital image features of renal structures relate to DN outcome. We next sought to understand how these 2 orthogonal data modes are related. These relationships were quantified using partial Spearman’s rank correlation coefficient, which controlled for several covariates, including sex, age, height, weight, hypertension, number of years with diagnosed diabetes, and history of stroke and ischemic heart disease. For this analysis, we used *partialcorr* command embedded in MATLAB (Mathworks, Natick, MA).^69^ Coefficients were measured for one-to-one relationships between quantified structural image features and protein expression or scores of molecular pathways formed by an ensemble of proteins. We adjusted the *p*-values of the correlation measures using the Benjamini-Hochberg method.^70^

For the above correlative study, to quantify the functionality of the measured urinary proteins at pathway level, we developed a scoring method for upregulation and downregulation of molecular pathways, using Ingenuity Pathway Analysis (IPA; Qiagen, Hilden, Germany).^71^ First, the proteins included in each pathway were identified. Then, the raw protein measurements were added by one, and the resulting values were log-transformed. The resulting data were then min-max scaled between 0-1 per protein basis. Following scaling, values were adjusted by their biological expectation of upregulation/downregulation in pertinent pathways, according to the analytical algorithms embedded in IPA. Namely, the scores of the proteins that were expected to be upregulated during the upregulation of a particular pathway were kept the same. Scores of the proteins that were expected to be downregulated during the upregulation of a particular pathway were revised by subtracting their pertinent values from 1. This method was followed so that all protein values would increase/decrease similarly during upregulation/downregulation of a pathway. Lastly, the scaled values of the proteins for each pathway were summed for each patient, to obtain one score per pathway. The pathways evaluated are listed in ***Supp. Table 6***.

### Discovery via Renal Tissue Image Pixel Parsing

Of the most highly correlated pairs (structural image features vs protein expression or pathway scores), we focused our studies on proteins or molecular pathways with biological relevance to renal pathology. This focus limited the proteins of interest to those expressed in renal cell types, and molecular pathways of interest to those involving aspects of renal function. Distributions across patients were investigated for structural image features highly correlated with these biologically relevant molecular candidates. Discovered image features were visualized in MATLAB, via mapping the quantified image features in the image space. Projected maps were qualitatively studied by renal pathology experts, and relationships with compelling trends were used for future hypothesis generation.

## Data Availability

Data & Code Availability
Codes for Panoptic Segmentation are available at https://github.com/SarderLab/Watershed_Panoptic_Segmentation
Codes for Object and Feature Extraction are available at https://github.com/SarderLab/Object_and_Feature_Extraction
Codes for Neural Network prediction of DN progression are available at https://github.com/SarderLab/DN_progression_prediction
WSIs, corresponding multi-compartmental segmentations, trained model files and all the corresponding outcomes, as well as confounding data for analyzing the performance our finding are available at https://bit.ly/3MhFYQc. Raw measured proteomics data is available via Supp. Table 7.

https://github.com/SarderLab/Watershed_Panoptic_Segmentation

https://github.com/SarderLab/DN_progression_prediction

https://bit.ly/3MhFYQc

## Data & Code Availability

Codes for Panoptic Segmentation are available at https://github.com/SarderLab/Watershed_Panoptic_Segmentation

Codes for Object and Feature Extraction are available at https://github.com/SarderLab/Object_and_Feature_Extraction

Codes for Neural Network prediction of DN progression are available at https://github.com/SarderLab/DN_progression_prediction

WSIs, corresponding multi-compartmental segmentations, trained model files and all the corresponding outcomes, as well as confounding data for analyzing the performance our finding are available at https://bit.ly/3MhFYQc. Raw measured proteomics data is available via ***Supp. Table 7*.**

## Author Contributions

NL corrected computational segmentations, conceptualized and performed the quantitative analyses, designed, and conducted the computational methods, completed all statistical analyses, interpreted the results, and wrote the manuscript. DY analyzed bioinformatics data, interpreted the results. DH generated proteomics data. BG guided NL in conducting the work, reviewed the codes, as well as critically analyzed results. KCM collected and supervised histology data. AR and JET critically analyzed the results of the correlation study to decipher the biological meaning of the image level features corresponding to molecular markers pertinent from the urinary proteomic data. JZ contributed with the statistical analysis conducted in the study. KYJ generated ground-truth segmentation boundaries for analyzing the performance of the multi-compartment renal tissue segmentation. SSH co-conceived the overall study with PS integrating urinary proteomics data with renal tissue image data, optimized urinary proteomics data generation, as well as spearheaded the database generation with matching renal tissue whole slide image data as well as with corresponding outcome data. PS co-conceived the study with SSH, assisted in manuscript preparation, coordinated with the study team, assisted in study design, supervised the computational implementation, and critically analyzed the results.

## Funding Statement

Pinaki Sarder’s work is supported by NIH-NIDDK grant R01 DK114485, R01 DK131189, R21 DK128668, via the opportunity pool funding mechanism, namely via the glue grant mechanism of the NIH-NIDDK Kidney Precision Medicine Project (KPMP) consortium grant U2C DK114886, NIH-OD Human Biomolecular Atlas Project (HuBMAP) consortium Integration, Visualization & Engagement (HIVE) project OT2 OD033753, NIH/NCI Coordinating and Data Management Center for Acquired Resistance to Therapy Network U24 CA274159, and start-up funding from University of Florida. Seung Seok Han’s work is supported by the SNUH Research Fund (26-2018-0040, 26-2022-0040).

## Disclosures

The authors have no conflicts of interest to declare.

---

**Table 1.**
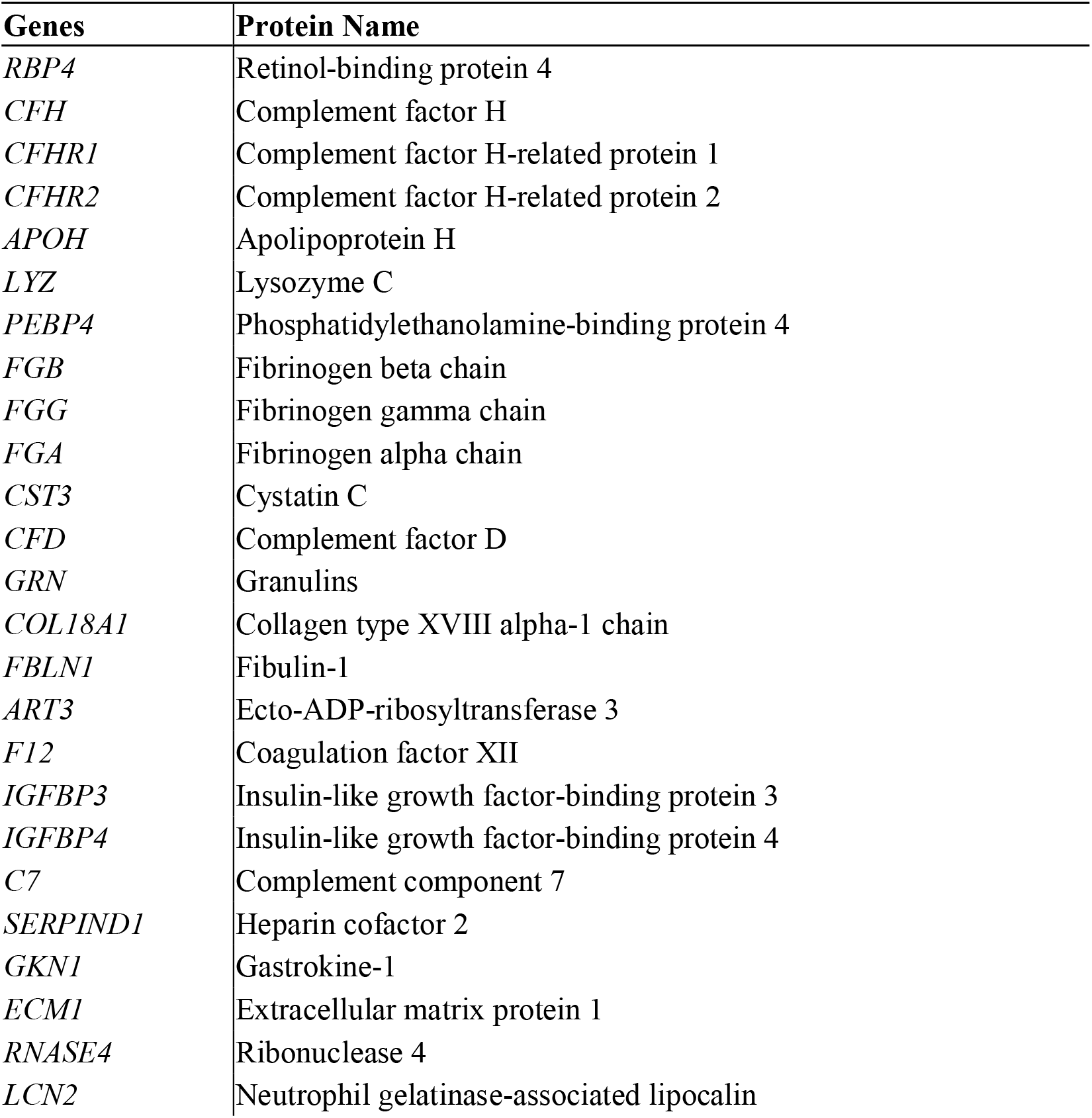
Up-regulated proteins in patients progressing to end-stage kidney disease.

**Table 2.**
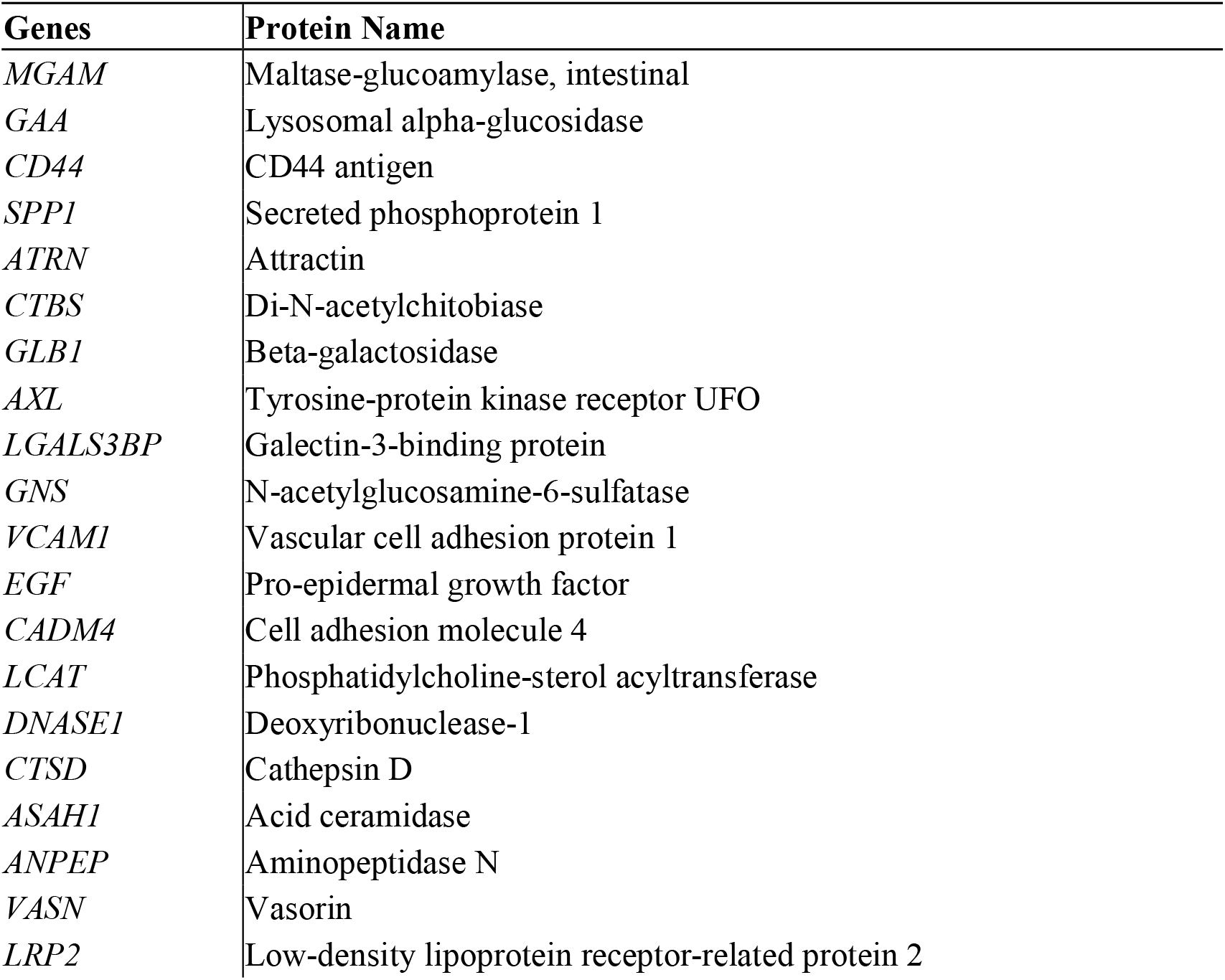
Down-regulated proteins in patients progressing to end-stage kidney disease.

| **Genes** | **Protein Name** |
| --- | --- |
| *MGAM* | Maltase-glucoamylase, intestinal |
| *GAA* | Lysosomal alpha-glucosidase |
| *CD44* | CD44 antigen |
| *SPP1* | Secreted phosphoprotein 1 |
| *ATRN* | Attractin |
| *CTBS* | Di-N-acetylchitobiase |
| *GLB1* | Beta-galactosidase |
| *AXL* | Tyrosine-protein kinase receptor UFO |
| *LGALS3BP* | Galectin-3-binding protein |
| *GNS* | N-acetylglucosamine-6-sulfatase |
| *VCAM1* | Vascular cell adhesion protein 1 |
| *EGF* | Pro-epidermal growth factor |
| *CADM4* | Cell adhesion molecule 4 |
| *LCAT* | Phosphatidylcholine-sterol acyltransferase |
| *DNASE1* | Deoxyribonuclease-1 |
| *CTSD* | Cathepsin D |
| *ASAH1* | Acid ceramidase |
| *ANPEP* | Aminopeptidase N |
| *VASN* | Vasorin |
| *LRP2* | Low-density lipoprotein receptor-related protein 2 |

## Notes

### Competing Interest Statement

The authors have declared no competing interest.

### Funding Statement

Pinaki Sarders work is supported by NIH-NIDDK grant R01 DK114485 R01 DK131189 and R21 DK128668 via the opportunity pool funding mechanism namely via the glue grant mechanism of the NIH-NIDDK Kidney Precision Medicine Project (KPMP) consortium grant U2C DK114886 NIH-OD Human Biomolecular Atlas Project (HuBMAP) consortium Integration Visualization & Engagement (HIVE) project OT2 OD033753 NIH/NCI Coordinating and Data Management Center for Acquired Resistance to Therapy Network U24 CA274159, and start-up funding from University of Florida. Seung Seok Hans work is supported by the SNUH Research Fund (26-2018-0040 26-2022-0040).

### Author Declarations

Human data collection followed protocols approved by the Institutional Review Board at the Seoul National University Hospital (H-1812-159-998).

