## Supplemental Document for "Discovery of Novel Digital Biomarkers for Type 2 Diabetic Nephropathy Classification via Integration of Urinary Proteomics and Pathology"

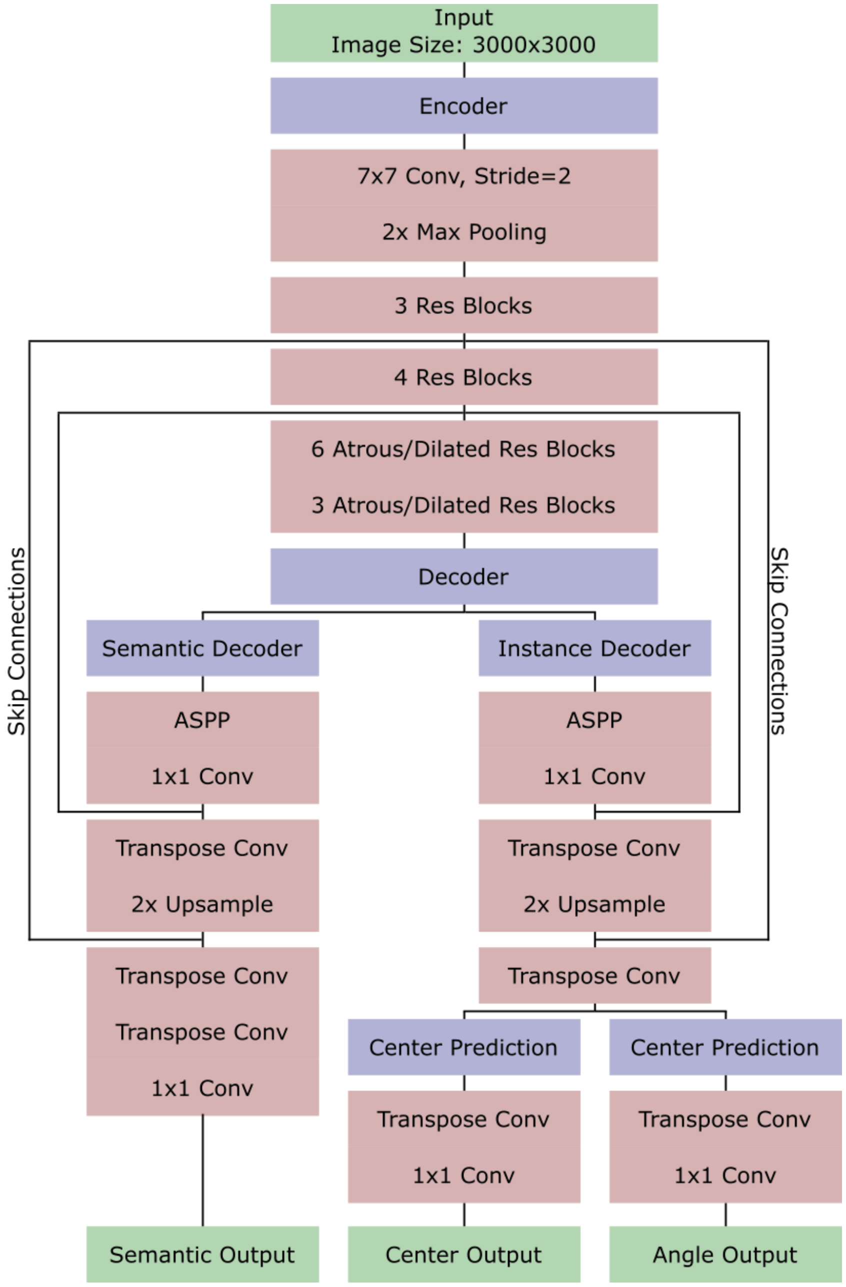


**Supp. Fig. 1. Panoptic segmentation architecture**

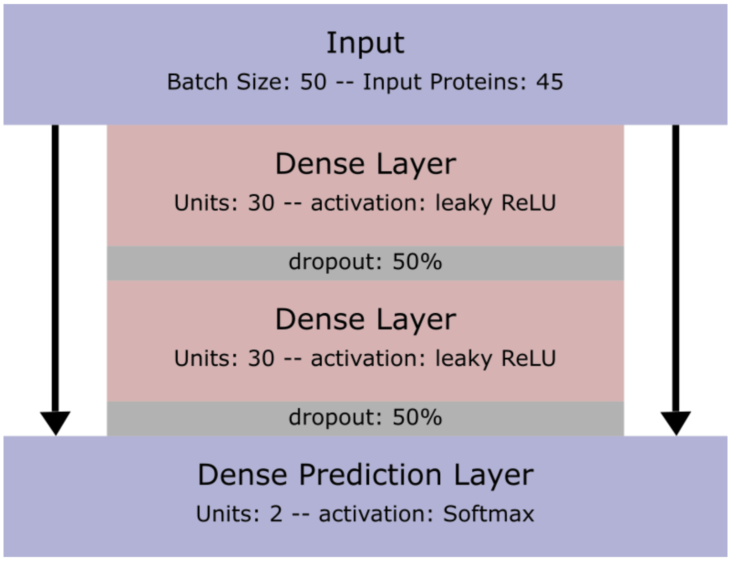


**Supp. Fig. 2. Fully connected neural network (FCNN) architecture**

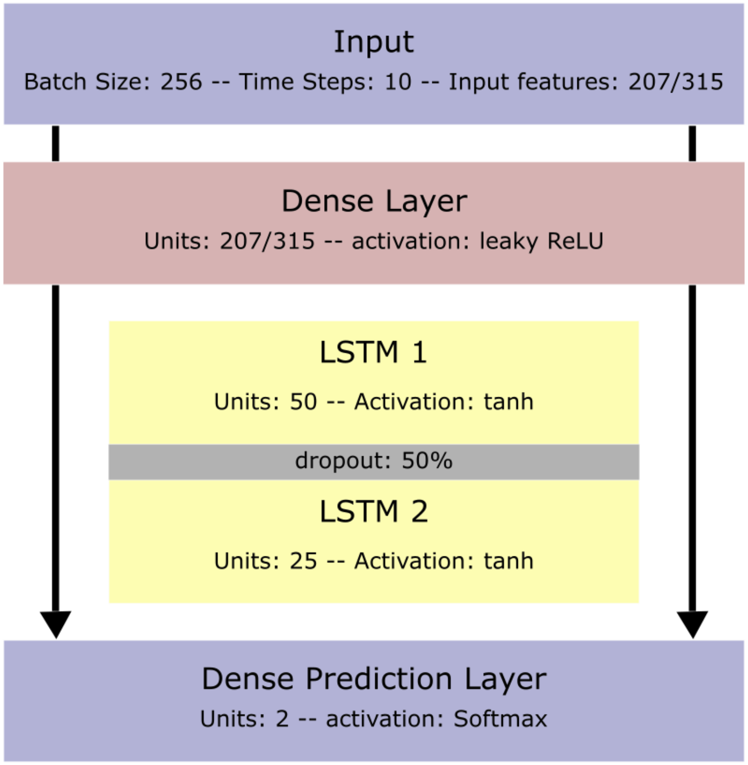


**Supp. Fig. 3. Recurrent neural network (RNN) architecture**

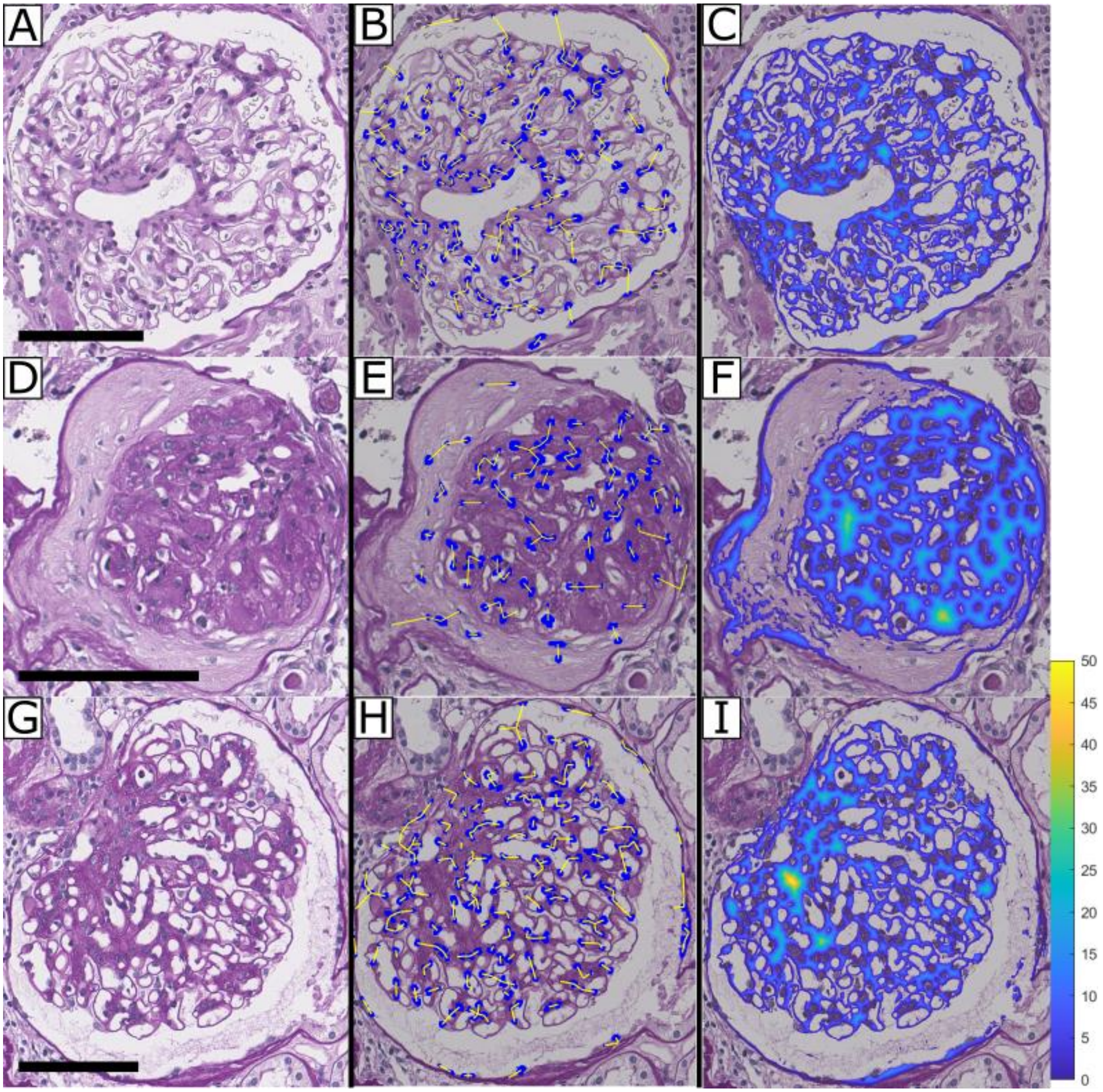


**Supp. Fig. 4. Glomerular feature extraction.** (A, D, G) Original images from WSI. (B, E, H) Representation of closest internuclear distance. (C, F, I) Representation of PAS+ thicknesses. Scale bars = 100 µm. These features have been expanded from the original manuscript by Ginley, B. et al. Computational Segmentation and Classification of Diabetic Glomerulosclerosis. J Am Soc Nephrol 30, 1953-1967 (2019). https://doi.org:10.1681/asn.2018121259

**Supp. Table 1. Baseline characteristics of the patients in the testing cohort**

| Characteristic | Total ($N$ = 56) | Non-ESKD ($n$ = 40) | ESKD ($n$ = 16) | *p Value* |
|---|---|---|---|---|
| Age (years) | 51.6 ± 10.9 | 51.1 ± 10.9 | 52.9 ± 11.2 | 0.575 |
| Male (%) | 75.0 | 75.0 | 75.0 | 1.000 |
| Body mass index (kg/m²) | 25.3 ± 3.5 | 25.4 ± 3.7 | 25.1 ± 3.3 | 0.712 |
| Duration of diabetes mellitus (years) | 10.0 (4.8–15.0) | 9.5 (3.8–14.2) | 11.5 (9.2–18.5) | 0.063 |
| Comorbidities (%) | | | | |
| Hypertension | 94.6 | 95 | 93.8 | 1.000 |
| History of ischemic heart disease | 7.1 | 7.5 | 6.2 | 1.000 |
| History of cerebrovascular disease | 3.6 | 2.5 | 6.2 | 0.494 |
| Laboratory findings | | | | |
| HbA1c (%) | 7.2 (6.6–8.0) | 7.2 (6.6–8.5) | 6.6 (6.5–7.3) | 0.059 |
| Blood urea nitrogen (mg/dl) | 24.5 (18.8–44.5) | 20.5 (16.0–31.0) | 52.5 (44.2–62.8) | <0.001 |
| Creatinine (mg/dl) | 1.54 (1.16–2.4) | 1.38 (1.09–1.58) | 4.47 (2.48–5.98) | <0.001 |
| eGFR (ml/min/1.73 m²) | 48.3 (23.7–69.9) | 56.8 (46.5–76.7) | 12.0 (9.3–24.5) | <0.001 |
| uPCR (g/g) | 5.5 (2.4–9.1) | 3.8 (1.9–7.2) | 9.0 (7.3–10.7) | 0.003 |
| Pathologic findings | | | | |
| Glomerular classification (%) | | | | 0.385 |
| I | 5.4 | 7.5 | 0 | |
| IIa | 10.7 | 12.5 | 6.2 | |
| IIb | 28.6 | 32.5 | 18.8 | |
| III | 14.3 | 15 | 12.5 | |
| IV | 41.1 | 32.5 | 62.5 | |
| IFTA score | 2 (1–3) | 2 (1–2) | 3 (3–3) | <0.001 |
| Interstitial inflammation score | 1 (1–1) | 1 (1–1) | 1 (1–1) | 0.805 |
| Arteriolar hyalinosis score | 2 (1–2) | 2 (1–2) | 2 (2–2) | 0.016 |
| Presence of large vessels (%) | 91.1 | 87.5 | 100 | 0.307 |
| Arteriosclerosis score | 1 (1–2) | 1 (1–2) | 2 (1–2) | 0.170 |

Abbreviations: ESKD, end-stage kidney disease; HbA1c, hemoglobin A1c; eGFR, estimated glomerular filtration rate; uPCR, random urine protein-to-creatinine ratio; IFTA, interstitial fibrosis and tubular atrophy.

**Supp. Table 2. Baseline characteristics of the patients in the validation cohort**

| Characteristic | Total ($n$ = 30) |
|---|---|
| Age (years) | 60.0 ± 10.9 |
| Male (%) | 66.7 |
| Body mass index (kg/m²) | 24.4 ± 3.2 |
| Duration of diabetes mellitus (years) | 10.0 (3.0–19.0) |
| Comorbidities (%) | |
| Hypertension | 96.7 |
| History of ischemic heart disease | 6.7 |
| History of cerebrovascular disease | 13.3 |
| Laboratory findings | |
| HbA1c (%) | 6.8 (6.2–8.4) |
| Blood urea nitrogen (mg/dl) | 26.0 (22.0–35.0) |
| Creatinine (mg/dl) | 1.56 (1.17–2.14) |
| eGFR (ml/min/1.73 m²) | 37.3 (21.4–59.3) |
| uPCR (g/g) | 3.1 (1.5–7.0) |
| Pathologic findings | |
| Glomerular classification (%) | |
| I | 13.3 |
| IIa | 13.3 |
| IIb | 26.7 |
| III | 16.7 |
| IV | 30.0 |
| IFTA score | 2 (1–3) |
| Interstitial inflammation score | 1 (1–1) |
| Arteriolar hyalinosis score | 1 (0–2) |
| Presence of large vessels (%) | 83.3 |
| Arteriosclerosis score | 1 (0–2) |

Abbreviations: ESKD, end-stage kidney disease; HbA1c, hemoglobin A1c; eGFR, estimated glomerular filtration rate; uPCR, random urine protein-to-creatinine ratio; IFTA, interstitial fibrosis and tubular atrophy.

**Supp. Table 3. Glomerular image features**

| Feature No. | Feature Description |
|---|---|
| 1 | Average luminal object solidity |
| 2 | Average PAS+ region contained in luminal object boundaries |
| 3 | Average nuclear region contained in luminal object boundaries |
| 4 | Sum total luminal objects' areas |
| 5 | Mean of luminal objects' areas |
| 6 | Median of luminal objects' areas |
| 7 | Luminal textural contrast |
| 8 | Luminal textural correlation |
| 9 | Luminal textural energy |
| 10 | Luminal textural homogeneity |
| 11 | Average PAS+ object solidity |
| 12 | Average lumina region contained in PAS+ object boundaries |
| 13 | Average nuclear region contained in PAS+ object boundaries |
| 14 | Sum total PAS+ objects' areas |
| 15 | Mean of PAS+ objects' areas |
| 16 | Median of PAS+ objects' areas |
| 17 | PAS+ textural contrast |
| 18 | PAS+ textural correlation |
| 19 | PAS+ textural energy |
| 20 | PAS+ textural homogeneity |
| 21 | Mean ratio of PAS+ pixels lying just outside nuclear perimeter to length of perimeter |
| 22 | Mean ratio of luminal pixels lying just outside nuclear perimeter to length of perimeter |
| 23 | Mean nuclear perimeter pixel count |
| 24 | Sum total nuclear area |
| 25 | Mean nuclear areas |
| 26 | Mode nuclear areas |
| 27 | Nuclear textural contrast |
| 28 | Nuclear textural correlation |
| 29 | Nuclear textural energy |
| 30 | Nuclear textural homogeneity |
| 31 | Mean distance of luminal object centroids from glomerular centroid |
| 32 | Mean of mean distances of luminal object centroids from glomerular boundary |
| 33 | Mean of maximum distances of luminal object centroids from glomerular boudnary |
| 34 | Mean of minimum distances of luminal object centroids from glomerular boudnary |
| 35 | Mean of mean distances of luminal object centroids from themselves |
| 36 | Mean of maximum distances of luminal object centroids from themselves |
| 37 | Mean of minimum distances of luminal object centroids from themselves |
| 38 | Mean distance of PAS+ object centroids from glomerular centroid |
| 39 | Mean of mean distances of PAS+ object centroids from glomerular boundary |

**Supp. Table 3. Glomerular image features**

| Feature No. | Feature Description |
|---|---|
| 40 | Mean of maximum distances of PAS+ object centroids from glomerular boudnary |
| 41 | Mean of minimum distances of PAS+ object centroids from glomerular boudnary |
| 42 | Mean of mean distances of PAS+ object centroids from themselves |
| 43 | Mean of maximum distances of PAS+ object centroids from themselves |
| 44 | Mean of minimum distances of PAS+ object centroids from themselves |
| 45 | Mean distance of nuclear object centroids from glomerular centroid |
| 46 | Mean of mean distances of nuclear object centroids from glomerular boundary |
| 47 | Mean of maximum distances of nuclear object centroids from glomerular boudnary |
| 48 | Mean of minimum distances of nuclear object centroids from glomerular boudnary |
| 49 | Mean of mean distances of nuclear object centroids from themselves |
| 50 | Mean of maximum distances of nuclear object centroids from themselves |
| 51 | Mean of minimum distances of nuclear object centroids from themselves |
| 52 | Total glomerular area |
| 53 | Total PAS+ object number |
| 54 | Total luminal object number |
| 55 | Total nucleus number |
| 56 | Sum of PAS+ distance transform values 0<d<=10 |
| 57 | Sum of PAS+ distance transform values 10<d<=20 |
| 58 | Sum of PAS+ distance transform values 20<d<=1000 |
| 59 | Maximum PAS+ distance transform value 10<d<=20 |
| 60 | Number of connected objects with PAS+ distance transform value 0<d<=10 |
| 61 | Number of connected objects with PAS+ distance transform value 10<d<=20 |
| 62 | Mean of PAS+ distance transform values 0<d<=10 |
| 63 | Mean of PAS+ distance transform values 10<d<=20 |
| 64 | Median of PAS+ distance transform values 0<d<=10 |
| 65 | Median of PAS+ distance transform values 10<d<=20 |
| 66 | Mean area of objects with PAS+ distance transform value 0<d<=10 |
| 67 | Median area of objects with PAS+ distance transform value 0<d<=10 |
| 68 | Maximum area of objects with PAS+ distance transform value 0<d<=10 |
| 69 | Mean area of objects with PAS+ distance transform value 10<d<=20 |
| 70 | Median area of objects with PAS+ distance transform value 10<d<=20 |
| 71 | Count of pixels with PAS+ distance transform value 1<d<=3 |
| 72 | Count of pixels with PAS+ distance transform value 3<d<=5 |
| 73 | Count of pixels with PAS+ distance transform value 5<d<=7 |
| 74 | Count of pixels with PAS+ distance transform value 7<d<=9 |
| 75 | Count of pixels with PAS+ distance transform value 9<d<=11 |
| 76 | Count of pixels with PAS+ distance transform value 11<d<=13 |
| 77 | Count of pixels with PAS+ distance transform value 13<d<=15 |
| 78 | Count of pixels with PAS+ distance transform value 15<d<=17 |

**Supp. Table 3. Glomerular image features**

| Feature No. | Feature Description |
|---|---|
| 79 | Count of pixels with PAS+ distance transform value 17<d<=19 |
| 80 | Count of pixels with PAS+ distance transform value 19<d<=21 |
| 81 | Count of pixels with PAS+ distance transform value 21<d<=23 |
| 82 | Count of pixels with PAS+ distance transform value 23<d<=25 |
| 83 | Count of pixels with PAS+ distance transform value 25<d<=27 |
| 84 | Count of pixels with PAS+ distance transform value 27<d<=29 |
| 85 | Count of pixels with PAS+ distance transform value 29<d<=31 |
| 86 | Count of pixels with PAS+ distance transform value 31<d<=33 |
| 87 | Count of pixels with PAS+ distance transform value 33<d<=35 |
| 88 | Count of pixels with PAS+ distance transform value 35<d<=37 |
| 89 | Count of pixels with PAS+ distance transform value 37<d<=39 |
| 90 | Count of pixels with PAS+ distance transform value 39<d<=41 |
| 91 | Count of pixels with PAS+ distance transform value 41<d<=43 |
| 92 | Count of pixels with PAS+ distance transform value 43<d<=45 |
| 93 | Count of pixels with PAS+ distance transform value 45<d<=47 |
| 94 | Count of pixels with PAS+ distance transform value 47<d<=49 |
| 95 | Count of pixels with PAS+ distance transform value 49<d<=51 |
| 96 | Count of pixels with PAS+ distance transform value 51<d<=53 |
| 97 | Count of pixels with PAS+ distance transform value 53<d<=55 |
| 98 | Count of pixels with PAS+ distance transform value 55<d<=57 |
| 99 | Count of pixels with PAS+ distance transform value 57<d<=59 |
| 100 | Count of pixels with PAS+ distance transform value 59<d<=61 |
| 101 | Count of pixels with PAS+ distance transform value 61<d<=63 |
| 102 | Count of pixels with PAS+ distance transform value 63<d<=65 |
| 103 | Count of pixels with PAS+ distance transform value 65<d<=67 |
| 104 | Count of pixels with PAS+ distance transform value 67<d<=69 |
| 105 | Count of pixels with PAS+ distance transform value 69<d<=71 |
| 106 | Count of pixels with PAS+ distance transform value 71<d<=73 |
| 107 | Count of pixels with PAS+ distance transform value 73<d<=75 |
| 108 | Count of pixels with PAS+ distance transform value 75<d<=77 |
| 109 | Count of pixels with PAS+ distance transform value 77<d<=79 |
| 110 | Count of pixels with PAS+ distance transform value 79<d<=2000 |
| 111 | Count of pixels with luminal distance transform value 1<d<=2 |
| 112 | Count of pixels with luminal distance transform value 2<d<=3 |
| 113 | Count of pixels with luminal distance transform value 3<d<=4 |
| 114 | Count of pixels with luminal distance transform value 4<d<=5 |
| 115 | Count of pixels with luminal distance transform value 5<d<=6 |
| 116 | Count of pixels with luminal distance transform value 6<d<=7 |
| 117 | Count of pixels with luminal distance transform value 7<d<=8 |

**Supp. Table 3. Glomerular image features**

| Feature No. | Feature Description |
|---|---|
| 118 | Count of pixels with luminal distance transform value 8<d<=9 |
| 119 | Count of pixels with luminal distance transform value 9<d<=10 |
| 120 | Count of pixels with luminal distance transform value 10<d<=11 |
| 121 | Count of pixels with luminal distance transform value 11<d<=12 |
| 122 | Count of pixels with luminal distance transform value 12<d<=13 |
| 123 | Count of pixels with luminal distance transform value 13<d<=14 |
| 124 | Count of pixels with luminal distance transform value 14<d<=15 |
| 125 | Count of pixels with luminal distance transform value 15<d<=16 |
| 126 | Count of pixels with luminal distance transform value 16<d<=17 |
| 127 | Count of pixels with luminal distance transform value 17<d<=18 |
| 128 | Count of pixels with luminal distance transform value 18<d<=19 |
| 129 | Count of pixels with luminal distance transform value 19<d<=20 |
| 130 | Count of pixels with luminal distance transform value 20<d<=21 |
| 131 | Count of pixels with luminal distance transform value 21<d<=22 |
| 132 | Count of pixels with luminal distance transform value 22<d<=23 |
| 133 | Count of pixels with luminal distance transform value 23<d<=24 |
| 134 | Count of pixels with luminal distance transform value 24<d<=25 |
| 135 | Count of pixels with luminal distance transform value 25<d<=26 |
| 136 | Count of pixels with luminal distance transform value 26<d<=27 |
| 137 | Count of pixels with luminal distance transform value 27<d<=28 |
| 138 | Count of pixels with luminal distance transform value 28<d<=29 |
| 139 | Count of pixels with luminal distance transform value 29<d<=30 |
| 140 | Count of pixels with luminal distance transform value 30<d<=31 |
| 141 | Count of pixels with luminal distance transform value 31<d<=32 |
| 142 | Count of pixels with luminal distance transform value 32<d<=33 |
| 143 | Count of pixels with luminal distance transform value 33<d<=34 |
| 144 | Count of pixels with luminal distance transform value 34<d<=35 |
| 145 | Count of pixels with luminal distance transform value 35<d<=36 |
| 146 | Count of pixels with luminal distance transform value 36<d<=37 |
| 147 | Count of pixels with luminal distance transform value 37<d<=38 |
| 148 | Count of pixels with luminal distance transform value 38<d<=39 |
| 149 | Count of pixels with luminal distance transform value 39<d<=40 |
| 150 | Count of pixels with luminal distance transform value 40<d<=41 |
| 151 | Count of pixels with luminal distance transform value 41<d<=42 |
| 152 | Count of pixels with luminal distance transform value 42<d<=43 |
| 153 | Count of pixels with luminal distance transform value 43<d<=44 |
| 154 | Count of pixels with luminal distance transform value 44<d<=45 |
| 155 | Count of pixels with luminal distance transform value 45<d<=46 |
| 156 | Count of pixels with luminal distance transform value 46<d<=47 |

**Supp. Table 3. Glomerular image features**

| Feature No. | Feature Description |
|---:|---|
| 157 | Count of pixels with luminal distance transform value 47<d<=48 |
| 158 | Count of pixels with luminal distance transform value 48<d<=49 |
| 159 | Count of pixels with luminal distance transform value 49<d<=50 |
| 160 | Count of pixels with luminal distance transform value 50<d<=51 |
| 161 | Count of pixels with luminal distance transform value 51<d<=52 |
| 162 | Count of pixels with luminal distance transform value 52<d<=53 |
| 163 | Count of pixels with luminal distance transform value 53<d<=54 |
| 164 | Count of pixels with luminal distance transform value 54<d<=55 |
| 165 | Count of pixels with luminal distance transform value 55<d<=56 |
| 166 | Count of pixels with luminal distance transform value 56<d<=57 |
| 167 | Count of pixels with luminal distance transform value 57<d<=58 |
| 168 | Count of pixels with luminal distance transform value 58<d<=59 |
| 169 | Count of pixels with luminal distance transform value 59<d<=60 |
| 170 | Count of pixels with luminal distance transform value 60<d<=2000 |
| 171 | Count of pixels with nuclear distance transform value 1<d<=2 |
| 172 | Count of pixels with nuclear distance transform value 2<d<=3 |
| 173 | Count of pixels with nuclear distance transform value 3<d<=4 |
| 174 | Count of pixels with nuclear distance transform value 4<d<=5 |
| 175 | Count of pixels with nuclear distance transform value 5<d<=6 |
| 176 | Count of pixels with nuclear distance transform value 6<d<=7 |
| 177 | Count of pixels with nuclear distance transform value 7<d<=8 |
| 178 | Count of pixels with nuclear distance transform value 8<d<=9 |
| 179 | Count of pixels with nuclear distance transform value 9<d<=10 |
| 180 | Count of pixels with nuclear distance transform value 10<d<=11 |
| 181 | Count of pixels with nuclear distance transform value 11<d<=12 |
| 182 | Count of pixels with nuclear distance transform value 12<d<=13 |
| 183 | Count of pixels with nuclear distance transform value 13<d<=14 |
| 184 | Count of pixels with nuclear distance transform value 14<d<=15 |
| 185 | Count of pixels with nuclear distance transform value 15<d<=16 |
| 186 | Count of pixels with nuclear distance transform value 16<d<=17 |
| 187 | Count of pixels with nuclear distance transform value 17<d<=18 |
| 188 | Count of pixels with nuclear distance transform value 18<d<=19 |
| 189 | Count of pixels with nuclear distance transform value 19<d<=20 |
| 190 | Count of pixels with nuclear distance transform value 20<d<=2000 |
| 191 | Count of pixels with glomerular distance transform value 2<d<=27 |
| 192 | Count of pixels with glomerular distance transform value 27<d<=52 |
| 193 | Count of pixels with glomerular distance transform value 52<d<=77 |
| 194 | Count of pixels with glomerular distance transform value 77<d<=102 |
| 195 | Count of pixels with glomerular distance transform value 102<d<=127 |

**Supp. Table 3. Glomerular image features**

| Feature No. | Feature Description |
|---|---|
| 196 | Count of pixels with glomerular distance transform value 127<d<=152 |
| 197 | Count of pixels with glomerular distance transform value 152<d<=177 |
| 198 | Count of pixels with glomerular distance transform value 177<d<=202 |
| 199 | Count of pixels with glomerular distance transform value 202<d<=227 |
| 200 | Count of pixels with glomerular distance transform value 227<d<=252 |
| 201 | Count of pixels with glomerular distance transform value 252<d<=277 |
| 202 | Count of pixels with glomerular distance transform value 277<d<=302 |
| 203 | Count of pixels with glomerular distance transform value 302<d<=327 |
| 204 | Count of pixels with glomerular distance transform value 327<d<=352 |
| 205 | Count of pixels with glomerular distance transform value 352<d<=377 |
| 206 | Count of pixels with glomerular distance transform value 377<d<=402 |
| 207 | Count of pixels with glomerular distance transform value 402<d<=427 |
| 208 | Count of pixels with glomerular distance transform value 427<d<=452 |
| 209 | Count of pixels with glomerular distance transform value 452<d<=477 |
| 210 | Count of pixels with glomerular distance transform value 477<d<=502 |
| 211 | Count of pixels with glomerular distance transform value 502<d<=527 |
| 212 | Count of pixels with glomerular distance transform value 527<d<=552 |
| 213 | Count of pixels with glomerular distance transform value 552<d<=577 |
| 214 | Count of pixels with glomerular distance transform value 577<d<=20000 |
| 215 | Mean of red values in PAS+ regions |
| 216 | Mean of green values in PAS+ regions |
| 217 | Mean of blue values in PAS+ regions |
| 218 | Std of red values in PAS+ regions |
| 219 | Std of green values in PAS+ regions |
| 220 | Std of blue values in PAS+ regions |
| 221 | Mean of red values in luminal regions |
| 222 | Mean of green values in luminal regions |
| 223 | Mean of blue values in luminal regions |
| 224 | Std of red values in luminal regions |
| 225 | Std of green values in luminal regions |
| 226 | Std of blue values in luminal regions |
| 227 | Mean of red values in nuclear regions |
| 228 | Mean of green values in nuclear regions |
| 229 | Mean of blue values in nuclear regions |
| 230 | Std of red values in nuclear regions |
| 231 | Std of green values in nuclear regions |
| 232 | Std of blue values in nuclear regions |
| 233 | Number nuclear pixels contained radius 0<r<100 |
| 234 | Number nuclear pixels contained radius 100<r<200 |

**Supp. Table 3. Glomerular image features**

| Feature No. | Feature Description |
|---:|---|
| 235 | Number nuclear pixels contained radius 200<r<300 |
| 236 | Number nuclear pixels contained radius 300<r<400 |
| 237 | Number nuclear pixels contained radius 400<r<500 |
| 238 | Number nuclear pixels contained radius 500<r<600 |
| 239 | Number nuclear pixels contained radius 600<r<700 |
| 240 | Number nuclear pixels contained radius 700<r<800 |
| 241 | Number nuclear pixels contained radius 800<r<900 |
| 242 | Number nuclear pixels contained radius 900<r<1000 |
| 243 | Number nuclear pixels contained radius 1000<r<1300 |
| 244 | Number luminal pixels contained radius 0<r<100 |
| 245 | Number luminal pixels contained radius 100<r<200 |
| 246 | Number luminal pixels contained radius 200<r<300 |
| 247 | Number luminal pixels contained radius 300<r<400 |
| 248 | Number luminal pixels contained radius 400<r<500 |
| 249 | Number luminal pixels contained radius 500<r<600 |
| 250 | Number luminal pixels contained radius 600<r<700 |
| 251 | Number luminal pixels contained radius 700<r<800 |
| 252 | Number luminal pixels contained radius 800<r<900 |
| 253 | Number luminal pixels contained radius 900<r<1000 |
| 254 | Number luminal pixels contained radius 1000<r<1300 |
| 255 | Number PAS+ pixels contained radius 0<r<100 |
| 256 | Number PAS+ pixels contained radius 100<r<200 |
| 257 | Number PAS+ pixels contained radius 200<r<300 |
| 258 | Number PAS+ pixels contained radius 300<r<400 |
| 259 | Number PAS+ pixels contained radius 400<r<500 |
| 260 | Number PAS+ pixels contained radius 500<r<600 |
| 261 | Number PAS+ pixels contained radius 600<r<700 |
| 262 | Number PAS+ pixels contained radius 700<r<800 |
| 263 | Number PAS+ pixels contained radius 800<r<900 |
| 264 | Number PAS+ pixels contained radius 900<r<1000 |
| 265 | Number PAS+ pixels contained radius 1000<r<1300 |
| 266 | Number nuclear pixels contained between theta -180<ϴ<-162 |
| 267 | Number nuclear pixels contained between theta -162<ϴ<-144 |
| 268 | Number nuclear pixels contained between theta -144<ϴ<-126 |
| 269 | Number nuclear pixels contained between theta -126<ϴ<-108 |
| 270 | Number nuclear pixels contained between theta -108<ϴ<-90 |
| 271 | Number nuclear pixels contained between theta -90<ϴ<-72 |
| 272 | Number nuclear pixels contained between theta -72<ϴ<-54 |
| 273 | Number nuclear pixels contained between theta -54<ϴ<-36 |

**Supp. Table 3. Glomerular image features**

| Feature No. | Feature Description |
|---|---|
| 274 | Number nuclear pixels contained between theta $-36<\Theta<-18$ |
| 275 | Number nuclear pixels contained between theta $-18<\Theta<0$ |
| 276 | Number nuclear pixels contained between theta $0<\Theta<18$ |
| 277 | Number nuclear pixels contained between theta $18<\Theta<36$ |
| 278 | Number nuclear pixels contained between theta $36<\Theta<54$ |
| 279 | Number nuclear pixels contained between theta $54<\Theta<72$ |
| 280 | Number nuclear pixels contained between theta $72<\Theta<90$ |
| 281 | Number nuclear pixels contained between theta $90<\Theta<108$ |
| 282 | Number nuclear pixels contained between theta $108<\Theta<126$ |
| 283 | Number nuclear pixels contained between theta $126<\Theta<144$ |
| 284 | Number nuclear pixels contained between theta $144<\Theta<162$ |
| 285 | Number nuclear pixels contained between theta $162<\Theta<180$ |
| 286 | 0.1 quantile of nuclear pixels from the boundary line |
| 287 | 0.2 quantile of nuclear pixels from the boundary line |
| 288 | 0.3 quantile of nuclear pixels from the boundary line |
| 289 | 0.4 quantile of nuclear pixels from the boundary line |
| 290 | 0.5 quantile of nuclear pixels from the boundary line |
| 291 | 0.6 quantile of nuclear pixels from the boundary line |
| 292 | 0.7 quantile of nuclear pixels from the boundary line |
| 293 | 0.8 quantile of nuclear pixels from the boundary line |
| 294 | 0.9 quantile of nuclear pixels from the boundary line |
| 295 | 1 quantile of nuclear pixels from the boundary line |
| 296 | 0.1 quantile of luminal pixels from the boundary line |
| 297 | 0.2 quantile of luminal pixels from the boundary line |
| 298 | 0.3 quantile of luminal pixels from the boundary line |
| 299 | 0.4 quantile of luminal pixels from the boundary line |
| 300 | 0.5 quantile of luminal pixels from the boundary line |
| 301 | 0.6 quantile of luminal pixels from the boundary line |
| 302 | 0.7 quantile of luminal pixels from the boundary line |
| 303 | 0.8 quantile of luminal pixels from the boundary line |
| 304 | 0.9 quantile of luminal pixels from the boundary line |
| 305 | 1 quantile of luminal pixels from the boundary line |
| 306 | 0.1 quantile of mesangial pixels from the boundary line |
| 307 | 0.2 quantile of mesangial pixels from the boundary line |
| 308 | 0.3 quantile of mesangial pixels from the boundary line |
| 309 | 0.4 quantile of mesangial pixels from the boundary line |
| 310 | 0.5 quantile of mesangial pixels from the boundary line |
| 311 | 0.6 quantile of mesangial pixels from the boundary line |
| 312 | 0.7 quantile of mesangial pixels from the boundary line |

**Supp. Table 3. Glomerular image features**

| Feature No. | Feature Description |
|---|---|
| 313 | 0.8 quantile of mesangial pixels from the boundary line |
| 314 | 0.9 quantile of mesangial pixels from the boundary line |
| 315 | 1 quantile of mesangial pixels from the boundary line |

**Supp. Table 4. Tubular image features**

| Feature No. | Feature Description |
|---|---|
| 1 | Average luminal object solidity |
| 2 | Average PAS+ region contained in luminal object boundaries |
| 3 | Average nuclear region contained in luminal object boundaries |
| 4 | Sum total luminal objects' areas |
| 5 | Mean of luminal objects' areas |
| 6 | Median of luminal objects' areas |
| 7 | Luminal textural contrast |
| 8 | Luminal textural correlation |
| 9 | Luminal textural energy |
| 10 | Luminal textural homogeneity |
| 11 | Average PAS+ object solidity |
| 12 | Average lumina region contained in PAS+ object boundaries |
| 13 | Average nuclear region contained in PAS+ object boundaries |
| 14 | Sum total PAS+ objects' areas |
| 15 | Mean of PAS+ objects' areas |
| 16 | Median of PAS+ objects' areas |
| 17 | PAS+ textural contrast |
| 18 | PAS+ textural correlation |
| 19 | PAS+ textural energy |
| 20 | PAS+ textural homogeneity |
| 21 | Mean ratio of PAS+ pixels lying just outside nuclear perimeter to length of perimeter |
| 22 | Mean ratio of luminal pixels lying just outside nuclear perimeter to length of perimeter |
| 23 | Mean nuclear perimeter pixel count |
| 24 | Sum total nuclear area |
| 25 | Mean nuclear areas |
| 26 | Mode nuclear areas |
| 27 | Nuclear textural contrast |
| 28 | Nuclear textural correlation |
| 29 | Nuclear textural energy |
| 30 | Nuclear textural homogeneity |
| 31 | Mean distance of nuclear object centroids from tubular centroid |

**Supp. Table 4. Tubular image features**

| Feature No. | Feature Description |
|---|---|
| 32 | Mean of mean distances of nuclear object centroids from tubular boundary |
| 33 | Mean of maximum distances of nuclear object centroids from tubular boundary |
| 34 | Mean of minimum distances of nuclear object centroids from tubular boundary |
| 35 | Mean of mean distances of nuclear object centroids from themselves |
| 36 | Mean of maximum distances of nuclear object centroids from themselves |
| 37 | Mean of minimum distances of nuclear object centroids from themselves |
| 38 | Total tubular area |
| 39 | Total PAS+ object number |
| 40 | Total luminal object number |
| 41 | Total nucleus number |
| 42 | Average TBM thickness per perimeter length |
| 43 | Maximum TBM thickness per perimeter length |
| 44 | Minimum TBM thickness per perimeter length |
| 45 | Average value of TBM distance transform |
| 46 | Maximum value of TBM distance transform |
| 47 | TBM textural energy |
| 48 | TBM textural correlation |
| 49 | TBM textural contrast |
| 50 | TBM textural homogeneity |
| 51 | Mean of red values in TBM regions |
| 52 | Mean of green values in TBM regions |
| 53 | Mean of blue values in TBM regions |
| 54 | Standard deviation of red values in TBM regions |
| 55 | Standard deviation of green values in TBM regions |
| 56 | Standard deviation of blue values in TBM regions |
| 57 | Sum total TBM area |
| 58 | Average solidity of TBM |
| 59 | Mean distance of TBM object centroids from tubular centroid |
| 60 | Mean of mean distances of TBM object centroids from tubular boundary |
| 61 | Mean of maximum distances of TBM object centroids from tubular boundary |
| 62 | Mean of minimum distances of TBM object centroids from tubular boundary |
| 63 | Mean of mean distances of TBM object centroids from themselves |
| 64 | Mean of maximum distances of TBM object centroids from themselves |
| 65 | Mean of minimum distances of TBM object centroids from themselves |
| 66 | Average ITO thickness per perimeter length |
| 67 | Maximum ITO thickness per perimeter length |
| 68 | Minimum ITO thickness per perimeter length |
| 69 | Average value of ITO distance transform |
| 70 | Maximum value of ITO distance transform |

**Supp. Table 4. Tubular image features**

| Feature No. | Feature Description |
|---|---|
| 71 | ITO textural energy |
| 72 | ITO textural correlation |
| 73 | ITO textural contrast |
| 74 | ITO textural homogeneity |
| 75 | Mean of red values in ITO regions |
| 76 | Mean of green values in ITO regions |
| 77 | Mean of blue values in ITO regions |
| 78 | Standard deviation of red values in ITO regions |
| 79 | Standard deviation of green values in ITO regions |
| 80 | Standard deviation of blue values in ITO regions |
| 81 | Standard deviation of blue values in ITO regions |
| 82 | Sum total ITO area |
| 83 | Average solidity of ITO |
| 84 | Mean distance of ITO object centroids from tubular centroid |
| 85 | Mean of mean distances of ITO object centroids from tubular boundary |
| 86 | Mean of maximum distances of ITO object centroids from tubular boundary |
| 87 | Mean of minimum distances of ITO object centroids from tubular boundary |
| 88 | Mean of mean distances of ITO object centroids from themselves |
| 89 | Mean of maximum distances of ITO object centroids from themselves |
| 90 | Mean of minimum distances of ITO object centroids from themselves |
| 91 | Tubular compactness |
| 92 | Tubular eccentricity |
| 93 | Equivalent diameter of a circle with measured tubular area |
| 94 | Tubule major axis length |
| 95 | Tubule minor axis length |
| 96 | Tubular perimeter |
| 97 | Tubular fiber length |
| 98 | Tubular fiber width |
| 99 | Tubular curl |
| 100 | Tubular solidity |
| 101 | Mean of red values in PAS+ regions |
| 102 | Mean of green values in PAS+ regions |
| 103 | Mean of blue values in PAS+ regions |
| 104 | Std of red values in PAS+ regions |
| 105 | Std of green values in PAS+ regions |
| 106 | Std of blue values in PAS+ regions |
| 107 | Mean of red values in luminal regions |
| 108 | Mean of green values in luminal regions |
| 109 | Mean of blue values in luminal regions |

**Supp. Table 4. Tubular image features**

| Feature No. | Feature Description |
|---|---|
| 110 | Std of red values in luminal regions |
| 111 | Std of green values in luminal regions |
| 112 | Std of blue values in luminal regions |
| 113 | Mean of red values in nuclear regions |
| 114 | Mean of green values in nuclear regions |
| 115 | Mean of blue values in nuclear regions |
| 116 | Std of red values in nuclear regions |
| 117 | Std of green values in nuclear regions |
| 118 | Std of blue values in nuclear regions |
| 119 | Minimum of luminal pixels weighted inversely by distance from tubular boundary |
| 120 | Maximum of luminal pixels weighted inversely by distance from tubular boundary |
| 121 | Mean of luminal pixels weighted inversely by distance from tubular boundary |
| 122 | Median of luminal pixels weighted inversely by distance from tubular boundary |
| 123 | Standard deviation of luminal pixels weighted inversely by distance from tubular boundary |
| 124 | Minimum of PAS+ pixels weighted inversely by distance from tubular boundary |
| 125 | Maximum of PAS+ pixels weighted inversely by distance from tubular boundary |
| 126 | Mean of PAS+ pixels weighted inversely by distance from tubular boundary |
| 127 | Median of PAS+ pixels weighted inversely by distance from tubular boundary |
| 128 | Standard deviation of PAS+ pixels weighted inversely by distance from tubular boundary |
| 129 | 0.1 quantile of nuclear pixels from the boundary line |
| 130 | 0.2 quantile of nuclear pixels from the boundary line |
| 131 | 0.3 quantile of nuclear pixels from the boundary line |
| 132 | 0.4 quantile of nuclear pixels from the boundary line |
| 133 | 0.5 quantile of nuclear pixels from the boundary line |
| 134 | 0.6 quantile of nuclear pixels from the boundary line |
| 135 | 0.7 quantile of nuclear pixels from the boundary line |
| 136 | 0.8 quantile of nuclear pixels from the boundary line |
| 137 | 0.9 quantile of nuclear pixels from the boundary line |
| 138 | 1 quantile of nuclear pixels from the boundary line |
| 139 | 0.1 quantile of luminal pixels from the boundary line |
| 140 | 0.2 quantile of luminal pixels from the boundary line |
| 141 | 0.3 quantile of luminal pixels from the boundary line |
| 142 | 0.4 quantile of luminal pixels from the boundary line |
| 143 | 0.5 quantile of luminal pixels from the boundary line |
| 144 | 0.6 quantile of luminal pixels from the boundary line |
| 145 | 0.7 quantile of luminal pixels from the boundary line |
| 146 | 0.8 quantile of luminal pixels from the boundary line |
| 147 | 0.9 quantile of luminal pixels from the boundary line |
| 148 | 1 quantile of luminal pixels from the boundary line |

**Supp. Table 4. Tubular image features**

| Feature No. | Feature Description |
|---|---|
| 149 | 0.1 quantile of mesangial pixels from the boundary line |
| 150 | 0.2 quantile of mesangial pixels from the boundary line |
| 151 | 0.3 quantile of mesangial pixels from the boundary line |
| 152 | 0.4 quantile of mesangial pixels from the boundary line |
| 153 | 0.5 quantile of mesangial pixels from the boundary line |
| 154 | 0.6 quantile of mesangial pixels from the boundary line |
| 155 | 0.7 quantile of mesangial pixels from the boundary line |
| 156 | 0.8 quantile of mesangial pixels from the boundary line |
| 157 | 0.9 quantile of mesangial pixels from the boundary line |
| 158 | 1 quantile of mesangial pixels from the boundary line |
| 159 | Maximum value of PAS+ distance transform |
| 160 | Maximum value of luminal distance transform |
| 161 | Maximum value of nuclear distance transform |
| 162 | Maximum value of tubular distance transform |
| 163 | Maximum of mean pixel distance from boundary per nucleus |
| 164 | Maximum of minimum pixel distance from boundary per nucleus |
| 165 | Maximum of maximum pixel distance from boundary per nucleus |
| 166 | Mean of mean pixel distance from boundary per nucleus |
| 167 | Mean of minimum pixel distance from boundary per nucleus |
| 168 | Mean of maximum pixel distance from boundary per nucleus |
| 169 | Minimum of mean pixel distance from boundary per nucleus |
| 170 | Minimum of minimum pixel distance from boundary per nucleus |
| 171 | Minimum of maximum pixel distance from boundary per nucleus |
| 172 | Variance of mean pixel distance from boundary per nucleus |
| 173 | Variance of minimum pixel distance from boundary per nucleus |
| 174 | Variance of maximum pixel distance from boundary per nucleus |
| 175 | Median of mean pixel distance from boundary per nucleus |
| 176 | Median of minimum pixel distance from boundary per nucleus |
| 177 | Median of maximum pixel distance from boundary per nucleus |
| 178 | Maximum of mean pixel distance from boundary per TBM object |
| 179 | Maximum of minimum pixel distance from boundary per TBM object |
| 180 | Maximum of maximum pixel distance from boundary per TBM object |
| 181 | Mean of mean pixel distance from boundary per TBM object |
| 182 | Mean of minimum pixel distance from boundary per TBM object |
| 183 | Mean of maximum pixel distance from boundary per TBM object |
| 184 | Minimum of mean pixel distance from boundary per TBM object |
| 185 | Minimum of minimum pixel distance from boundary per TBM object |
| 186 | Minimum of maximum pixel distance from boundary per TBM object |
| 187 | Variance of mean pixel distance from boundary per TBM object |

**Supp. Table 4. Tubular image features**

| Feature No. | Feature Description |
|---|---|
| 188 | Variance of minimum pixel distance from boundary per TBM object |
| 189 | Variance of maximum pixel distance from boundary per TBM object |
| 190 | Median of mean pixel distance from boundary per TBM object |
| 191 | Median of minimum pixel distance from boundary per TBM object |
| 192 | Median of maximum pixel distance from boundary per TBM object |
| 193 | Maximum of mean pixel distance from boundary per ITO |
| 194 | Maximum of minimum pixel distance from boundary per ITO |
| 195 | Maximum of maximum pixel distance from boundary per ITO |
| 196 | Mean of mean pixel distance from boundary per ITO |
| 197 | Mean of minimum pixel distance from boundary per ITO |
| 198 | Mean of maximum pixel distance from boundary per ITO |
| 199 | Minimum of mean pixel distance from boundary per ITO |
| 200 | Minimum of minimum pixel distance from boundary per ITO |
| 201 | Minimum of maximum pixel distance from boundary per ITO |
| 202 | Variance of mean pixel distance from boundary per ITO |
| 203 | Variance of minimum pixel distance from boundary per ITO |
| 204 | Variance of maximum pixel distance from boundary per ITO |
| 205 | Median of mean pixel distance from boundary per ITO |
| 206 | Median of minimum pixel distance from boundary per ITO |
| 207 | Median of maximum pixel distance from boundary per ITO |

**Supp. Table 5. All urinary proteins measured**

| Protein No. | Protein names | Gene names |
|---|---|---|
| 1 | | |
| 2 | | |
| 3 | Nuclear speckle splicing regulatory protein 1 | CCDC55 |
| 4 | Serpin B6 | SERPINB6 |
| 5 | EH domain-containing protein 1 | EHD1 |
| 6 | Sushi, von Willebrand factor type A, EGF and pentraxin domain-containing protein 1 | SVEP1 |
| 7 | Ig kappa chain V-III region VH | IGKV3-7 |
| 8 | | IGLV4-69 |
| 9 | | IGLV8-61 |
| 10 | | IGLV4-60 |
| 11 | | IGLV6-57 |
| 12 | Ig lambda chain V-I region NIG-64 | IGLV1-51 |
| 13 | | IGLV1-47 |
| 14 | | IGLV7-46 |
| 15 | | IGLV1-40 |

**Supp. Table 5. All urinary proteins measured**

| Protein No. | Protein names | Gene names |
|---|---|---|
| 16 | | IGLV5-37 |
| 17 | | IGLV3-27 |
| 18 | | IGLV3-25 |
| 19 | | IGLV3-21 |
| 20 | | IGLV3-19 |
| 21 | | IGLV2-18 |
| 22 | | IGLV2-14 |
| 23 | | IGLV2-11 |
| 24 | | IGLV3-10 |
| 25 | | IGLV3-9 |
| 26 | Ig lambda-2 chain C regions | IGLC2 |
| 27 | Ig lambda-7 chain C region | IGLC7 |
| 28 | | IGHA2 |
| 29 | | IGHV3-64 |
| 30 | | IGHV4-4 |
| 31 | | IGKV2D-24 |
| 32 | Ig kappa chain V-II region RPMI 6410 | IGKV2-30 |
| 33 | | IGKV2D-30 |
| 34 | | IGKV1D-37 |
| 35 | | PECAM1 |
| 36 | | IGHV3OR16-12 |
| 37 | | IGHV1OR15-1 |
| 38 | | IGHV3OR15-7 |
| 39 | TATA-binding protein-associated factor 2N | TAF15 |
| 40 | | IGLV5-45 |
| 41 | Ig heavy chain V-III region 23 | IGHV3-23 |
| 42 | | IGHV3-53 |
| 43 | | |
| 44 | | IGKV1D-8 |
| 45 | Ester hydrolase C11orf54 | C11orf54 |
| 46 | Collagen alpha-2(I) chain | COL1A2 |
| 47 | Ig kappa chain V-I region WAT | |
| 48 | Roundabout homolog 1 | ROBO1 |
| 49 | Ig kappa chain C region | IGKC |
| 50 | UBX domain-containing protein 1 | UBXN1 |
| 51 | Soluble scavenger receptor cysteine-rich domain-containing protein SSC5D | SSC5D |
| 52 | | IGHV3-49 |
| 53 | Heterogeneous nuclear ribonucleoproteins A2/B1 | HNRNPA2B1 |
| 54 | Bifunctional ATP-dependent dihydroxyacetone kinase/FAD-AMP lyase (cyclizing) | TKFC |

**Supp. Table 5. All urinary proteins measured**

| Protein No. | Protein names | Gene names |
|---|---|---|
| 55 | Ig delta chain C region | IGHD |
| 56 | Chondroitin sulfate proteoglycan 5 | CSPG5 |
| 57 | Laminin subunit alpha-3 | LAMA3 |
| 58 | | OSCAR |
| 59 | Aromatic-L-amino-acid decarboxylase | DDC |
| 60 | | IGHG1 |
| 61 | Testis-expressed sequence 2 protein | TEX2 |
| 62 | Scavenger receptor class F member 2 | SCARF2 |
| 63 | CD177 antigen | CD177 |
| 64 | | IGKV1-27 |
| 65 | Clathrin heavy chain | CLTC |
| 66 | Ig gamma-3 chain C region | IGHG3 |
| 67 | | IGHV3-72 |
| 68 | Mucosal addressin cell adhesion molecule 1 | MADCAM1 |
| 69 | Immunoglobulin lambda-like polypeptide 5 | IGLL5 |
| 70 | Transmembrane protease serine 13 | TMPRSS13 |
| 71 | | IGHV3-64 |
| 72 | Mucin-like protein 1 | MUCL1 |
| 73 | Mucin-1 | MUC1 |
| 74 | Syntaxin-binding protein 2 | STXBP2 |
| 75 | | ALB |
| 76 | Tropomyosin alpha-3 chain | TPM3 |
| 77 | Gamma-adducin | ADD3 |
| 78 | Integrin beta | ITGB2 |
| 79 | Proteasomal ubiquitin receptor ADRM1 | ADRM1 |
| 80 | Cadherin-4 | CDH4 |
| 81 | IgGFc-binding protein | FCGBP |
| 82 | Folate receptor gamma | FOLR3 |
| 83 | | IGHG3 |
| 84 | Basal cell adhesion molecule | BCAM |
| 85 | Collagen alpha-1(V) chain | COL5A1 |
| 86 | Vacuolar protein sorting-associated protein VTA1 homolog | VTA1 |
| 87 | Ig gamma-1 chain C region | IGHG1 |
| 88 | | IGHG1 |
| 89 | Laminin subunit alpha-2 | LAMA2 |
| 90 | Ig mu chain C region | IGHM |
| 91 | Seizure 6-like protein 2 | SEZ6L2 |
| 92 | | IGKC |
| 93 | | IGLL5 |

**Supp. Table 5. All urinary proteins measured**

| Protein No. | Protein names | Gene names |
|---|---|---|
| 94 | UTP--glucose-1-phosphate uridylyltransferase | UGP2 |
| 95 | Salivary acidic proline-rich phosphoprotein 1/2 | PRH1 |
| 96 | Plakophilin-1 | PKP1 |
| 97 | Tumor protein D54 | TPD52L2 |
| 98 | Integral membrane protein DGCR2/IDD | DGCR2 |
| 99 | Myristoylated alanine-rich C-kinase substrate | MARCKS |
| 100 | | IGKV1D-33 |
| 101 | Receptor protein-tyrosine kinase | MERTK |
| 102 | Low affinity immunoglobulin gamma Fc region receptor III-B | FCGR3B |
| 103 | Colipase | CLPS |
| 104 | | IGKV3-11 |
| 105 | | IGHG1 |
| 106 | Hypoxia up-regulated protein 1 | HYOU1 |
| 107 | | IGHG1 |
| 108 | Natriuretic peptides A | NPPA |
| 109 | Pro-cathepsin H | CTSH |
| 110 | Collagen alpha-1(XV) chain | COL15A1 |
| 111 | | |
| 112 | Kinesin-like protein | CENPE |
| 113 | | IGKV2D-29 |
| 114 | Ig kappa chain V-II region Cum | IGKV2-40 |
| 115 | Collagen alpha-1(VI) chain | COL6A1 |
| 116 | Heterogeneous nuclear ribonucleoprotein M | HNRNPM |
| 117 | | IGKC |
| 118 | Growth hormone receptor | GHR |
| 119 | Ras-related protein Rab-18 | RAB18 |
| 120 | | IGHG1 |
| 121 | 15 kDa selenoprotein | SEP15 |
| 122 | Glutathione peroxidase | GPX3 |
| 123 | C-C motif chemokine 15 | CCL15 |
| 124 | Nebulin | NEB |
| 125 | Aggrecan core protein | ACAN |
| 126 | Ig kappa chain V-II region TEW | IGKC |
| 127 | Complement C1s subcomponent | C1S |
| 128 | AP-2 complex subunit beta | AP2B1 |
| 129 | Basigin | BSG |
| 130 | Carcinoembryonic antigen-related cell adhesion molecule 8 | CEACAM8 |
| 131 | Inter-alpha-trypsin inhibitor heavy chain H5 | ITIH5 |
| 132 | Serum amyloid A-4 protein | SAA2-SAA4 |

**Supp. Table 5. All urinary proteins measured**

| Protein No. | Protein names | Gene names |
|---|---|---|
| 133 | Mucin-5AC | MUC5AC |
| 134 | Stathmin | STMN1 |
| 135 | Major prion protein | PRNP |
| 136 | Flotillin-1 | FLOT1 |
| 137 | Neuroblastoma suppressor of tumorigenicity 1 | NBL1 |
| 138 | ADP-ribosyl cyclase/cyclic ADP-ribose hydrolase 2 | BST1 |
| 139 | Putative keratin-87 protein | KRT87P |
| 140 | IgLON family member 5 | IGLON5 |
| 141 | Leukocyte immunoglobulin-like receptor subfamily A member 5 | LILRA5 |
| 142 | Aminopeptidase B | RNPEP |
| 143 | N-acylglucosamine 2-epimerase | RENBP |
| 144 | Protein shisa-7 | SHISA7 |
| 145 | V-set and transmembrane domain-containing protein 2B | VSTM2B |
| 146 | Alpha-endosulfine | ENSA |
| 147 | Collagen alpha-6(VI) chain | COL6A6 |
| 148 | Matrilin-4 | MATN4 |
| 149 | Tetraspanin | CD9 |
| 150 | Inositol oxygenase | MIOX |
| 151 | BPI fold-containing family A member 1 | BPIFA1 |
| 152 | Trypsin-2 | PRSS2 |
| 153 | Insulin-like growth factor-binding protein 3 | IGFBP3 |
| 154 | Alpha-2-macroglobulin-like protein 1 | A2ML1 |
| 155 | Protein FAM3B | FAM3B |
| 156 | Small ubiquitin-related modifier 3 | SUMO3 |
| 157 | Leukocyte immunoglobulin-like receptor subfamily B member 1 | LILRB1 |
| 158 | Putative neutrophil cytosol factor 1C | NCF1C |
| 159 | G-protein coupled receptor family C group 5 member C | GPRC5C |
| 160 | Platelet-derived growth factor subunit B | PDGFB |
| 161 | Interleukin-2 receptor subunit beta | IL2RB |
| 162 | Seizure 6-like protein | SEZ6L |
| 163 | Apolipoprotein C-III | APOC3 |
| 164 | Sulfurtransferase | MPST |
| 165 | Signal peptide, CUB and EGF-like domain-containing protein 1 | SCUBE1 |
| 166 | Receptor protein-tyrosine kinase | EPHB2 |
| 167 | Astrocytic phosphoprotein PEA-15 | PEA15 |
| 168 | Latrophilin-2 | ADGRL2 |
| 169 | Lymphocyte function-associated antigen 3 | CD58 |
| 170 | Dyslexia-associated protein KIAA0319-like protein | KIAA0319L |
| 171 | Complement decay-accelerating factor | CD55 |

**Supp. Table 5. All urinary proteins measured**

| Protein No. | Protein names | Gene names |
|---|---|---|
| 172 | WASH complex subunit FAM21C | FAM21A |
| 173 | Protein FAM151A | FAM151A |
| 174 | Prosaposin | PSAP |
| 175 | Microfibrillar-associated protein 5 | MFAP5 |
| 176 | Myelin regulatory factor | MYRF |
| 177 | Sodium channel subunit beta-1 | SCN1B |
| 178 | Citrate synthase | CS |
| 179 | Gamma-glutamyltransferase 6 | GGT6 |
| 180 | Complement C1r subcomponent | C1R |
| 181 | Complement factor B | CFB |
| 182 | Tumor necrosis factor receptor superfamily member 1B | TNFR1B |
| 183 | Transcobalamin-2 | TCN2 |
| 184 | Echinoderm microtubule-associated protein-like 4 | EML4 |
| 185 | Epithelial cell adhesion molecule | EPCAM |
| 186 | GTP-binding nuclear protein Ran | RAN |
| 187 | Abl interactor 1 | ABI1 |
| 188 | Serum albumin | ALB |
| 189 | Sodium/nucleoside cotransporter 1 | SLC28A1 |
| 190 | Inter-alpha-trypsin inhibitor heavy chain H4 | ITIH4 |
| 191 | CapZ-interacting protein | RCSD1 |
| 192 | Metalloendopeptidase | MEP1A |
| 193 | Parvalbumin alpha | PVALB |
| 194 | 10 kDa heat shock protein, mitochondrial | HSPE1 |
| 195 | Interleukin-1 receptor type 1 | IL1R1 |
| 196 | Prothymosin alpha | PTMA |
| 197 | Butyrophilin subfamily 2 member A2 | BTN2A2 |
| 198 | Exostosin-like 2 | EXTL2 |
| 199 | Protein shisa-5 | SHISA5 |
| 200 | Mucin-13 | MUC13 |
| 201 | Type 2 lactosamine alpha-2,3-sialyltransferase | ST3GAL6 |
| 202 | Charged multivesicular body protein 2b | CHMP2B |
| 203 | Profilin | PFN2 |
| 204 | Sorcin | SRI |
| 205 | Protein FAM107B | FAM107B |
| 206 | Troponin C, skeletal muscle | TNNC2 |
| 207 | Immunoglobulin superfamily member 8 | IGSF8 |
| 208 | 40S ribosomal protein SA | RPSA |
| 209 | | TF |
| 210 | Immunoglobulin superfamily member 11 | IGSF11 |

**Supp. Table 5. All urinary proteins measured**

| Protein No. | Protein names | Gene names |
|---|---|---|
| 211 | Tissue factor pathway inhibitor | TFPI |
| 212 | Nuclear receptor-binding protein | NRBP1 |
| 213 | Lipoma-preferred partner | LPP |
| 214 | Voltage-dependent calcium channel subunit alpha-2/delta-2 | CACNA2D2 |
| 215 | Protein lifeguard 3 | TMBIM1 |
| 216 | C-X-C chemokine receptor type 2 | CXCR2 |
| 217 | Paired immunoglobulin-like type 2 receptor alpha | PILRA |
| 218 | Acylamino-acid-releasing enzyme | APEH |
| 219 | | TFPI |
| 220 | Transmembrane protease serine 2 | TMPRSS2 |
| 221 | V-type proton ATPase subunit B, kidney isoform | ATP6V1B1 |
| 222 | Myotrophin | MTPN |
| 223 | HEPACAM family member 2 | HEPACAM2 |
| 224 | Zinc transporter ZIP5 | SLC39A5 |
| 225 | Echinoderm microtubule-associated protein-like 2 | EML2 |
| 226 | Leukocyte immunoglobulin-like receptor subfamily B member 4 | LILRB4 |
| 227 | Putative transmembrane protein INAFM1 | INAFM1 |
| 228 | Insulin-like growth factor-binding protein 1 | IGFBP1 |
| 229 | Transmembrane protein 198 | TMEM198 |
| 230 | Insulin-like growth factor-binding protein 5 | IGFBP5 |
| 231 | Synaptophysin-like protein 1 | SYPL1 |
| 232 | Vacuolar protein sorting-associated protein 37D | VPS37D |
| 233 | Procollagen C-endopeptidase enhancer 2 | PCOLCE2 |
| 234 | Neuronal cell adhesion molecule | NRCAM |
| 235 | | TIGIT |
| 236 | SPARC | SPARC |
| 237 | Neuron-specific protein family member 1 | D4S234E |
| 238 | Target of Nesh-SH3 | ABI3BP |
| 239 | Glucosamine-6-phosphate isomerase | GNPDA1 |
| 240 | Complement C1q subcomponent subunit B | C1QB |
| 241 | Protein NOXP20 | FAM114A1 |
| 242 | Matrix extracellular phosphoglycoprotein | MEPE |
| 243 | PDZ and LIM domain protein 7 | PDLIM7 |
| 244 | Vesicular integral-membrane protein VIP36 | LMAN2 |
| 245 | Rap guanine nucleotide exchange factor 6 | RAPGEF6 |
| 246 | Alpha-L-iduronidase | IDUA |
| 247 | | GC |
| 248 | Cadherin-6 | CDH6 |
| 249 | Collagen alpha-1(XII) chain | COL12A1 |

**Supp. Table 5. All urinary proteins measured**

| Protein No. | Protein names | Gene names |
|---|---|---|
| 250 | Corticotropin-releasing factor-binding protein | CRHBP |
| 251 | Ribonuclease T2 | RNASET2 |
| 252 | Netrin receptor UNC5C | UNC5C |
| 253 | Proton-coupled amino acid transporter 2 | SLC36A2 |
| 254 | | PDGFRB |
| 255 | Copper transport protein ATOX1 | ATOX1 |
| 256 | Zinc transporter ZIP14 | SLC39A14 |
| 257 | Tubulin-specific chaperone A | TBCA |
| 258 | Syndecan | SDC2 |
| 259 | S-phase kinase-associated protein 1 | SKP1 |
| 260 | Latrophilin-3 | ADGRL3 |
| 261 | Vitamin K-dependent protein C | PROC |
| 262 | Protein-tyrosine-phosphatase | PTPRM |
| 263 | Tenascin-X | TNXB |
| 264 | Ezrin | EZR |
| 265 | Protein YIPF3 | YIPF3 |
| 266 | Maltase-glucoamylase, intestinal | MGAM |
| 267 | Neurexin-1 | NRXN1 |
| 268 | Calpastatin | CAST |
| 269 | Thrombospondin-4 | THBS4 |
| 270 | Calcitonin gene-related peptide 2 | CALCB |
| 271 | Urokinase-type plasminogen activator | PLAU |
| 272 | | |
| 273 | Lysosome-associated membrane glycoprotein 3 | LAMP3 |
| 274 | Folate receptor beta | FOLR2 |
| 275 | Casein kinase II subunit alpha 3 | CSNK2A1 |
| 276 | Sodium/calcium exchanger 1 | SLC8A1 |
| 277 | Latent-transforming growth factor beta-binding protein 1 | LTBP1 |
| 278 | Microtubule-associated protein | MAP4 |
| 279 | Hematopoietic lineage cell-specific protein | HCLS1 |
| 280 | Mucin-4 | MUC4 |
| 281 | Neuropilin-1 | NRP1 |
| 282 | Properdin | CFP |
| 283 | Alpha-mannosidase | MAN2B2 |
| 284 | Transporter | SLC6A19 |
| 285 | Catenin beta-1 | CTNNB1 |
| 286 | Epsilon-sarcoglycan | SGCE |
| 287 | Macrophage colony-stimulating factor 1 receptor | CSF1R |
| 288 | Receptor protein-tyrosine kinase | EPHA4 |

**Supp. Table 5. All urinary proteins measured**

| Protein No. | Protein names | Gene names |
|---|---|---|
| 289 | ATP-binding cassette sub-family A member 2 | ABCA2 |
| 290 | Mucin-20 | MUC20 |
| 291 | Tetranectin | CLEC3B |
| 292 | Pro-opiomelanocortin | POMC |
| 293 | Band 4.1-like protein 2 | EPB41L2 |
| 294 | Thy-1 membrane glycoprotein | THY1 |
| 295 | Mth938 domain-containing protein | AAMDC |
| 296 | Transmembrane protein 25 | TMEM25 |
| 297 | Mitogen-activated protein kinase | MAPK3 |
| 298 | Sodium channel subunit beta-3 | SCN3B |
| 299 | Ras-related protein R-Ras2 | RRAS2 |
| 300 | Junctional adhesion molecule-like | AMICA1 |
| 301 | Ras-related protein Rab-2A | RAB2A |
| 302 | ADM | ADM |
| 303 | Ras-related protein Rab-1B | RAB1B |
| 304 | Puromycin-sensitive aminopeptidase | NPEPPS |
| 305 | Tetraspanin | CD151 |
| 306 | Potassium voltage-gated channel subfamily E member 3 | KCNE3 |
| 307 | Brain-specific angiogenesis inhibitor 2 | ADGRB2 |
| 308 | Thioredoxin reductase 1, cytoplasmic | TXNRD1 |
| 309 | Nucleosome assembly protein 1-like 4 | NAP1L4 |
| 310 | CD59 glycoprotein | CD59 |
| 311 | Multivesicular body subunit 12A | MVB12A |
| 312 | Layilin | LAYN |
| 313 | Alpha-crystallin B chain | CRYAB |
| 314 | Serine protease 23 | PRSS23 |
| 315 | Cysteine and glycine-rich protein 1 | CSRP1 |
| 316 | Signal-regulatory protein beta-2 | SIRPB2 |
| 317 | Serine/threonine-protein kinase WNK1 | WNK1 |
| 318 | DNA damage-binding protein 1 | DDB1 |
| 319 | Maestro heat-like repeat-containing protein family member 2B | MROH2B |
| 320 | Amyloid-like protein 1 | APLP1 |
| 321 | 4F2 cell-surface antigen heavy chain | SLC3A2 |
| 322 | Scavenger receptor cysteine-rich type 1 protein M130 | CD163 |
| 323 | Malectin | MLEC |
| 324 | Ubiquitin-60S ribosomal protein L40 | UBC |
| 325 | Testican-2 | SPOCK2 |
| 326 | Tumor susceptibility gene 101 protein | TSG101 |
| 327 | Protein FAM198A | FAM198A |

**Supp. Table 5. All urinary proteins measured**

| Protein No. | Protein names | Gene names |
|---|---|---|
| 328 | Limbic system-associated membrane protein | LSAMP |
| 329 | Activin receptor type-1B | ACVR1B |
| 330 | Sodium/calcium exchanger 2 | SLC8A2 |
| 331 | Neutral alpha-glucosidase AB | GANAB |
| 332 | Oxidized low-density lipoprotein receptor 1 | OLR1 |
| 333 | Coagulation factor VII | F7 |
| 334 | Destrin | DSTN |
| 335 | Dickkopf-related protein 3 | DKK3 |
| 336 | Glyoxalase domain-containing protein 4 | GLOD4 |
| 337 | Tumor necrosis factor receptor superfamily member 14 | TNFRSF14 |
| 338 | Natural killer cells antigen CD94 | KLRD1 |
| 339 | Tetraspanin | CD63 |
| 340 | LIM domain and actin-binding protein 1 | LIMA1 |
| 341 | Cadherin-19 | CDH19 |
| 342 | Dynactin subunit 2 | DCTN2 |
| 343 | Vacuolar protein sorting-associated protein 29 | VPS29 |
| 344 | Keratin, type I cytoskeletal 18 | KRT18 |
| 345 | Protein canopy homolog 2 | CNPY2 |
| 346 | Matrix metalloproteinase-19 | MMP19 |
| 347 | 3(2),5-bisphosphate nucleotidase 1 | BPNT1 |
| 348 | Myosin light polypeptide 6 | MYL6 |
| 349 | Heterogeneous nuclear ribonucleoprotein A1 | HNRNPA1 |
| 350 | Protein sidekick-1 | SDK1 |
| 351 | Golgi integral membrane protein 4 | GOLIM4 |
| 352 | V-set and transmembrane domain-containing protein 2A | VSTM2A |
| 353 | Roundabout homolog 2 | ROBO2 |
| 354 | Disintegrin and metalloproteinase domain-containing protein 9 | ADAM9 |
| 355 | | EPHB6 |
| 356 | Endothelial cell-selective adhesion molecule | ESAM |
| 357 | Carboxylic ester hydrolase | BCHE |
| 358 | Elongation factor 1-beta | EEF1B2 |
| 359 | Transferrin receptor protein 1 | TFRC |
| 360 | Dipeptidyl peptidase 3 | DPP3 |
| 361 | Adenylate kinase 2, mitochondrial | AK2 |
| 362 | Ubiquitin-conjugating enzyme E2 variant 1 | UBE2V1 |
| 363 | V-type proton ATPase subunit D | ATP6V1D |
| 364 | Protein unc-79 homolog | UNC79 |
| 365 | Acylphosphatase | ACYP1 |
| 366 | Protein Z-dependent protease inhibitor | SERPINA10 |

**Supp. Table 5. All urinary proteins measured**

| Protein No. | Protein names | Gene names |
|---|---|---|
| 367 | | RNASE1 |
| 368 | | SERPINA3 |
| 369 | Proteasome subunit alpha type | PSMA6 |
| 370 | Latent-transforming growth factor beta-binding protein 2 | LTBP2 |
| 371 | | FBLN5 |
| 372 | Ectonucleoside triphosphate diphosphohydrolase 5 | ENTPD5 |
| 373 | Glia maturation factor beta | GMFB |
| 374 | Fibulin-5 | FBLN5 |
| 375 | Heterogeneous nuclear ribonucleoproteins C1/C2 | HNRNPC |
| 376 | Melanoma inhibitory activity protein 2 | MIA2 |
| 377 | Protein NDRG2 | NDRG2 |
| 378 | Cochlin | COCH |
| 379 | Laminin subunit beta-1 | LAMB1 |
| 380 | Hepatocyte growth factor-like protein | MST1 |
| 381 | Solute carrier family 12 member 2 | SLC12A2 |
| 382 | Complement factor I | CFI |
| 383 | Cartilage oligomeric matrix protein | COMP |
| 384 | Neurexin-2 | NRXN2 |
| 385 | Corneodesmosin | CDSN |
| 386 | Cadherin-23 | CDH23 |
| 387 | V-set and transmembrane domain-containing protein 1 | VSTM1 |
| 388 | Aldehyde dehydrogenase family 1 member A3 | ALDH1A3 |
| 389 | Zyxin | ZYX |
| 390 | Syntaxin-binding protein 5 | STXBP5 |
| 391 | Interleukin-10 receptor subunit beta | IL10RB |
| 392 | Serine/threonine-protein kinase SIK3 | SIK3 |
| 393 | CD99 antigen-like protein 2 | CD99L2 |
| 394 | Drebrin-like protein | DBNL |
| 395 | Glycodelin | PAEP |
| 396 | Low affinity immunoglobulin gamma Fc region receptor III-A | FCGR3A |
| 397 | Calmodulin | CALM2 |
| 398 | Cyclic AMP-dependent transcription factor ATF-6 beta | ATF6B |
| 399 | Amphiphysin | AMPH |
| 400 | 2-deoxynucleoside 5-phosphate N-hydrolase 1 | DNPH1 |
| 401 | ADP-ribosyl cyclase/cyclic ADP-ribose hydrolase 1 | CD38 |
| 402 | Adaptin ear-binding coat-associated protein 2 | NECAP2 |
| 403 | Thioredoxin domain-containing protein 15 | TXNDC15 |
| 404 | Interleukin-6 receptor subunit alpha | IL6R |
| 405 | Plasma kallikrein | KLKB1 |

**Supp. Table 5. All urinary proteins measured**

| Protein No. | Protein names | Gene names |
|---|---|---|
| 406 | Retinol-binding protein 1 | RBP1 |
| 407 | Nephronectin | NPNT |
| 408 | Latent-transforming growth factor beta-binding protein 3 | LTBP3 |
| 409 | Acetyltransferase component of pyruvate dehydrogenase complex | DLAT |
| 410 | ADP-sugar pyrophosphatase | NUDT5 |
| 411 | Cysteine-rich protein 2 | CRIP2 |
| 412 | Transmembrane 7 superfamily member 3 | TM7SF3 |
| 413 | alpha-1,2-Mannosidase | MAN1B1 |
| 414 | Receptor protein serine/threonine kinase | TGFBR1 |
| 415 | Serine/arginine-rich splicing factor 9 | SRSF9 |
| 416 | Beta-2-microglobulin | B2M |
| 417 | Proteasome subunit alpha type | PSMA4 |
| 418 | Acidic leucine-rich nuclear phosphoprotein 32 family member A | ANP32A |
| 419 | Signal peptide peptidase-like 2A | SPPL2A |
| 420 | Ras-related protein Rab-8B | RAB8B |
| 421 | TM2 domain-containing protein 3 | TM2D3 |
| 422 | Aflatoxin B1 aldehyde reductase member 2 | AKR7A2 |
| 423 | Probable ATP-dependent RNA helicase DDX17 | DDX17 |
| 424 | Cytochrome c oxidase subunit 5A, mitochondrial | COX5A |
| 425 | Protein CCSMST1 | C16orf91 |
| 426 | Hydroxyacylglutathione hydrolase, mitochondrial | HAGH |
| 427 | Dynein light chain roadblock-type 1 | DYNLRB2 |
| 428 | Interleukin-4 receptor subunit alpha | IL4R |
| 429 | Beta-hexosaminidase | HEXA |
| 430 | Cysteine-rich secretory protein LCCL domain-containing 2 | CRISPLD2 |
| 431 | Lysyl oxidase homolog 1 | LOXL1 |
| 432 | Ly6/PLAUR domain-containing protein 6B | LYPD6B |
| 433 | ATP-binding cassette sub-family B member 6, mitochondrial | ABCB6 |
| 434 | Neurofascin | NFASC |
| 435 | Protein-L-isoaspartate O-methyltransferase | PCMT1 |
| 436 | Proline-rich acidic protein 1 | PRAP1 |
| 437 | Platelet-derived growth factor subunit A | PDGFA |
| 438 | Zinc finger protein 185 | ZNF185 |
| 439 | Collagen alpha-1(XXVIII) chain | COL28A1 |
| 440 | Reticulon-4 receptor | RTN4R |
| 441 | Translin | TSN |
| 442 | CD99 antigen | CD99 |
| 443 | Grancalcin | GCA |
| 444 | Putative protein FAM10A4 | ST13 |

**Supp. Table 5. All urinary proteins measured**

| Protein No. | Protein names | Gene names |
|---|---|---|
| 445 | Phospholipid scramblase 1 | PLSCR1 |
| 446 | Neural cell adhesion molecule 2 | NCAM2 |
| 447 | Sex hormone-binding globulin | SHBG |
| 448 | Eukaryotic translation initiation factor 5A | EIF5A |
| 449 | Brain-specific angiogenesis inhibitor 1-associated protein 2 | BAIAP2 |
| 450 | Sarcalumenin | SRL |
| 451 | C-type lectin domain family 10 member A | CLEC10A |
| 452 | Fibroblast growth factor receptor | FGFR3 |
| 453 | Low-density lipoprotein receptor | LDLR |
| 454 | Cathelicidin antimicrobial peptide | CAMP |
| 455 | Semaphorin-4B | SEMA4B |
| 456 | Integrin alpha-7 | ITGA7 |
| 457 | Cysteine-rich secretory protein 3 | CRISP3 |
| 458 | Receptor protein-tyrosine kinase | FGFR4 |
| 459 | Fructose-bisphosphate aldolase | ALDOA |
| 460 | Trans-Golgi network integral membrane protein 2 | TGOLN2 |
| 461 | Rho GDP-dissociation inhibitor 1 | ARHGDIA |
| 462 | Cadherin-8 | CDH8 |
| 463 | L-xylulose reductase | DCXR |
| 464 | | SECTM1 |
| 465 | CMRF35-like molecule 1 | CD300LF |
| 466 | Galectin | LGALS9 |
| 467 | Uncharacterized protein C1orf159 | C1orf159 |
| 468 | Eukaryotic initiation factor 4A-I | EIF4A2 |
| 469 | Secernin-2 | SCRN2 |
| 470 | Flotillin-2 | FLOT2 |
| 471 | T-cell antigen CD7 | CD7 |
| 472 | Receptor protein-tyrosine kinase | ERBB2 |
| 473 | | SECTM1 |
| 474 | Intercellular adhesion molecule 2 | ICAM2 |
| 475 | Sodium channel protein | SCN4A |
| 476 | Fractalkine | CX3CL1 |
| 477 | Unconventional myosin-Id | MYO1D |
| 478 | Myosin regulatory light chain 12A | MYL12A |
| 479 | Lutropin subunit beta | LHB |
| 480 | Collagen alpha-1(XIV) chain | COL14A1 |
| 481 | Tryptase alpha/beta-1 | TPSAB1 |
| 482 | Low affinity immunoglobulin epsilon Fc receptor | FCER2 |
| 483 | ICOS ligand | ICOSLG |

**Supp. Table 5. All urinary proteins measured**

| Protein No. | Protein names | Gene names |
|---|---|---|
| 484 | WW domain-binding protein 2 | WBP2 |
| 485 | TAR DNA-binding protein 43 | TARDBP |
| 486 | Kin of IRRE-like protein 2 | KIRREL2 |
| 487 | Tubulin-folding cofactor B | TBCB |
| 488 | | PVRL2 |
| 489 | Angiopoietin-related protein 6 | ANGPTL6 |
| 490 | Envoplakin | EVPL |
| 491 | Periplakin | PPL |
| 492 | Glucosidase 2 subunit beta | PRKCSH |
| 493 | Histone H3 | H3F3B |
| 494 | Transmembrane protein 256 | TMEM256 |
| 495 | Cartilage intermediate layer protein 2 | CILP2 |
| 496 | Asialoglycoprotein receptor 1 | ASGR1 |
| 497 | | MXRA7 |
| 498 | Apolipoprotein C-II | APOC4-APOC2 |
| 499 | Complement factor D | CFD |
| 500 | Apolipoprotein C-I | APOC1 |
| 501 | Glutathione peroxidase | GPX4 |
| 502 | ATP synthase subunit alpha, mitochondrial | ATP5A1 |
| 503 | Follistatin-related protein 3 | FSTL3 |
| 504 | Calponin | CNN2 |
| 505 | Microfibril-associated glycoprotein 4 | MFAP4 |
| 506 | IGF-like family receptor 1 | IGFLR1 |
| 507 | Aquaporin-1 | AQP1 |
| 508 | Iron/zinc purple acid phosphatase-like protein | PAPL |
| 509 | | A1BG |
| 510 | CD209 antigen | CD209 |
| 511 | | C3 |
| 512 | ER membrane protein complex subunit 10 | EMC10 |
| 513 | Deoxyribonuclease-2-alpha | DNASE2 |
| 514 | PDZ and LIM domain protein 1 | PDLIM1 |
| 515 | Synaptosomal-associated protein 23 | SNAP23 |
| 516 | Mannan-binding lectin serine protease 2 | MASP2 |
| 517 | Signal-regulatory protein beta-1 | SIRPB1 |
| 518 | Membrane-associated progesterone receptor component 1 | PGRMC1 |
| 519 | Chloride intracellular channel protein 1 | CLIC1 |
| 520 | Uroplakin-1a | UPK1A |
| 521 | Prostaglandin E2 receptor EP3 subtype | PTGER3 |
| 522 | Sulfhydryl oxidase 1 | QSOX1 |

**Supp. Table 5. All urinary proteins measured**

| Protein No. | Protein names | Gene names |
|---|---|---|
| 523 | Beta-mannosidase | MANBA |
| 524 | Agrin | AGRN |
| 525 | Myc box-dependent-interacting protein 1 | BIN1 |
| 526 | Uroplakin-2 | UPK2 |
| 527 | Neural cell adhesion molecule L1-like protein | CHL1 |
| 528 | Delta-like protein 1 | DLL1 |
| 529 | Syntenin-1 | SDCBP |
| 530 | Syntenin-1 | SDCBP |
| 531 | Podocalyxin | PODXL |
| 532 | Lysosomal alpha-mannosidase | MAN2B1 |
| 533 | Fructose-1,6-bisphosphatase isozyme 2 | FBP2 |
| 534 | Pyridoxal kinase | PDXK |
| 535 | Immunoglobulin superfamily containing leucine-rich repeat protein | ISLR |
| 536 | Neurocan core protein | NCAN |
| 537 | Disintegrin and metalloproteinase domain-containing protein 10 | ADAM10 |
| 538 | Inositol monophosphatase 2 | IMPA2 |
| 539 | Na(+)/H(+) exchange regulatory cofactor NHE-RF1 | SLC9A3R1 |
| 540 | Tripeptidyl-peptidase 1 | TPP1 |
| 541 | Apolipoprotein L1 | APOL1 |
| 542 | Tumor necrosis factor receptor superfamily member 10C | TNFRSF10C |
| 543 | Proteasome subunit alpha type-7 | PSMA7 |
| 544 | Protocadherin-17 | PCDH17 |
| 545 | Plexin-B2 | PLXNB2 |
| 546 | A disintegrin and metalloproteinase with thrombospondin motifs 3 | ADAMTS3 |
| 547 | Actin-related protein 2/3 complex subunit 1B | ARPC1B |
| 548 | Actin-related protein 2/3 complex subunit 2 | ARPC2 |
| 549 | Actin-related protein 2/3 complex subunit 3 | ARPC3 |
| 550 | Ephrin type-B receptor 6 | EPHB6 |
| 551 | ADAM DEC1 | ADAMDEC1 |
| 552 | Laminin subunit alpha-5 | LAMA5 |
| 553 | Neurosecretory protein VGF | VGF |
| 554 | Synaptobrevin homolog YKT6 | YKT6 |
| 555 | Fatty acid-binding protein, brain | FABP7 |
| 556 | Leucine-rich repeat transmembrane protein FLRT2 | FLRT2 |
| 557 | Kunitz-type protease inhibitor 1 | SPINT1 |
| 558 | Trehalase | TREH |
| 559 | Kunitz-type protease inhibitor 2 | SPINT2 |
| 560 | Zinc finger protein 749 | ZNF749 |
| 561 | Thioredoxin-like protein 1 | TXNL1 |

**Supp. Table 5. All urinary proteins measured**

| Protein No. | Protein names | Gene names |
|---|---|---|
| 562 | Prominin-1 | PROM1 |
| 563 | Beta-1,4-glucuronyltransferase 1 | B4GAT1 |
| 564 | E3 ubiquitin-protein ligase RNF13 | RNF13 |
| 565 | Carbonic anhydrase 12 | CA12 |
| 566 | Charged multivesicular body protein 2a | CHMP2A |
| 567 | Prostate stem cell antigen | PSCA |
| 568 | Alpha-actinin-4 | ACTN4 |
| 569 | Calumenin | CALU |
| 570 | CD5 antigen-like | CD5L |
| 571 | Xaa-Pro aminopeptidase 2 | XPNPEP2 |
| 572 | Glia maturation factor gamma | GMFG |
| 573 | Transmembrane protease serine 11D | TMPRSS11D |
| 574 | Perilipin-1 | PLIN1 |
| 575 | Protocadherin-7 | PCDH7 |
| 576 | Guanine nucleotide-binding protein G(I)/G(S)/G(O) subunit gamma-7 | GNG7 |
| 577 | Sushi domain-containing protein 5 | SUSD5 |
| 578 | Lymphocyte antigen 75 | LY75 |
| 579 | Neuropilin-2 | NRP2 |
| 580 | Long-chain-fatty-acid--CoA ligase 4 | ACSL4 |
| 581 | Cubilin | CUBN |
| 582 | Beta-1,4-galactosyltransferase 3 | B4GALT3 |
| 583 | Toll-like receptor 2 | TLR2 |
| 584 | GDNF family receptor alpha-3 | GFRA3 |
| 585 | Tetraspanin-1 | TSPAN1 |
| 586 | Perilipin-3 | PLIN3 |
| 587 | Protein-tyrosine sulfotransferase 2 | TPST2 |
| 588 | Prosaposin receptor GPR37L1 | GPR37L1 |
| 589 | Protein CutA | CUTA |
| 590 | Sodium channel subunit beta-2 | SCN2B |
| 591 | EF-hand calcium-binding domain-containing protein 14 | EFCAB14 |
| 592 | WD repeat-containing protein 1 | WDR1 |
| 593 | Slit homolog 3 protein | SLIT3 |
| 594 | Copine-3 | CPNE3 |
| 595 | Gamma-glutamylcyclotransferase | GGCT |
| 596 | Cadherin-16 | CDH16 |
| 597 | Cartilage intermediate layer protein 1 | CILP |
| 598 | Programmed cell death protein 6 | PDCD6 |
| 599 | Vacuolar protein sorting-associated protein 4B | VPS4B |
| 600 | Core histone macro-H2A.1 | H2AFY |

**Supp. Table 5. All urinary proteins measured**

| Protein No. | Protein names | Gene names |
|---|---|---|
| 601 | SH3 domain-binding glutamic acid-rich-like protein | SH3BGRL |
| 602 | Filamin-B | FLNB |
| 603 | Serine/threonine-protein kinase/endoribonuclease IRE1 | ERN1 |
| 604 | Glypican-4 | GPC4 |
| 605 | Ceroid-lipofuscinosis neuronal protein 5 | CLN5 |
| 606 | Tumor necrosis factor receptor superfamily member 21 | TNFRSF21 |
| 607 | Splicing factor 3B subunit 1 | SF3B1 |
| 608 | Peptidoglycan recognition protein 1 | PGLYRP1 |
| 609 | Protein CREG1 | CREG1 |
| 610 | Ficolin-3 | FCN3 |
| 611 | UDP-GalNAc:beta-1,3-N-acetylgalactosaminyltransferase 1 | B3GALNT1 |
| 612 | Renin receptor | ATP6AP2 |
| 613 | Isocitrate dehydrogenase [NADP] cytoplasmic | IDH1 |
| 614 | Attractin | ATRN |
| 615 | Cytosolic 10-formyltetrahydrofolate dehydrogenase | ALDH1L1 |
| 616 | Dysferlin | DYSF |
| 617 | Gamma-butyrobetaine dioxygenase | BBOX1 |
| 618 | Carboxypeptidase D | CPD |
| 619 | WNT1-inducible-signaling pathway protein 2 | WISP2 |
| 620 | N(G),N(G)-dimethylarginine dimethylaminohydrolase 1 | DDAH1 |
| 621 | Galactosylgalactosylxylosylprotein 3-beta-glucuronosyltransferase 3 | B3GAT3 |
| 622 | Lymphocyte antigen 6H | LY6H |
| 623 | Latrophilin-1 | LPHN1 |
| 624 | Endonuclease domain-containing 1 protein | ENDOD1 |
| 625 | SLIT and NTRK-like protein 5 | SLITRK5 |
| 626 | Sciellin | SCEL |
| 627 | Reticulon-3 | RTN3 |
| 628 | Protocadherin-8 | PCDH8 |
| 629 | Ly6/PLAUR domain-containing protein 3 | LYPD3 |
| 630 | 6-phosphogluconolactonase | PGLS |
| 631 | WNT1-inducible-signaling pathway protein 1 | WISP1 |
| 632 | Papilin | PAPLN |
| 633 | Apolipoprotein M | APOM |
| 634 | A disintegrin and metalloproteinase with thrombospondin motifs 2 | ADAMTS2 |
| 635 | Pantetheinase | VNN1 |
| 636 | Neuronal pentraxin receptor | NPTXR |
| 637 | Serine/threonine-protein kinase OSR1 | OXSR1 |
| 638 | N(G),N(G)-dimethylarginine dimethylaminohydrolase 2 | DDAH2 |
| 639 | Formimidoyltransferase-cyclodeaminase | FTCD |

**Supp. Table 5. All urinary proteins measured**

| Protein No. | Protein names | Gene names |
|---|---|---|
| 640 | EGF-containing fibulin-like extracellular matrix protein 2 | EFEMP2 |
| 641 | CD160 antigen | CD160 |
| 642 | Reversion-inducing cysteine-rich protein with Kazal motifs | RECK |
| 643 | Interleukin-18-binding protein | IL18BP |
| 644 | Napsin-A | NAPSA |
| 645 | Molybdopterin synthase sulfur carrier subunit | MOCS2 |
| 646 | Cytochrome b5 | CYB5A |
| 647 | L-lactate dehydrogenase A chain | LDHA |
| 648 | Retinal dehydrogenase 1 | ALDH1A1 |
| 649 | Glutathione reductase, mitochondrial | GSR |
| 650 | Superoxide dismutase [Cu-Zn] | SOD1 |
| 651 | Ceruloplasmin | CP |
| 652 | Coagulation factor VIII | F8 |
| 653 | Purine nucleoside phosphorylase | PNP |
| 654 | Hypoxanthine-guanine phosphoribosyltransferase | HPRT1 |
| 655 | Epidermal growth factor receptor | EGFR |
| 656 | Phosphoglycerate kinase 1 | PGK1 |
| 657 | Prothrombin | F2 |
| 658 | Haptoglobin | HP |
| 659 | Haptoglobin-related protein | HPR |
| 660 | Coagulation factor IX | F9 |
| 661 | Coagulation factor X | F10 |
| 662 | Plasminogen | PLG |
| 663 | Coagulation factor XII | F12 |
| 664 | Carbonic anhydrase 1 | CA1 |
| 665 | Carbonic anhydrase 2 | CA2 |
| 666 | Argininosuccinate synthase | ASS1 |
| 667 | Pancreatic secretory trypsin inhibitor | SPINK1 |
| 668 | Antithrombin-III | SERPINC1 |
| 669 | Alpha-1-antitrypsin | SERPINA1 |
| 670 | Alpha-1-antichymotrypsin | SERPINA3 |
| 671 | Angiotensinogen | AGT |
| 672 | Alpha-2-macroglobulin | A2M |
| 673 | Complement C3 | C3 |
| 674 | Complement C5 | C5 |
| 675 | Cystatin-C | CST3 |
| 676 | Cystatin-S | CST4 |
| 677 | Cystatin-SN | CST1 |
| 678 | Cystatin-A | CSTA |

**Supp. Table 5. All urinary proteins measured**

| Protein No. | Protein names | Gene names |
|---|---|---|
| 679 | Kininogen-1 | KNG1 |
| 680 | Kininogen-1 | KNG1 |
| 681 | GTPase NRas | NRAS |
| 682 | Pro-epidermal growth factor | EGF |
| 683 | Proenkephalin-A | PENK |
| 684 | Follitropin subunit beta | FSHB |
| 685 | Calcitonin | CALCA |
| 686 | Insulin-like growth factor II | IGF2 |
| 687 | Immunoglobulin J chain | IGJ |
| 688 | Ig kappa chain V-I region AU | |
| 689 | Ig kappa chain V-I region CAR | |
| 690 | Ig kappa chain V-I region DEE | |
| 691 | Ig kappa chain V-I region EU | |
| 692 | Ig kappa chain V-I region HK101 | |
| 693 | Ig kappa chain V-I region HK102 | IGKV1-5 |
| 694 | Ig kappa chain V-I region Ka | |
| 695 | Ig kappa chain V-I region Kue | |
| 696 | Ig kappa chain V-I region Lay | |
| 697 | Ig kappa chain V-I region Rei | |
| 698 | Ig kappa chain V-I region Roy | |
| 699 | Ig kappa chain V-I region Scw | |
| 700 | Ig kappa chain V-I region WEA | |
| 701 | Ig kappa chain V-I region Wes | |
| 702 | Ig kappa chain V-I region Mev | |
| 703 | Ig kappa chain V-I region Ni | |
| 704 | Ig kappa chain V-II region FR | |
| 705 | Ig kappa chain V-II region MIL | |
| 706 | Ig kappa chain V-III region B6 | |
| 707 | Ig kappa chain V-III region SIE | |
| 708 | Ig kappa chain V-III region NG9 | |
| 709 | Ig kappa chain V-III region Ti | |
| 710 | Ig kappa chain V-III region POM | |
| 711 | Ig kappa chain V-IV region Len | |
| 712 | Ig lambda chain V-I region NEW | |
| 713 | Ig lambda chain V-II region TRO | |
| 714 | Ig lambda chain V-II region BUR | |
| 715 | Ig lambda chain V-III region SH | |
| 716 | Ig lambda chain V-IV region X | |
| 717 | Ig lambda chain V-IV region Hil | |

**Supp. Table 5. All urinary proteins measured**

| Protein No. | Protein names | Gene names |
| --- | --- | --- |
| 718 | Ig lambda chain V-V region DEL | |
| 719 | Ig lambda chain V-VI region AR | |
| 720 | Ig heavy chain V-I region HG3 | |
| 721 | Ig heavy chain V-III region WEA | |
| 722 | Ig heavy chain V-III region TIL | |
| 723 | Ig heavy chain V-III region BUT | |
| 724 | Ig heavy chain V-III region NIE | |
| 725 | Ig heavy chain V-III region KOL | |
| 726 | Ig heavy chain V-III region BUR | |
| 727 | Ig heavy chain V-III region LAY | |
| 728 | Ig heavy chain V-III region WAS | |
| 729 | Ig heavy chain V-III region ZAP | |
| 730 | Ig heavy chain V-III region JON | |
| 731 | Ig heavy chain V-III region GAL | |
| 732 | Ig heavy chain V-II region WAH | |
| 733 | Ig heavy chain V-II region NEWM | |
| 734 | Polymeric immunoglobulin receptor | PIGR |
| 735 | Ig epsilon chain C region | IGHE |
| 736 | Ig gamma-2 chain C region | IGHG2 |
| 737 | Ig gamma-4 chain C region | IGHG4 |
| 738 | Ig alpha-1 chain C region | IGHA1 |
| 739 | Ig alpha-2 chain C region | IGHA2 |
| 740 | Hemoglobin subunit delta | HBD |
| 741 | Myoglobin | MB |
| 742 | Collagen alpha-1(I) chain | COL1A1 |
| 743 | Collagen alpha-1(II) chain | COL2A1 |
| 744 | Collagen alpha-1(III) chain | COL3A1 |
| 745 | Collagen alpha-1(IV) chain | COL4A1 |
| 746 | Prelamin-A/C | LMNA |
| 747 | Apolipoprotein A-I | APOA1 |
| 748 | Apolipoprotein E | APOE |
| 749 | Fibrinogen alpha chain | FGA |
| 750 | Fibrinogen beta chain | FGB |
| 751 | Fibrinogen gamma chain | FGG |
| 752 | C-reactive protein | CRP |
| 753 | Serum amyloid P-component | APCS |
| 754 | Complement C1q subcomponent subunit C | C1QC |
| 755 | Complement component C9 | C9 |
| 756 | Beta-2-glycoprotein 1 | APOH |

**Supp. Table 5. All urinary proteins measured**

| Protein No. | Protein names | Gene names |
|---|---|---|
| 757 | Leucine-rich alpha-2-glycoprotein | LRG1 |
| 758 | Fibronectin | FN1 |
| 759 | Protein AMBP | AMBP |
| 760 | Alpha-1-acid glycoprotein 1 | ORM1 |
| 761 | Alpha-2-HS-glycoprotein | AHSG |
| 762 | Transthyretin | TTR |
| 763 | Vitamin D-binding protein | GC |
| 764 | Platelet basic protein | PPBP |
| 765 | Serotransferrin | TF |
| 766 | Lactotransferrin | LTF |
| 767 | Hemopexin | HPX |
| 768 | Ferritin light chain | FTL |
| 769 | Ferritin heavy chain | FTH1 |
| 770 | Metallothionein-2 | MT2A |
| 771 | Submaxillary gland androgen-regulated protein 3B | SMR3B |
| 772 | Angiogenin | ANG |
| 773 | Coagulation factor XI | F11 |
| 774 | Antileukoproteinase | SLPI |
| 775 | C4b-binding protein alpha chain | C4BPA |
| 776 | Vitronectin | VTN |
| 777 | Catalase | CAT |
| 778 | Glucosylceramidase | GBA |
| 779 | Tissue alpha-L-fucosidase | FUCA1 |
| 780 | Cystatin-B | CSTB |
| 781 | Annexin A1 | ANXA1 |
| 782 | Apolipoprotein B-100 | APOB |
| 783 | Trefoil factor 1 | TFF1 |
| 784 | Superoxide dismutase [Mn], mitochondrial | SOD2 |
| 785 | Phosphatidylcholine-sterol acyltransferase | LCAT |
| 786 | Histidine-rich glycoprotein | HRG |
| 787 | Ig kappa chain V-III region GOL | |
| 788 | Ig kappa chain V-III region CLL | |
| 789 | Ig lambda chain V-I region WAH | |
| 790 | Ig lambda chain V-II region NIG-84 | |
| 791 | Ig lambda chain V region 4A | |
| 792 | Alpha-1B-glycoprotein | A1BG |
| 793 | Alpha-1B-glycoprotein | A1BG |
| 794 | HLA class II histocompatibility antigen gamma chain | CD74 |
| 795 | von Willebrand factor | VWF |

**Supp. Table 5. All urinary proteins measured**

| Protein No. | Protein names | Gene names |
|---|---|---|
| 796 | Semenogelin-1 | SEMG1 |
| 797 | Glyceraldehyde-3-phosphate dehydrogenase | GAPDH |
| 798 | Argininosuccinate lyase | ASL |
| 799 | Ig kappa chain V-I region BAN | |
| 800 | Ig kappa chain V-I region Walker | |
| 801 | Alpha-amylase 1 | AMY1A |
| 802 | Pancreatic alpha-amylase | AMY2A |
| 803 | Heat shock protein beta-1 | HSPB1 |
| 804 | Guanine nucleotide-binding protein G(i) subunit alpha-2 | GNAI2 |
| 805 | Sodium/potassium-transporting ATPase subunit beta-1 | ATP1B1 |
| 806 | Secretogranin-1 | CHGB |
| 807 | Fructose-bisphosphate aldolase B | ALDOB |
| 808 | Amyloid beta A4 protein | APP |
| 809 | Arginase-1 | ARG1 |
| 810 | Apolipoprotein D | APOD |
| 811 | Aldehyde dehydrogenase, mitochondrial | ALDH2 |
| 812 | Protein S100-A8 | S100A8 |
| 813 | Plasminogen activator inhibitor 2 | SERPINB2 |
| 814 | Plasma serine protease inhibitor | SERPINA5 |
| 815 | Plasma protease C1 inhibitor | SERPING1 |
| 816 | Coagulation factor XIII B chain | F13B |
| 817 | Myeloperoxidase | MPO |
| 818 | Alkaline phosphatase, tissue-nonspecific isozyme | ALPL |
| 819 | Intercellular adhesion molecule 1 | ICAM1 |
| 820 | 60S acidic ribosomal protein P2 | RPLP2 |
| 821 | Neuroendocrine protein 7B2 | SCG5 |
| 822 | Lithostathine-1-alpha | REG1A |
| 823 | HLA class I histocompatibility antigen, A-24 alpha chain | HLA-A |
| 824 | Thyroxine-binding globulin | SERPINA7 |
| 825 | Heparin cofactor 2 | SERPIND1 |
| 826 | Integrin beta-1 | ITGB1 |
| 827 | Calbindin | CALB1 |
| 828 | Myosin light chain 1/3, skeletal muscle isoform | MYL1 |
| 829 | Serine protease hepsin | HPN |
| 830 | Collagen alpha-2(V) chain | COL5A2 |
| 831 | Alpha-galactosidase A | GLA |
| 832 | Ig kappa chain V-III region IARC/BL41 | |
| 833 | Ig kappa chain V-IV region | IGKV4-1 |
| 834 | Ig kappa chain V-IV region B17 | |

**Supp. Table 5. All urinary proteins measured**

| Protein No. | Protein names | Gene names |
|---|---|---|
| 835 | Ig lambda chain V-VI region WLT | |
| 836 | Ig heavy chain V-I region Mot | |
| 837 | Ig heavy chain V-II region ARH-77 | |
| 838 | Gelsolin | GSN |
| 839 | Gelsolin | GSN |
| 840 | ATP synthase subunit beta, mitochondrial | ATP5B |
| 841 | Complement C2 | C2 |
| 842 | Protein S100-A9 | S100A9 |
| 843 | Apolipoprotein A-IV | APOA4 |
| 844 | Carcinoembryonic antigen-related cell adhesion molecule 5 | CEACAM5 |
| 845 | Alpha-enolase | ENO1 |
| 846 | Glycogen phosphorylase, liver form | PYGL |
| 847 | Glucose-6-phosphate isomerase | GPI |
| 848 | Lipoprotein lipase | LPL |
| 849 | Kallikrein-1 | KLK1 |
| 850 | Ig lambda chain V-I region EPS | |
| 851 | Ig lambda chain V-IV region MOL | |
| 852 | Acyl-CoA-binding protein | DBI |
| 853 | Fatty acid-binding protein, liver | FABP1 |
| 854 | L-lactate dehydrogenase B chain | LDHB |
| 855 | Thrombomodulin | THBD |
| 856 | Vitamin K-dependent protein S | PROS1 |
| 857 | Protein disulfide-isomerase | P4HB |
| 858 | Prostate-specific antigen | KLK3 |
| 859 | Asialoglycoprotein receptor 2 | ASGR2 |
| 860 | Cathepsin D | CTSD |
| 861 | Annexin A2 | ANXA2 |
| 862 | Complement component C8 alpha chain | C8A |
| 863 | Complement component C8 beta chain | C8B |
| 864 | Platelet glycoprotein Ib alpha chain | GP1BA |
| 865 | Complement component C8 gamma chain | C8G |
| 866 | Calpain-1 catalytic subunit | CAPN1 |
| 867 | Carbonic anhydrase 3 | CA3 |
| 868 | Involucrin | IVL |
| 869 | Decorin | DCN |
| 870 | Beta-hexosaminidase subunit beta | HEXB |
| 871 | Cathepsin L1 | CTSL |
| 872 | Profilin-1 | PFN1 |
| 873 | Adenine phosphoribosyltransferase | APRT |

**Supp. Table 5. All urinary proteins measured**

| Protein No. | Protein names | Gene names |
|---|---|---|
| 874 | Cathepsin B | CTSB |
| 875 | Heat shock protein HSP 90-alpha | HSP90AA1 |
| 876 | Ribonuclease pancreatic | RNASE1 |
| 877 | Heat shock 70 kDa protein 1A | HSPA1A |
| 878 | Beta-microseminoprotein | MSMB |
| 879 | Annexin A6 | ANXA6 |
| 880 | Tumor necrosis factor receptor superfamily member 16 | NGFR |
| 881 | Tumor necrosis factor receptor superfamily member 16 | NGFR |
| 882 | Multidrug resistance protein 1 | ABCB1 |
| 883 | Corticosteroid-binding globulin | SERPINA6 |
| 884 | Beta-glucuronidase | GUSB |
| 885 | Heat shock protein HSP 90-beta | HSP90AB1 |
| 886 | Neutrophil elastase | ELANE |
| 887 | 72 kDa type IV collagenase | MMP2 |
| 888 | Stromelysin-1 | MMP3 |
| 889 | Extracellular superoxide dismutase [Cu-Zn] | SOD3 |
| 890 | Cathepsin G | CTSG |
| 891 | Neprilysin | MME |
| 892 | Matrix Gla protein | MGP |
| 893 | Apolipoprotein(a) | LPA |
| 894 | Monocyte differentiation antigen CD14 | CD14 |
| 895 | Collagen alpha-2(IV) chain | COL4A2 |
| 896 | Hepatocyte growth factor receptor | MET |
| 897 | Melanotransferrin | MFI2 |
| 898 | Complement factor H | CFH |
| 899 | Vimentin | VIM |
| 900 | Alpha-2-antiplasmin | SERPINF2 |
| 901 | Annexin A5 | ANXA5 |
| 902 | Chymotrypsin-like elastase family member 3A | CELA3A |
| 903 | Dopamine beta-hydroxylase | DBH |
| 904 | Glutathione S-transferase A2 | GSTA2 |
| 905 | Glutathione S-transferase P | GSTP1 |
| 906 | Cystatin-SA | CST2 |
| 907 | Matrilysin | MMP7 |
| 908 | Villin-1 | VIL1 |
| 909 | Galectin-1 | LGALS1 |
| 910 | Dihydropteridine reductase | QDPR |
| 911 | Fructose-1,6-bisphosphatase 1 | FBP1 |
| 912 | 2,3-cyclic-nucleotide 3-phosphodiesterase | CNP |

**Supp. Table 5. All urinary proteins measured**

| Protein No. | Protein names | Gene names |
|---|---|---|
| 913 | Macrophage colony-stimulating factor 1 | CSF1 |
| 914 | Platelet-derived growth factor receptor beta | PDGFRB |
| 915 | Gastric inhibitory polypeptide | GIP |
| 916 | Tumor-associated calcium signal transducer 2 | TACSTD2 |
| 917 | Intestinal-type alkaline phosphatase | ALPI |
| 918 | Ubiquitin carboxyl-terminal hydrolase isozyme L1 | UCHL1 |
| 919 | Furin | FURIN |
| 920 | Leukotriene A-4 hydrolase | LTA4H |
| 921 | Fructose-bisphosphate aldolase C | ALDOC |
| 922 | Complement C4-A | C4A |
| 923 | Complement C4-B | C4B |
| 924 | Protein ELFN1 | ELFN1 |
| 925 | Pepsin A-3 | PGA3 |
| 926 | Serum amyloid A-1 protein | SAA1 |
| 927 | Serum amyloid A-2 protein | SAA2 |
| 928 | Putative transmembrane protein INAFM2 | INAFM2 |
| 929 | Adrenodoxin, mitochondrial | FDX1 |
| 930 | Serglycin | SRGN |
| 931 | Non-secretory ribonuclease | RNASE2 |
| 932 | Lysosomal alpha-glucosidase | GAA |
| 933 | Histone H1.4 | HIST1H1E |
| 934 | Osteopontin | SPP1 |
| 935 | Osteopontin | SPP1 |
| 936 | Osteopontin | SPP1 |
| 937 | Osteopontin | SPP1 |
| 938 | Receptor-type tyrosine-protein phosphatase F | PTPRF |
| 939 | Thioredoxin | TXN |
| 940 | Complement component C7 | C7 |
| 941 | Chromogranin-A | CHGA |
| 942 | Mast/stem cell growth factor receptor Kit | KIT |
| 943 | S-formylglutathione hydrolase | ESD |
| 944 | 60 kDa heat shock protein, mitochondrial | HSPD1 |
| 945 | Clusterin | CLU |
| 946 | 78 kDa glucose-regulated protein | HSPA5 |
| 947 | Laminin subunit gamma-1 | LAMC1 |
| 948 | Lysosomal acid phosphatase | ACP2 |
| 949 | Heat shock cognate 71 kDa protein | HSPA8 |
| 950 | Integrin alpha-M | ITGAM |
| 951 | Mannose-binding protein C | MBL2 |

**Supp. Table 5. All urinary proteins measured**

| Protein No. | Protein names | Gene names |
|---|---|---|
| 952 | Ras-related protein Ral-A | RALA |
| 953 | Ras-related protein Ral-B | RALB |
| 954 | Lysosome-associated membrane glycoprotein 1 | LAMP1 |
| 955 | Fibroblast growth factor receptor 1 | FGFR1 |
| 956 | Cholesteryl ester transfer protein | CETP |
| 957 | Uteroglobin | SCGB1A1 |
| 958 | Cation-independent mannose-6-phosphate receptor | IGF2R |
| 959 | Alcohol dehydrogenase class-3 | ADH5 |
| 960 | Fatty acid-binding protein, intestinal | FABP2 |
| 961 | Collagen alpha-2(VI) chain | COL6A2 |
| 962 | Collagen alpha-2(VI) chain | COL6A2 |
| 963 | Collagen alpha-3(VI) chain | COL6A3 |
| 964 | Coagulation factor V | F5 |
| 965 | Nucleoprotein TPR | TPR |
| 966 | Prolactin-inducible protein | PIP |
| 967 | Creatine kinase B-type | CKB |
| 968 | Low affinity immunoglobulin gamma Fc region receptor II-a | FCGR2A |
| 969 | High affinity immunoglobulin epsilon receptor subunit alpha | FCER1A |
| 970 | Annexin A3 | ANXA3 |
| 971 | Eosinophil cationic protein | RNASE3 |
| 972 | Alpha-actinin-1 | ACTN1 |
| 973 | Angiotensin-converting enzyme | ACE |
| 974 | Cadherin-1 | CDH1 |
| 975 | Promotilin | MLN |
| 976 | Myosin-7 | MYH7 |
| 977 | Proto-oncogene tyrosine-protein kinase Src | SRC |
| 978 | Xaa-Pro dipeptidase | PEPD |
| 979 | Gamma-interferon-inducible lysosomal thiol reductase | IFI30 |
| 980 | Lysosome-associated membrane glycoprotein 2 | LAMP2 |
| 981 | Ribonuclease inhibitor | RNH1 |
| 982 | Secretogranin-2 | SCG2 |
| 983 | Neural cell adhesion molecule 1 | NCAM1 |
| 984 | Versican core protein | VCAN |
| 985 | Elongation factor 2 | EEF2 |
| 986 | Protein disulfide-isomerase A4 | PDIA4 |
| 987 | Complement component C6 | C6 |
| 988 | Carcinoembryonic antigen-related cell adhesion molecule 1 | CEACAM1 |
| 989 | Carcinoembryonic antigen-related cell adhesion molecule 1 | CEACAM1 |
| 990 | Bone marrow proteoglycan | PRG2 |

**Supp. Table 5. All urinary proteins measured**

| Protein No. | Protein names | Gene names |
|---|---|---|
| 991 | HLA class II histocompatibility antigen, DRB1-4 beta chain | HLA-DRB1 |
| 992 | Plastin-2 | LCP1 |
| 993 | Plastin-3 | PLS3 |
| 994 | Collagen alpha-2(XI) chain | COL11A2 |
| 995 | L-selectin | SELL |
| 996 | Macrophage migration inhibitory factor | MIF |
| 997 | Farnesyl pyrophosphate synthase | FDPS |
| 998 | Carboxypeptidase M | CPM |
| 999 | Nidogen-1 | NID1 |
| 1000 | Alcohol dehydrogenase [NADP(+)] | AKR1A1 |
| 1001 | Pyruvate kinase PKM | PKM |
| 1002 | Endoplasmin | HSP90B1 |
| 1003 | Matrix metalloproteinase-9 | MMP9 |
| 1004 | Carboxypeptidase A1 | CPA1 |
| 1005 | Carboxypeptidase B | CPB1 |
| 1006 | Fatty acid-binding protein, adipocyte | FABP4 |
| 1007 | Glutamine synthetase | GLUL |
| 1008 | Aldose reductase | AKR1B1 |
| 1009 | Aminopeptidase N | ANPEP |
| 1010 | Poliovirus receptor | PVR |
| 1011 | Ras-related C3 botulinum toxin substrate 2 | RAC2 |
| 1012 | Carboxypeptidase N catalytic chain | CPN1 |
| 1013 | Arylsulfatase A | ARSA |
| 1014 | Beta-1,4-galactosyltransferase 1 | B4GALT1 |
| 1015 | Prostatic acid phosphatase | ACPP |
| 1016 | Folate receptor alpha | FOLR1 |
| 1017 | 15-hydroxyprostaglandin dehydrogenase [NAD(+)] | HPGD |
| 1018 | Membrane cofactor protein | CD46 |
| 1019 | N-acetylglucosamine-6-sulfatase | GNS |
| 1020 | Immunoglobulin lambda-like polypeptide 1 | IGLL1 |
| 1021 | Arylsulfatase B | ARSB |
| 1022 | Beta-galactoside alpha-2,6-sialyltransferase 1 | ST6GAL1 |
| 1023 | Desmoplakin | DSP |
| 1024 | Metalloproteinase inhibitor 2 | TIMP2 |
| 1025 | Atrial natriuretic peptide receptor 1 | NPR1 |
| 1026 | CD44 antigen | CD44 |
| 1027 | Integrin beta-4 | ITGB4 |
| 1028 | Carbonyl reductase [NADPH] 1 | CBR1 |
| 1029 | Pancreatic triacylglycerol lipase | PNLIP |

**Supp. Table 5. All urinary proteins measured**

| Protein No. | Protein names | Gene names |
|---|---|---|
| 1030 | Platelet-derived growth factor receptor alpha | PDGFRA |
| 1031 | Beta-galactosidase | GLB1 |
| 1032 | Platelet endothelial cell adhesion molecule | PECAM1 |
| 1033 | Histone H1.5 | HIST1H1B |
| 1034 | Dipeptidase 1 | DPEP1 |
| 1035 | Carboxypeptidase E | CPE |
| 1036 | Fumarylacetoacetase | FAH |
| 1037 | Y-box-binding protein 3 | YBX3 |
| 1038 | Alpha-N-acetylgalactosaminidase | NAGA |
| 1039 | Heat shock 70 kDa protein 6 | HSPA6 |
| 1040 | Aspartate aminotransferase, cytoplasmic | GOT1 |
| 1041 | Interferon alpha/beta receptor 1 | IFNAR1 |
| 1042 | Bactericidal permeability-increasing protein | BPI |
| 1043 | Atrial natriuretic peptide receptor 3 | NPR3 |
| 1044 | Sphingomyelin phosphodiesterase | SMPD1 |
| 1045 | Calpain-2 catalytic subunit | CAPN2 |
| 1046 | Endoglin | ENG |
| 1047 | Ganglioside GM2 activator | GM2A |
| 1048 | Galectin-3 | LGALS3 |
| 1049 | Insulin-like growth factor-binding protein 2 | IGFBP2 |
| 1050 | Ig kappa chain V-III region HAH | |
| 1051 | Ig kappa chain V-III region HIC | |
| 1052 | Vinculin | VCL |
| 1053 | Lipopolysaccharide-binding protein | LBP |
| 1054 | Interleukin-1 receptor antagonist protein | IL1RN |
| 1055 | Phosphoglycerate mutase 1 | PGAM1 |
| 1056 | Syndecan-1 | SDC1 |
| 1057 | Peptidyl-glycine alpha-amidating monooxygenase | PAM |
| 1058 | Cadherin-2 | CDH2 |
| 1059 | Vascular cell adhesion protein 1 | VCAM1 |
| 1060 | Tumor necrosis factor receptor superfamily member 1A | TNFRSF1A |
| 1061 | Gamma-glutamyltranspeptidase 1 | GGT1 |
| 1062 | Alpha-1-acid glycoprotein 2 | ORM2 |
| 1063 | Amiloride-sensitive amine oxidase [copper-containing] | AOC1 |
| 1064 | Inter-alpha-trypsin inhibitor heavy chain H2 | ITIH2 |
| 1065 | Inter-alpha-trypsin inhibitor heavy chain H1 | ITIH1 |
| 1066 | Elafin | PI3 |
| 1067 | Alpha-amylase 2B | AMY2B |
| 1068 | Thymidine phosphorylase | TYMP |

**Supp. Table 5. All urinary proteins measured**

| Protein No. | Protein names | Gene names |
|---|---|---|
| 1069 | Annexin A7 | ANXA7 |
| 1070 | Myeloid cell surface antigen CD33 | CD33 |
| 1071 | Gastricsin | PGC |
| 1072 | Kallikrein-2 | KLK2 |
| 1073 | Azurocidin | AZU1 |
| 1074 | Ras-related protein Rab-6A | RAB6A |
| 1075 | Lamin-B1 | LMNB1 |
| 1076 | Pregnancy zone protein | PZP |
| 1077 | Mimecan | OGN |
| 1078 | Ephrin-A1 | EFNA1 |
| 1079 | C4b-binding protein beta chain | C4BPB |
| 1080 | Myelin-associated glycoprotein | MAG |
| 1081 | N(4)-(beta-N-acetylglucosaminyl)-L-asparaginase | AGA |
| 1082 | Glutathione S-transferase Mu 3 | GSTM3 |
| 1083 | V-type proton ATPase subunit B, brain isoform | ATP6V1B2 |
| 1084 | Filamin-A | FLNA |
| 1085 | Cytoplasmic aconitate hydratase | ACO1 |
| 1086 | Glycerol-3-phosphate dehydrogenase [NAD(+)], cytoplasmic | GPD1 |
| 1087 | Ephrin type-A receptor 1 | EPHA1 |
| 1088 | Fibroblast growth factor receptor 2 | FGFR2 |
| 1089 | Biglycan | BGN |
| 1090 | Oxysterol-binding protein 1 | OSBP |
| 1091 | Tenascin-X | TNXB |
| 1092 | Cadherin-3 | CDH3 |
| 1093 | Iduronate 2-sulfatase | IDS |
| 1094 | Ubiquitin-like modifier-activating enzyme 1 | UBA1 |
| 1095 | Cornifin-B | SPRR1B |
| 1096 | Insulin-like growth factor-binding protein 4 | IGFBP4 |
| 1097 | Solute carrier family 2, facilitated glucose transporter member 5 | SLC2A5 |
| 1098 | Carbonic anhydrase 4 | CA4 |
| 1099 | Carboxypeptidase N subunit 2 | CPN2 |
| 1100 | Vitamin K-dependent protein Z | PROZ |
| 1101 | Neutrophil collagenase | MMP8 |
| 1102 | Macrophage mannose receptor 1 | MRC1 |
| 1103 | Ig heavy chain V-I region V35 | |
| 1104 | Fibulin-1 | FBLN1 |
| 1105 | Fibulin-1 | FBLN1 |
| 1106 | Peptidyl-prolyl cis-trans isomerase B | PPIB |
| 1107 | Protein S100-A1 | S100A1 |

**Supp. Table 5. All urinary proteins measured**

| Protein No. | Protein names | Gene names |
|---|---|---|
| 1108 | Receptor-type tyrosine-protein phosphatase delta | PTPRD |
| 1109 | Receptor-type tyrosine-protein phosphatase gamma | PTPRG |
| 1110 | Receptor-type tyrosine-protein phosphatase zeta | PTPRZ1 |
| 1111 | Adenosylhomocysteinase | AHCY |
| 1112 | Cofilin-1 | CFL1 |
| 1113 | Myeloblastin | PRTN3 |
| 1114 | Endothelin B receptor | EDNRB |
| 1115 | Insulin-like growth factor-binding protein 6 | IGFBP6 |
| 1116 | Acetyl-CoA acetyltransferase, mitochondrial | ACAT1 |
| 1117 | Tenascin | TNC |
| 1118 | Deoxyribonuclease-1 | DNASE1 |
| 1119 | Proteinase-activated receptor 1 | F2R |
| 1120 | Zinc-alpha-2-glycoprotein | AZGP1 |
| 1121 | Cathepsin S | CTSS |
| 1122 | Proteasome subunit alpha type-1 | PSMA1 |
| 1123 | Proteasome subunit alpha type-3 | PSMA3 |
| 1124 | Protein S100-P | S100P |
| 1125 | Collagen alpha-3(V) chain | COL5A3 |
| 1126 | Tumor necrosis factor receptor superfamily member 5 | CD40 |
| 1127 | Moesin | MSN |
| 1128 | Alpha-1,3-mannosyl-glycoprotein 2-beta-N-acetylglucosaminyltransferase | MGAT1 |
| 1129 | High mobility group protein B2 | HMGB2 |
| 1130 | Elongation factor 1-gamma | EEF1G |
| 1131 | CD27 antigen | CD27 |
| 1132 | Ciliary neurotrophic factor receptor subunit alpha | CNTFR |
| 1133 | Erythrocyte band 7 integral membrane protein | STOM |
| 1134 | Serum paraoxonase/arylesterase 1 | PON1 |
| 1135 | 14-3-3 protein theta | YWHAQ |
| 1136 | Calmodulin-like protein 3 | CALML3 |
| 1137 | Dipeptidyl peptidase 4 | DPP4 |
| 1138 | Calreticulin | CALR |
| 1139 | Calnexin | CANX |
| 1140 | Interleukin-1 receptor type 2 | IL1R2 |
| 1141 | Proteasome subunit alpha type-5 | PSMA5 |
| 1142 | Proteasome subunit beta type-4 | PSMB4 |
| 1143 | Protein-lysine 6-oxidase | LOX |
| 1144 | Cystatin-D | CST5 |
| 1145 | Granulins | GRN |
| 1146 | Cytosol aminopeptidase | LAP3 |

**Supp. Table 5. All urinary proteins measured**

| Protein No. | Protein names | Gene names |
|---|---|---|
| 1147 | Hematopoietic progenitor cell antigen CD34 | CD34 |
| 1148 | Tumor necrosis factor receptor superfamily member 8 | TNFRSF8 |
| 1149 | Connective tissue growth factor | CTGF |
| 1150 | Ephrin type-A receptor 2 | EPHA2 |
| 1151 | Tyrosine-protein phosphatase non-receptor type 6 | PTPN6 |
| 1152 | Cellular retinoic acid-binding protein 2 | CRABP2 |
| 1153 | Transketolase | TKT |
| 1154 | Serpin B3 | SERPINB3 |
| 1155 | Kallistatin | SERPINA4 |
| 1156 | Guanine nucleotide-binding protein subunit alpha-11 | GNA11 |
| 1157 | Delta-1-pyrroline-5-carboxylate dehydrogenase, mitochondrial | ALDH4A1 |
| 1158 | Phenazine biosynthesis-like domain-containing protein | PBLD |
| 1159 | Peroxiredoxin-6 | PRDX6 |
| 1160 | Flavin reductase (NADPH) | BLVRB |
| 1161 | Peroxiredoxin-5, mitochondrial | PRDX5 |
| 1162 | D-dopachrome decarboxylase | DDT |
| 1163 | GTP cyclohydrolase 1 feedback regulatory protein | GCHFR |
| 1164 | Thioredoxin-dependent peroxide reductase, mitochondrial | PRDX3 |
| 1165 | Phosphatidylethanolamine-binding protein 1 | PEBP1 |
| 1166 | Protein disulfide-isomerase A3 | PDIA3 |
| 1167 | HLA class I histocompatibility antigen, A-34 alpha chain | HLA-A |
| 1168 | HLA class I histocompatibility antigen, B-42 alpha chain | HLA-B |
| 1169 | Tyrosine-protein kinase receptor UFO | AXL |
| 1170 | Sodium- and chloride-dependent GABA transporter 1 | SLC6A1 |
| 1171 | Leukocyte elastase inhibitor | SERPINB1 |
| 1172 | Lipocalin-1 | LCN1 |
| 1173 | Coronin-1A | CORO1A |
| 1174 | Rab GDP dissociation inhibitor alpha | GDI1 |
| 1175 | Protein S100-A7 | S100A7 |
| 1176 | Fibroblast growth factor 9 | FGF9 |
| 1177 | Syndecan-4 | SDC4 |
| 1178 | Caspase-14 | CASP14 |
| 1179 | 14-3-3 protein beta/alpha | YWHAB |
| 1180 | 14-3-3 protein sigma | SFN |
| 1181 | Protein S100-A11 | S100A11 |
| 1182 | Neural cell adhesion molecule L1 | L1CAM |
| 1183 | Peroxiredoxin-2 | PRDX2 |
| 1184 | 4-hydroxyphenylpyruvate dioxygenase | HPD |
| 1185 | Desmoglein-3 | DSG3 |

**Supp. Table 5. All urinary proteins measured**

| Protein No. | Protein names | Gene names |
|---|---|---|
| 1186 | Cadherin-5 | CDH5 |
| 1187 | Kinesin-1 heavy chain | KIF5B |
| 1188 | Mannosyl-oligosaccharide 1,2-alpha-mannosidase IA | MAN1A1 |
| 1189 | N-acetylgalactosamine-6-sulfatase | GALNS |
| 1190 | Ribonuclease 4 | RNASE4 |
| 1191 | Macrosialin | CD68 |
| 1192 | Serine hydroxymethyltransferase, cytosolic | SHMT1 |
| 1193 | Protein EVI2B | EVI2B |
| 1194 | Heat shock 70 kDa protein 4 | HSPA4 |
| 1195 | Glypican-1 | GPC1 |
| 1196 | Radixin | RDX |
| 1197 | Sepiapterin reductase | SPR |
| 1198 | Cornifin-A | SPRR1A |
| 1199 | Small proline-rich protein 2B | SPRR2B |
| 1200 | Small proline-rich protein 2A | SPRR2A |
| 1201 | Fibrillin-1 | FBN1 |
| 1202 | Phosphoenolpyruvate carboxykinase, cytosolic [GTP] | PCK1 |
| 1203 | Myosin-9 | MYH9 |
| 1204 | Tyrosine-protein kinase receptor Tie-1 | TIE1 |
| 1205 | Coatomer subunit beta | COPB2 |
| 1206 | Basigin | BSG |
| 1207 | Glutaredoxin-1 | GLRX |
| 1208 | Insulin-like growth factor-binding protein complex acid labile subunit | IGFALS |
| 1209 | Vascular endothelial growth factor receptor 2 | KDR |
| 1210 | 26S protease regulatory subunit 7 | PSMC2 |
| 1211 | Chitinase-3-like protein 1 | CHI3L1 |
| 1212 | V-type proton ATPase subunit E 1 | ATP6V1E1 |
| 1213 | 7,8-dihydro-8-oxoguanine triphosphatase | NUDT1 |
| 1214 | Phosphoglucomutase-1 | PGM1 |
| 1215 | Tumor necrosis factor receptor superfamily member 3 | LTBR |
| 1216 | Serpin B5 | SERPINB5 |
| 1217 | Pigment epithelium-derived factor | SERPINF1 |
| 1218 | Dihydrolipoyllysine-residue succinyltransferase component of 2-oxoglutarate dehydrogenase complex, mitochondrial | DLST |
| 1219 | Complement factor H-related protein 2 | CFHR2 |
| 1220 | TGF-beta receptor type-2 | TGFBR2 |
| 1221 | Transgelin-2 | TAGLN2 |
| 1222 | Transaldolase | TALDO1 |
| 1223 | V-type proton ATPase catalytic subunit A | ATP6V1A |
| 1224 | Collagen alpha-1(XVIII) chain | COL18A1 |

**Supp. Table 5. All urinary proteins measured**

| Protein No. | Protein names | Gene names |
|---|---|---|
| 1225 | Macrophage-capping protein | CAPG |
| 1226 | Interleukin-6 receptor subunit beta | IL6ST |
| 1227 | Carcinoembryonic antigen-related cell adhesion molecule 6 | CEACAM6 |
| 1228 | Malate dehydrogenase, cytoplasmic | MDH1 |
| 1229 | Malate dehydrogenase, mitochondrial | MDH2 |
| 1230 | Leptin | LEP |
| 1231 | Aquaporin-2 | AQP2 |
| 1232 | Myeloid cell nuclear differentiation antigen | MNDA |
| 1233 | Prostaglandin-H2 D-isomerase | PTGDS |
| 1234 | Aldo-keto reductase family 1 member C3 | AKR1C3 |
| 1235 | Tyrosine-protein kinase FRK | FRK |
| 1236 | Leukemia inhibitory factor receptor | LIFR |
| 1237 | Lysosomal Pro-X carboxypeptidase | PRCP |
| 1238 | Cell surface glycoprotein MUC18 | MCAM |
| 1239 | Biotinidase | BTD |
| 1240 | Nicotinamide phosphoribosyltransferase | NAMPT |
| 1241 | Killer cell immunoglobulin-like receptor 2DL1 | KIR2DL1 |
| 1242 | Afamin | AFM |
| 1243 | Peptidyl-prolyl cis-trans isomerase C | PPIC |
| 1244 | Adapter molecule crk | CRK |
| 1245 | Neurogenic locus notch homolog protein 1 | NOTCH1 |
| 1246 | Utrophin | UTRN |
| 1247 | Ras GTPase-activating-like protein IQGAP1 | IQGAP1 |
| 1248 | 3-hydroxyanthranilate 3,4-dioxygenase | HAAO |
| 1249 | F-actin-capping protein subunit beta | CAPZB |
| 1250 | Stromal cell-derived factor 1 | CXCL12 |
| 1251 | Lithostathine-1-beta | REG1B |
| 1252 | Interferon alpha/beta receptor 2 | IFNAR2 |
| 1253 | Serpin B4 | SERPINB4 |
| 1254 | Glutathione synthetase | GSS |
| 1255 | Mannan-binding lectin serine protease 1 | MASP1 |
| 1256 | Protein NOV homolog | NOV |
| 1257 | CD97 antigen | CD97 |
| 1258 | 4-trimethylaminobutyraldehyde dehydrogenase | ALDH9A1 |
| 1259 | Protein-glutamine gamma-glutamyltransferase 4 | TGM4 |
| 1260 | Alpha-aminoadipic semialdehyde dehydrogenase | ALDH7A1 |
| 1261 | Proteasome subunit beta type-3 | PSMB3 |
| 1262 | Proteasome subunit beta type-2 | PSMB2 |
| 1263 | Histidine triad nucleotide-binding protein 1 | HINT1 |

**Supp. Table 5. All urinary proteins measured**

| Protein No. | Protein names | Gene names |
|---|---|---|
| 1264 | Retinoic acid receptor responder protein 1 | RARRES1 |
| 1265 | Selenoprotein P | SEPP1 |
| 1266 | Ketohexokinase | KHK |
| 1267 | Retinol-binding protein 2 | RBP2 |
| 1268 | Histamine N-methyltransferase | HNMT |
| 1269 | Guanine nucleotide-binding protein G(q) subunit alpha | GNAQ |
| 1270 | Rab GDP dissociation inhibitor beta | GDI2 |
| 1271 | Vasodilator-stimulated phosphoprotein | VASP |
| 1272 | Dynamin-2 | DNM2 |
| 1273 | Palmitoyl-protein thioesterase 1 | PPT1 |
| 1274 | T-complex protein 1 subunit theta | CCT8 |
| 1275 | T-complex protein 1 subunit delta | CCT4 |
| 1276 | Annexin A11 | ANXA11 |
| 1277 | Ras-related protein Rab-5C | RAB5C |
| 1278 | Ras-related protein Rab-7a | RAB7A |
| 1279 | Gastrotropin | FABP6 |
| 1280 | Amiloride-sensitive sodium channel subunit gamma | SCNN1G |
| 1281 | Galactokinase | GALK1 |
| 1282 | Glypican-3 | GPC3 |
| 1283 | N-sulphoglucosamine sulphohydrolase | SGSH |
| 1284 | Hepatoma-derived growth factor | HDGF |
| 1285 | Lumican | LUM |
| 1286 | Prolargin | PRELP |
| 1287 | Heterogeneous nuclear ribonucleoprotein A3 | HNRNPA3 |
| 1288 | 6-phosphogluconate dehydrogenase, decarboxylating | PGD |
| 1289 | Rho GDP-dissociation inhibitor 2 | ARHGDIB |
| 1290 | Ribonuclease UK114 | HRSP12 |
| 1291 | Hexokinase-3 | HK3 |
| 1292 | Ephrin-B2 | EFNB2 |
| 1293 | Stanniocalcin-1 | STC1 |
| 1294 | F-actin-capping protein subunit alpha-1 | CAPZA1 |
| 1295 | Biliverdin reductase A | BLVRA |
| 1296 | Dipeptidyl peptidase 1 | CTSC |
| 1297 | IST1 homolog | IST1 |
| 1298 | Cysteine-rich secretory protein 1 | CRISP1 |
| 1299 | Voltage-dependent calcium channel subunit alpha-2/delta-1 | CACNA2D1 |
| 1300 | Heat shock-related 70 kDa protein 2 | HSPA2 |
| 1301 | Sodium/potassium-transporting ATPase subunit gamma | FXYD2 |
| 1302 | UV excision repair protein RAD23 homolog A | RAD23A |

**Supp. Table 5. All urinary proteins measured**

| Protein No. | Protein names | Gene names |
|---|---|---|
| 1303 | UV excision repair protein RAD23 homolog B | RAD23B |
| 1304 | Ephrin type-B receptor 3 | EPHB3 |
| 1305 | Arylsulfatase F | ARSF |
| 1306 | Alpha-N-acetylglucosaminidase | NAGLU |
| 1307 | Galactocerebrosidase | GALC |
| 1308 | Growth arrest-specific protein 1 | GAS1 |
| 1309 | Secreted Ly-6/uPAR-related protein 1 | SLURP1 |
| 1310 | Solute carrier family 12 member 3 | SLC12A3 |
| 1311 | Phospholipid transfer protein | PLTP |
| 1312 | Transitional endoplasmic reticulum ATPase | VCP |
| 1313 | Inhibin beta C chain | INHBC |
| 1314 | Pancreatic secretory granule membrane major glycoprotein GP2 | GP2 |
| 1315 | Laminin subunit beta-2 | LAMB2 |
| 1316 | Cadherin-11 | CDH11 |
| 1317 | Cadherin-13 | CDH13 |
| 1318 | Cadherin-15 | CDH15 |
| 1319 | BH3-interacting domain death agonist | BID |
| 1320 | GDNF family receptor alpha-1 | GFRA1 |
| 1321 | Eukaryotic translation initiation factor 6 | EIF6 |
| 1322 | Junctional adhesion molecule B | JAM2 |
| 1323 | Ras-related protein Rab-25 | RAB25 |
| 1324 | Neutrophil defensin 3 | DEFA3 |
| 1325 | Actin-related protein 2/3 complex subunit 4 | ARPC4 |
| 1326 | Beta-defensin 1 | DEFB1 |
| 1327 | Triosephosphate isomerase | TPI1 |
| 1328 | Keratin-associated protein 10-6 | KRTAP10-6 |
| 1329 | Actin, cytoplasmic 1 | ACTB |
| 1330 | Cell division control protein 42 homolog | CDC42 |
| 1331 | Ras-related protein Rab-5B | RAB5B |
| 1332 | Ras-related protein Rab-10 | RAB10 |
| 1333 | Ubiquitin-conjugating enzyme E2 N | UBE2N |
| 1334 | Ras-related protein Rab-14 | RAB14 |
| 1335 | Actin-related protein 3 | ACTR3 |
| 1336 | Actin-related protein 2 | ACTR2 |
| 1337 | Ras-related protein Rap-1b | RAP1B |
| 1338 | Pterin-4-alpha-carbinolamine dehydratase | PCBD1 |
| 1339 | Lysozyme C | LYZ |
| 1340 | Epididymal secretory protein E1 | NPC2 |
| 1341 | Ubiquitin-fold modifier 1 | UFM1 |

**Supp. Table 5. All urinary proteins measured**

| Protein No. | Protein names | Gene names |
|---|---|---|
| 1342 | Nuclear transport factor 2 | NUTF2 |
| 1343 | 14-3-3 protein gamma | YWHAG |
| 1344 | 14-3-3 protein epsilon | YWHAE |
| 1345 | Thymosin beta-4 | TMSB4X |
| 1346 | ADP-ribosylation factor 6 | ARF6 |
| 1347 | Histone H4 | HIST1H4A |
| 1348 | Ras-related protein Rap-1A | RAP1A |
| 1349 | Guanine nucleotide-binding protein G(I)/G(S)/G(T) subunit beta-2 | GNB2 |
| 1350 | Peptidyl-prolyl cis-trans isomerase A | PPIA |
| 1351 | Peptidyl-prolyl cis-trans isomerase FKBP1A | FKBP1A |
| 1352 | Growth factor receptor-bound protein 2 | GRB2 |
| 1353 | Ras-related C3 botulinum toxin substrate 1 | RAC1 |
| 1354 | Guanine nucleotide-binding protein G(s) subunit alpha isoforms short | GNAS |
| 1355 | 14-3-3 protein zeta/delta | YWHAZ |
| 1356 | Actin, cytoplasmic 2 | ACTG1 |
| 1357 | Thymosin beta-10 | TMSB10 |
| 1358 | Tropomyosin alpha-4 chain | TPM4 |
| 1359 | Actin, alpha skeletal muscle | ACTA1 |
| 1360 | Tubulin alpha-1B chain | TUBA1B |
| 1361 | Tubulin beta-4B chain | TUBB4B |
| 1362 | Hemoglobin subunit beta | HBB |
| 1363 | Hemoglobin subunit gamma-1 | HBG1 |
| 1364 | Hemoglobin subunit alpha | HBA1 |
| 1365 | Coxsackievirus and adenovirus receptor | CXADR |
| 1366 | Tyrosine-protein phosphatase non-receptor type substrate 1 | SIRPA |
| 1367 | Disintegrin and metalloproteinase domain-containing protein 8 | ADAM8 |
| 1368 | Glutathione S-transferase omega-1 | GSTO1 |
| 1369 | Protein jagged-1 | JAG1 |
| 1370 | Interleukin-13 receptor subunit alpha-1 | IL13RA1 |
| 1371 | Phosphatidylinositol-glycan-specific phospholipase D | GPLD1 |
| 1372 | Neutrophil gelatinase-associated lipocalin | LCN2 |
| 1373 | Metallothionein-1H | MT1H |
| 1374 | Protein delta homolog 1 | DLK1 |
| 1375 | Ig heavy chain V-III region GAR | |
| 1376 | Protein S100-A12 | S100A12 |
| 1377 | Brain acid soluble protein 1 | BASP1 |
| 1378 | Ig lambda chain V-III region LOI | |
| 1379 | Hepcidin | HAMP |
| 1380 | Dermcidin | DCD |

**Supp. Table 5. All urinary proteins measured**

| Protein No. | Protein names | Gene names |
|---|---|---|
| 1381 | Retinol-binding protein 5 | RBP5 |
| 1382 | ADAMTS-like protein 3 | ADAMTSL3 |
| 1383 | Serine protease HTRA3 | HTRA3 |
| 1384 | Ig kappa chain V-IV region STH | |
| 1385 | ADP-ribosylation factor 1 | ARF1 |
| 1386 | Fibulin-2 | FBLN2 |
| 1387 | Basement membrane-specific heparan sulfate proteoglycan core protein | HSPG2 |
| 1388 | Polycystin-1 | PKD1 |
| 1389 | Low-density lipoprotein receptor-related protein 2 | LRP2 |
| 1390 | Ephrin-B1 | EFNB1 |
| 1391 | Sorbitol dehydrogenase | SORD |
| 1392 | Heterogeneous nuclear ribonucleoprotein U | HNRNPU |
| 1393 | CD83 antigen | CD83 |
| 1394 | Di-N-acetylchitobiase | CTBS |
| 1395 | Fatty acid-binding protein, epidermal | FABP5 |
| 1396 | Adenylyl cyclase-associated protein 1 | CAP1 |
| 1397 | Interferon-induced transmembrane protein 3 | IFITM3 |
| 1398 | Tyrosine-protein kinase transmembrane receptor ROR1 | ROR1 |
| 1399 | Tyrosine-protein kinase transmembrane receptor ROR2 | ROR2 |
| 1400 | Transgelin | TAGLN |
| 1401 | N-acylethanolamine-hydrolyzing acid amidase | NAAA |
| 1402 | Semenogelin-2 | SEMG2 |
| 1403 | Collagen alpha-1(VII) chain | COL7A1 |
| 1404 | Desmoglein-1 | DSG1 |
| 1405 | Desmocollin-2 | DSC2 |
| 1406 | Guanylin | GUCA2A |
| 1407 | Nucleobindin-1 | NUCB1 |
| 1408 | A-kinase anchor protein 12 | AKAP12 |
| 1409 | Complement factor H-related protein 3 | CFHR3 |
| 1410 | Aminoacylase-1 | ACY1 |
| 1411 | Transforming growth factor beta receptor type 3 | TGFBR3 |
| 1412 | Trefoil factor 2 | TFF2 |
| 1413 | Urokinase plasminogen activator surface receptor | PLAUR |
| 1414 | Complement factor H-related protein 1 | CFHR1 |
| 1415 | Glutamate carboxypeptidase 2 | FOLH1 |
| 1416 | Neurogenic locus notch homolog protein 2 | NOTCH2 |
| 1417 | Hepatocyte growth factor activator | HGFAC |
| 1418 | Lactoylglutathione lyase | GLO1 |
| 1419 | Activin receptor type-1 | ACVR1 |

**Supp. Table 5. All urinary proteins measured**

| Protein No. | Protein names | Gene names |
|---|---|---|
| 1420 | 14-3-3 protein eta | YWHAH |
| 1421 | Galectin-10 | CLC |
| 1422 | Inter-alpha-trypsin inhibitor heavy chain H3 | ITIH3 |
| 1423 | Regenerating islet-derived protein 3-alpha | REG3A |
| 1424 | Proteasome activator complex subunit 1 | PSME1 |
| 1425 | Tyrosine-protein kinase receptor TYRO3 | TYRO3 |
| 1426 | Amyloid-like protein 2 | APLP2 |
| 1427 | Fibromodulin | FMOD |
| 1428 | Peroxiredoxin-1 | PRDX1 |
| 1429 | Glutamyl aminopeptidase | ENPEP |
| 1430 | Dermatopontin | DPT |
| 1431 | Trefoil factor 3 | TFF3 |
| 1432 | Apoptosis regulator BAX | BAX |
| 1433 | Prolow-density lipoprotein receptor-related protein 1 | LRP1 |
| 1434 | Rho GTPase-activating protein 1 | ARHGAP1 |
| 1435 | Protocadherin-1 | PCDH1 |
| 1436 | Protein-glutamine gamma-glutamyltransferase E | TGM3 |
| 1437 | Quinone oxidoreductase | CRYZ |
| 1438 | Epithelial discoidin domain-containing receptor 1 | DDR1 |
| 1439 | Galectin-3-binding protein | LGALS3BP |
| 1440 | Desmocollin-1 | DSC1 |
| 1441 | Testican-1 | SPOCK1 |
| 1442 | Fibrinogen-like protein 1 | FGL1 |
| 1443 | Sialic acid-binding Ig-like lectin 14 | SIGLEC14 |
| 1444 | Neuroblast differentiation-associated protein AHNAK | AHNAK |
| 1445 | Apolipoprotein B receptor | APOBR |
| 1446 | Polypeptide N-acetylgalactosaminyltransferase 1 | GALNT1 |
| 1447 | Bone marrow stromal antigen 2 | BST2 |
| 1448 | Hyaluronidase-1 | HYAL1 |
| 1449 | EGF-containing fibulin-like extracellular matrix protein 1 | EFEMP1 |
| 1450 | EGF-containing fibulin-like extracellular matrix protein 1 | EFEMP1 |
| 1451 | Follistatin-related protein 1 | FSTL1 |
| 1452 | Contactin-1 | CNTN1 |
| 1453 | Cadherin-17 | CDH17 |
| 1454 | Receptor-type tyrosine-protein phosphatase eta | PTPRJ |
| 1455 | Tyrosine-protein phosphatase non-receptor type 13 | PTPN13 |
| 1456 | Epidermal growth factor receptor kinase substrate 8 | EPS8 |
| 1457 | Transcription initiation factor TFIID subunit 10 | TAF10 |
| 1458 | T-lymphoma invasion and metastasis-inducing protein 1 | TIAM1 |

**Supp. Table 5. All urinary proteins measured**

| Protein No. | Protein names | Gene names |
|---|---|---|
| 1459 | Secretory phospholipase A2 receptor | PLA2R1 |
| 1460 | Secreted phosphoprotein 24 | SPP2 |
| 1461 | PDZK1-interacting protein 1 | PDZK1IP1 |
| 1462 | BMP and activin membrane-bound inhibitor homolog | BAMBI |
| 1463 | Multimerin-1 | MMRN1 |
| 1464 | Selenium-binding protein 1 | SELENBP1 |
| 1465 | Chitotriosidase-1 | CHIT1 |
| 1466 | Inactive tyrosine-protein kinase 7 | PTK7 |
| 1467 | Receptor-type tyrosine-protein phosphatase S | PTPRS |
| 1468 | Mesothelin | MSLN |
| 1469 | Insulin-like growth factor I | IGF-I |
| 1470 | Disintegrin and metalloproteinase domain-containing protein 15 | ADAM15 |
| 1471 | Transmembrane emp24 domain-containing protein 1 | TMED1 |
| 1472 | Sequestosome-1 | SQSTM1 |
| 1473 | Ecto-ADP-ribosyltransferase 3 | ART3 |
| 1474 | Acid ceramidase | ASAH1 |
| 1475 | Polycystin-2 | PKD2 |
| 1476 | Semaphorin-5A | SEMA5A |
| 1477 | Solute carrier family 12 member 1 | SLC12A1 |
| 1478 | Ras-related protein Rab-32 | RAB32 |
| 1479 | CD166 antigen | ALCAM |
| 1480 | Apolipoprotein F | APOF |
| 1481 | Spectrin alpha chain, non-erythrocytic 1 | SPTAN1 |
| 1482 | Bone morphogenetic protein receptor type-2 | BMPR2 |
| 1483 | Carcinoembryonic antigen-related cell adhesion molecule 7 | CEACAM7 |
| 1484 | Coactosin-like protein | COTL1 |
| 1485 | Lysosome membrane protein 2 | SCARB2 |
| 1486 | Nidogen-2 | NID2 |
| 1487 | Dystroglycan | DAG1 |
| 1488 | Desmoglein-2 | DSG2 |
| 1489 | Tripartite motif-containing protein 29 | TRIM29 |
| 1490 | Dihydropyrimidinase-related protein 3 | DPYSL3 |
| 1491 | Lymphocyte antigen 6D | LY6D |
| 1492 | Src substrate cortactin | CTTN |
| 1493 | Fibroleukin | FGL2 |
| 1494 | Filamin-C | FLNC |
| 1495 | Frizzled-2 | FZD2 |
| 1496 | Guanine nucleotide-binding protein subunit alpha-13 | GNA13 |
| 1497 | Growth arrest-specific protein 6 | GAS6 |

**Supp. Table 5. All urinary proteins measured**

| Protein No. | Protein names | Gene names |
|---|---|---|
| 1498 | cGMP-inhibited 3,5-cyclic phosphodiesterase A | PDE3A |
| 1499 | WAP four-disulfide core domain protein 2 | WFDC2 |
| 1500 | SPARC-like protein 1 | SPARCL1 |
| 1501 | Hyaluronan-binding protein 2 | HABP2 |
| 1502 | Desmocollin-3 | DSC3 |
| 1503 | Plastin-1 | PLS1 |
| 1504 | Major vault protein | MVP |
| 1505 | LIM and SH3 domain protein 1 | LASP1 |
| 1506 | Ketimine reductase mu-crystallin | CRYM |
| 1507 | Prostaglandin reductase 1 | PTGR1 |
| 1508 | Transmembrane glycoprotein NMB | GPNMB |
| 1509 | Opioid-binding protein/cell adhesion molecule | OPCML |
| 1510 | Calcium-activated chloride channel regulator 4 | CLCA4 |
| 1511 | C-type lectin domain family 5 member A | CLEC5A |
| 1512 | Early endosome antigen 1 | EEA1 |
| 1513 | Protein disulfide-isomerase A6 | PDIA6 |
| 1514 | Procollagen C-endopeptidase enhancer 1 | PCOLCE |
| 1515 | Plectin | PLEC |
| 1516 | Inorganic pyrophosphatase | PPA1 |
| 1517 | Nectin-1 | PVRL1 |
| 1518 | Nicotinate-nucleotide pyrophosphorylase [carboxylating] | QPRT |
| 1519 | Reticulocalbin-1 | RCN1 |
| 1520 | Receptor tyrosine-protein kinase erbB-4 | ERBB4 |
| 1521 | Poly(rC)-binding protein 1 | PCBP1 |
| 1522 | Ephrin type-A receptor 7 | EPHA7 |
| 1523 | Ficolin-2 | FCN2 |
| 1524 | Transforming growth factor-beta-induced protein ig-h3 | TGFBI |
| 1525 | Na(+)/H(+) exchange regulatory cofactor NHE-RF2 | SLC9A3R2 |
| 1526 | Myosin light chain kinase, smooth muscle | MYLK |
| 1527 | Probable E3 ubiquitin-protein ligase HERC1 | HERC1 |
| 1528 | Cystatin-M | CST6 |
| 1529 | Adipogenesis regulatory factor | ADIRF |
| 1530 | Adiponectin | ADIPOQ |
| 1531 | V-type proton ATPase subunit S1 | ATP6AP1 |
| 1532 | Ras-related protein Rab-11B | RAB11B |
| 1533 | Insulin-like growth factor-binding protein 7 | IGFBP7 |
| 1534 | Laminin subunit alpha-4 | LAMA4 |
| 1535 | C3a anaphylatoxin chemotactic receptor | C3AR1 |
| 1536 | Extracellular matrix protein 1 | ECM1 |

**Supp. Table 5. All urinary proteins measured**

| Protein No. | Protein names | Gene names |
|---|---|---|
| 1537 | C-C motif chemokine 14 | CCL14 |
| 1538 | Drebrin | DBN1 |
| 1539 | Prostasin | PRSS8 |
| 1540 | Guanylate cyclase activator 2B | GUCA2B |
| 1541 | 2,4-dienoyl-CoA reductase, mitochondrial | DECR1 |
| 1542 | Alpha-mannosidase 2 | MAN2A1 |
| 1543 | Glutaminyl-peptide cyclotransferase | QPCT |
| 1544 | Discoidin domain-containing receptor 2 | DDR2 |
| 1545 | Receptor-type tyrosine-protein phosphatase-like N | PTPRN |
| 1546 | V-type proton ATPase subunit F | ATP6V1F |
| 1547 | Prostate-associated microseminoprotein | MSMP |
| 1548 | Transmembrane protein 132A | TMEM132A |
| 1549 | Inverted formin-2 | INF2 |
| 1550 | Fer-1-like protein 6 | FER1L6 |
| 1551 | HLA class II histocompatibility antigen, DRB1-8 beta chain | HLA-DRB1 |
| 1552 | Leucine-rich repeat flightless-interacting protein 1 | LRRFIP1 |
| 1553 | Nucleoside diphosphate kinase | NME1-NME2 |
| 1554 | Chromodomain-helicase-DNA-binding protein 9 | CHD9 |
| 1555 | CMRF35-like molecule 2 | CD300E |
| 1556 | Hydrocephalus-inducing protein homolog | HYDIN |
| 1557 | Cell adhesion molecule-related/down-regulated by oncogenes | CDON |
| 1558 | TBC1 domain family member 10B | TBC1D10B |
| 1559 | Choline transporter-like protein 4 | SLC44A4 |
| 1560 | Fibulin-7 | FBLN7 |
| 1561 | Cytochrome b reductase 1 | CYBRD1 |
| 1562 | Proline-rich transmembrane protein 3 | PRRT3 |
| 1563 | Metalloproteinase inhibitor 1 | TIMP1 |
| 1564 | Protein crumbs homolog 2 | CRB2 |
| 1565 | Rho-related GTP-binding protein RhoC | RHOC |
| 1566 | Melanoma inhibitory activity protein 3 | MIA3 |
| 1567 | NHL repeat-containing protein 3 | NHLRC3 |
| 1568 | Olfactomedin-like protein 2A | OLFML2A |
| 1569 | Serine/threonine-protein kinase 24 | STK24 |
| 1570 | Forkhead-associated domain-containing protein 1 | FHAD1 |
| 1571 | | SERPINB3 |
| 1572 | Collectin-12 | COLEC12 |
| 1573 | Sorting nexin-5 | SNX5 |
| 1574 | Proteasome inhibitor PI31 subunit | PSMF1 |
| 1575 | P-selectin | SELP |

**Supp. Table 5. All urinary proteins measured**

| Protein No. | Protein names | Gene names |
|---|---|---|
| 1576 | Tetratricopeptide repeat protein 38 | TTC38 |
| 1577 | Protein S100-A2 | S100A2 |
| 1578 | HLA class I histocompatibility antigen, alpha chain G | HLA-G |
| 1579 | Chloride intracellular channel protein 3 | CLIC3 |
| 1580 | Complement receptor type 1 | CR1 |
| 1581 | Calsyntenin-1 | CLSTN1 |
| 1582 | Allograft inflammatory factor 1 | AIF1 |
| 1583 | Protein FAM171A1 | FAM171A1 |
| 1584 | FRAS1-related extracellular matrix protein 2 | FREM2 |
| 1585 | D-3-phosphoglycerate dehydrogenase | PHGDH |
| 1586 | UMP-CMP kinase | CMPK1 |
| 1587 | Slit homolog 1 protein | SLIT1 |
| 1588 | SH3 domain-binding glutamic acid-rich-like protein 3 | SH3BGRL3 |
| 1589 | Nicastrin | NCSTN |
| 1590 | V-set domain-containing T-cell activation inhibitor 1 | VTCN1 |
| 1591 | Na(+)/H(+) exchange regulatory cofactor NHE-RF3 | PDZK1 |
| 1592 | Protein delta homolog 2 | DLK2 |
| 1593 | Tubulointerstitial nephritis antigen | TINAG |
| 1594 | Probable G-protein coupled receptor 110 | GPR110 |
| 1595 | Transmembrane protein 59 | TMEM59 |
| 1596 | Glutathione S-transferase Mu 5 | GSTM5 |
| 1597 | Ribosyldihydronicotinamide dehydrogenase [quinone] | NQO2 |
| 1598 | | CFH |
| 1599 | Signal-regulatory protein beta-1 isoform 3 | SIRPB1 |
| 1600 | Protein-tyrosine-phosphatase | PTPRK |
| 1601 | Protocadherin-9 | PCDH9 |
| 1602 | Putative elongation factor 1-alpha-like 3 | EEF1A1P5 |
| 1603 | BRO1 domain-containing protein BROX | BROX |
| 1604 | Retinol-binding protein 4 | RBP4 |
| 1605 | Platelet endothelial aggregation receptor 1 | PEAR1 |
| 1606 | Low-density lipoprotein receptor-related protein 11 | LRP11 |
| 1607 | Golgi-associated plant pathogenesis-related protein 1 | GLIPR2 |
| 1608 | Interleukin-2 receptor subunit alpha | IL2RA |
| 1609 | Translationally-controlled tumor protein | TPT1 |
| 1610 | CD276 antigen | CD276 |
| 1611 | Carboxypeptidase Z | CPZ |
| 1612 | Cyclic AMP-responsive element-binding protein 3-like protein 3 | CREB3L3 |
| 1613 | Natural cytotoxicity triggering receptor 3 ligand 1 | NCR3LG1 |
| 1614 | Transmembrane protein PVRIG | PVRIG |

**Supp. Table 5. All urinary proteins measured**

| Protein No. | Protein names | Gene names |
|---|---|---|
| 1615 | Vasorin | VASN |
| 1616 | Secreted frizzled-related protein 4 | SFRP4 |
| 1617 | Leukocyte-associated immunoglobulin-like receptor 1 | LAIR1 |
| 1618 | Twinfilin-2 | TWF2 |
| 1619 | Leukocyte-associated immunoglobulin-like receptor 2 | LAIR2 |
| 1620 | Uncharacterized protein C16orf46 | C16orf46 |
| 1621 | Annexin | ANXA4 |
| 1622 | Phospholipase B-like 1 | PLBD1 |
| 1623 | Protein CASC4 | CASC4 |
| 1624 | von Willebrand factor A domain-containing protein 1 | VWA1 |
| 1625 | 2-oxoglutarate and iron-dependent oxygenase domain-containing protein 3 | OGFOD3 |
| 1626 | Chondroitin sulfate proteoglycan 4 | CSPG4 |
| 1627 | Protein FAM198B | FAM198B |
| 1628 | Protein shisa-2 homolog | SHISA2 |
| 1629 | Suprabasin | SBSN |
| 1630 | Suprabasin | SBSN |
| 1631 | Ectonucleotide pyrophosphatase/phosphodiesterase family member 7 | ENPP7 |
| 1632 | Lipocalin-15 | LCN15 |
| 1633 | Olfactomedin-4 | OLFM4 |
| 1634 | Plexin domain-containing protein 2 | PLXDC2 |
| 1635 | UPF0764 protein C16orf89 | C16orf89 |
| 1636 | Ly6/PLAUR domain-containing protein 2 | LYPD2 |
| 1637 | C-type lectin domain family 4 member G | CLEC4G |
| 1638 | Peptidase inhibitor 16 | PI16 |
| 1639 | CMRF35-like molecule 9 | CD300LG |
| 1640 | Leucine-rich repeat neuronal protein 1 | LRRN1 |
| 1641 | C-type lectin domain family 9 member A | CLEC9A |
| 1642 | ADAMTS-like protein 4 | ADAMTSL4 |
| 1643 | Protocadherin Fat 4 | FAT4 |
| 1644 | Mucin-6 | MUC6 |
| 1645 | Nicotinate phosphoribosyltransferase | NAPRT |
| 1646 | Gliomedin | GLDN |
| 1647 | Scavenger receptor class A member 5 | SCARA5 |
| 1648 | Tropomyosin alpha-1 chain | TPM1 |
| 1649 | Ferric-chelate reductase 1 | FRRS1 |
| 1650 | Protein shisa-6 homolog | SHISA6 |
| 1651 | Hemojuvelin | HFE2 |
| 1652 | F-box only protein 50 | NCCRP1 |
| 1653 | Cyclic AMP-responsive element-binding protein 3-like protein 2 | CREB3L2 |

**Supp. Table 5. All urinary proteins measured**

| Protein No. | Protein names | Gene names |
|---|---|---|
| 1654 | Small integral membrane protein 5 | SMIM5 |
| 1655 | Isoaspartyl peptidase/L-asparaginase | ASRGL1 |
| 1656 | Glycerophosphodiester phosphodiesterase domain-containing protein 3 | GDPD3 |
| 1657 | Charged multivesicular body protein 1b | CHMP1B |
| 1658 | Neuronal growth regulator 1 | NEGR1 |
| 1659 | Low-density lipoprotein receptor-related protein 10 | LRP10 |
| 1660 | Growth/differentiation factor 7 | GDF7 |
| 1661 | UPF0669 protein C6orf120 | C6orf120 |
| 1662 | Vitelline membrane outer layer protein 1 homolog | VMO1 |
| 1663 | Podocan | PODN |
| 1664 | B- and T-lymphocyte attenuator | BTLA |
| 1665 | E3 ubiquitin-protein ligase HUWE1 | HUWE1 |
| 1666 | Transmembrane emp24 domain-containing protein 4 | TMED4 |
| 1667 | Multiple epidermal growth factor-like domains protein 8 | MEGF8 |
| 1668 | N-acetylgalactosaminyltransferase 7 | GALNT7 |
| 1669 | G-protein coupled receptor 126 | GPR126 |
| 1670 | C-type lectin domain family 14 member A | CLEC14A |
| 1671 | Uncharacterized protein C14orf37 | C14orf37 |
| 1672 | Serpin A11 | SERPINA11 |
| 1673 | Out at first protein homolog | OAF |
| 1674 | Reticulon-4 receptor-like 2 | RTN4RL2 |
| 1675 | Fermitin family homolog 3 | FERMT3 |
| 1676 | Thioredoxin domain-containing protein 5 | TXNDC5 |
| 1677 | Integral membrane protein GPR180 | GPR180 |
| 1678 | Cullin-associated NEDD8-dissociated protein 1 | CAND1 |
| 1679 | Transmembrane emp24 domain-containing protein 7 | TMED7 |
| 1680 | Xylosyltransferase 1 | XYLT1 |
| 1681 | Copine-8 | CPNE8 |
| 1682 | Plexin domain-containing protein 1 | PLXDC1 |
| 1683 | Adipocyte enhancer-binding protein 1 | AEBP1 |
| 1684 | Phospholipase D3 | PLD3 |
| 1685 | Protein AHNAK2 | AHNAK2 |
| 1686 | SLIT and NTRK-like protein 4 | SLITRK4 |
| 1687 | Choline transporter-like protein 2 | SLC44A2 |
| 1688 | MAX gene-associated protein | MGA |
| 1689 | Extracellular sulfatase Sulf-2 | SULF2 |
| 1690 | Ubiquitin-conjugating enzyme E2 variant 3 | UEVLD |
| 1691 | Transmembrane protein 192 | TMEM192 |
| 1692 | Osteoclast-associated immunoglobulin-like receptor | OSCAR |

**Supp. Table 5. All urinary proteins measured**

| Protein No. | Protein names | Gene names |
|---|---|---|
| 1693 | Probable G-protein coupled receptor 116 | GPR116 |
| 1694 | G-protein coupled receptor 64 | GPR64 |
| 1695 | Cell adhesion molecule 3 | CADM3 |
| 1696 | Prominin-2 | PROM2 |
| 1697 | Protein unc-80 homolog | UNC80 |
| 1698 | Ly6/PLAUR domain-containing protein 1 | LYPD1 |
| 1699 | Latent-transforming growth factor beta-binding protein 4 | LTBP4 |
| 1700 | Leucine-rich repeat-containing protein 25 | LRRC25 |
| 1701 | Maturin | MTURN |
| 1702 | Cell adhesion molecule 2 | CADM2 |
| 1703 | Secreted frizzled-related protein 1 | SFRP1 |
| 1704 | Leukocyte immunoglobulin-like receptor subfamily A member 3 | LILRA3 |
| 1705 | Discoidin, CUB and LCCL domain-containing protein 1 | DCBLD1 |
| 1706 | Golgi membrane protein 1 | GOLM1 |
| 1707 | E3 ubiquitin-protein ligase RNF149 | RNF149 |
| 1708 | Keratinocyte-associated transmembrane protein 2 | KCT2 |
| 1709 | Group XV phospholipase A2 | PLA2G15 |
| 1710 | NAD(P)H-hydrate epimerase | APOA1BP |
| 1711 | Hemicentin-2 | HMCN2 |
| 1712 | MAPK-interacting and spindle-stabilizing protein-like | MAPK1IP1L |
| 1713 | Beta-1,3-N-acetylglucosaminyltransferase lunatic fringe | LFNG |
| 1714 | DCC-interacting protein 13-beta | APPL2 |
| 1715 | Retinoic acid-induced protein 3 | GPRC5A |
| 1716 | Torsin-1A-interacting protein 2 | TOR1AIP2 |
| 1717 | Delta and Notch-like epidermal growth factor-related receptor | DNER |
| 1718 | Thiosulfate sulfurtransferase/rhodanese-like domain-containing protein 1 | TSTD1 |
| 1719 | Cell adhesion molecule 4 | CADM4 |
| 1720 | Putative phospholipase B-like 2 | PLBD2 |
| 1721 | Protocadherin-19 | PCDH19 |
| 1722 | T-cell immunomodulatory protein | ITFG1 |
| 1723 | Membrane protein FAM174A | FAM174A |
| 1724 | 5(3)-deoxyribonucleotidase, cytosolic type | NT5C |
| 1725 | BPI fold-containing family B member 1 | BPIFB1 |
| 1726 | Hepatitis A virus cellular receptor 2 | HAVCR2 |
| 1727 | Immunoglobulin superfamily DCC subclass member 4 | IGDCC4 |
| 1728 | Epidermal growth factor receptor kinase substrate 8-like protein 1 | EPS8L1 |
| 1729 | WASP homolog-associated protein with actin, membranes and microtubules | WHAMM |
| 1730 | Leucine-rich repeat-containing protein 15 | LRRC15 |
| 1731 | Marginal zone B- and B1-cell-specific protein | MZB1 |

**Supp. Table 5. All urinary proteins measured**

| Protein No. | Protein names | Gene names |
|---|---|---|
| 1732 | Programmed cell death 6-interacting protein | PDCD6IP |
| 1733 | Leucine-rich repeat neuronal protein 4 | LRRN4 |
| 1734 | Soluble calcium-activated nucleotidase 1 | CANT1 |
| 1735 | Intelectin-1 | ITLN1 |
| 1736 | Stabilin-2 | STAB2 |
| 1737 | High affinity immunoglobulin alpha and immunoglobulin mu Fc receptor | FCAMR |
| 1738 | Oncoprotein-induced transcript 3 protein | OIT3 |
| 1739 | Titin | TTN |
| 1740 | Roundabout homolog 4 | ROBO4 |
| 1741 | Acid sphingomyelinase-like phosphodiesterase 3a | SMPDL3A |
| 1742 | Acid sphingomyelinase-like phosphodiesterase 3b | SMPDL3B |
| 1743 | Complement factor H-related protein 4 | CFHR4 |
| 1744 | Protein FAM3C | FAM3C |
| 1745 | Protein NDRG1 | NDRG1 |
| 1746 | Peroxidasin homolog | PXDN |
| 1747 | Sortilin-related receptor | SORL1 |
| 1748 | Nectin-2 | PVRL2 |
| 1749 | Serine protease HTRA1 | HTRA1 |
| 1750 | Tenascin-R | TNR |
| 1751 | Secreted frizzled-related protein 3 | FRZB |
| 1752 | Gamma-glutamyl hydrolase | GGH |
| 1753 | Neogenin | NEO1 |
| 1754 | Kallikrein-6 | KLK6 |
| 1755 | Ubiquitin fusion degradation protein 1 homolog | UFD1L |
| 1756 | Golgi apparatus protein 1 | GLG1 |
| 1757 | Proteoglycan 4 | PRG4 |
| 1758 | Betaine--homocysteine S-methyltransferase 1 | BHMT |
| 1759 | Homogentisate 1,2-dioxygenase | HGD |
| 1760 | Liver-expressed antimicrobial peptide 2 | LEAP2 |
| 1761 | Myeloid-derived growth factor | MYDGF |
| 1762 | Tumor necrosis factor receptor superfamily member 19L | RELT |
| 1763 | Uncharacterized protein C11orf52 | C11orf52 |
| 1764 | Mas-related G-protein coupled receptor member F | MRGPRF |
| 1765 | Phosphotriesterase-related protein | PTER |
| 1766 | EF-hand domain-containing protein D2 | EFHD2 |
| 1767 | Aldose 1-epimerase | GALM |
| 1768 | Collagen triple helix repeat-containing protein 1 | CTHRC1 |
| 1769 | BTB/POZ domain-containing protein KCTD12 | KCTD12 |
| 1770 | Reticulocalbin-3 | RCN3 |

**Supp. Table 5. All urinary proteins measured**

| Protein No. | Protein names | Gene names |
|---|---|---|
| 1771 | Zymogen granule protein 16 homolog B | ZG16B |
| 1772 | Carboxymethylenebutenolidase homolog | CMBL |
| 1773 | Phosphoinositide-3-kinase-interacting protein 1 | PIK3IP1 |
| 1774 | Kinesin-like protein KIF12 | KIF12 |
| 1775 | Protein S100-A16 | S100A16 |
| 1776 | DPH3 homolog | DPH3 |
| 1777 | Phosphoglucomutase-2 | PGM2 |
| 1778 | Brevican core protein | BCAN |
| 1779 | PDZ and LIM domain protein 5 | PDLIM5 |
| 1780 | Cysteine-rich with EGF-like domain protein 1 | CRELD1 |
| 1781 | N-acyl-aromatic-L-amino acid amidohydrolase (carboxylate-forming) | ACY3 |
| 1782 | ERO1-like protein alpha | ERO1L |
| 1783 | Immunoglobulin superfamily member 21 | IGSF21 |
| 1784 | V-set and immunoglobulin domain-containing protein 2 | VSIG2 |
| 1785 | Alpha/beta hydrolase domain-containing protein 14B | ABHD14B |
| 1786 | Carboxypeptidase B2 | CPB2 |
| 1787 | Kin of IRRE-like protein 1 | KIRREL |
| 1788 | Beta-galactoside alpha-2,6-sialyltransferase 2 | ST6GAL2 |
| 1789 | Protocadherin-16 | DCHS1 |
| 1790 | Beta-Ala-His dipeptidase | CNDP1 |
| 1791 | Cytosolic non-specific dipeptidase | CNDP2 |
| 1792 | Receptor protein-tyrosine kinase | EPHB4 |
| 1793 | Fc receptor-like protein 1 | FCRL1 |
| 1794 | Intraflagellar transport protein 74 homolog | IFT74 |
| 1795 | Junctional sarcoplasmic reticulum protein 1 | JSRP1 |
| 1796 | Kremen protein 1 | KREMEN1 |
| 1797 | Nectin-4 | PVRL4 |
| 1798 | Serpin B12 | SERPINB12 |
| 1799 | N-acetylmuramoyl-L-alanine amidase | PGLYRP2 |
| 1800 | Erythroid membrane-associated protein | ERMAP |
| 1801 | SLIT and NTRK-like protein 1 | SLITRK1 |
| 1802 | Proton myo-inositol cotransporter | SLC2A13 |
| 1803 | Vacuolar protein sorting-associated protein 35 | VPS35 |
| 1804 | Vacuolar protein sorting-associated protein 13A | VPS13A |
| 1805 | Hemicentin-1 | HMCN1 |
| 1806 | Phosphatidylethanolamine-binding protein 4 | PEBP4 |
| 1807 | Probable carboxypeptidase X1 | CPXM1 |
| 1808 | Niban-like protein 1 | FAM129B |
| 1809 | Protein deglycase DJ-1 | PARK7 |

**Supp. Table 5. All urinary proteins measured**

| Protein No. | Protein names | Gene names |
|---|---|---|
| 1810 | Sialidase-1 | NEU1 |
| 1811 | Sortilin | SORT1 |
| 1812 | Synaptic vesicle membrane protein VAT-1 homolog | VAT1 |
| 1813 | Legumain | LGMN |
| 1814 | Neuroserpin | SERPINI1 |
| 1815 | Protein S100-A13 | S100A13 |
| 1816 | Oncostatin-M-specific receptor subunit beta | OSMR |
| 1817 | Cell growth regulator with EF hand domain protein 1 | CGREF1 |
| 1818 | Cell surface A33 antigen | GPA33 |
| 1819 | Smoothened homolog | SMO |
| 1820 | Histone H2A type 1-J | HIST1H2AJ |
| 1821 | Retinoic acid receptor responder protein 2 | RARRES2 |
| 1822 | Myocilin | MYOC |
| 1823 | Osteomodulin | OMD |
| 1824 | Growth/differentiation factor 15 | GDF15 |
| 1825 | Programmed cell death 1 ligand 2 | PDCD1LG2 |
| 1826 | Calsyntenin-3 | CLSTN3 |
| 1827 | Coronin-1B | CORO1B |
| 1828 | Thioredoxin domain-containing protein 17 | TXNDC17 |
| 1829 | Serine/threonine-protein phosphatase CPPED1 | CPPED1 |
| 1830 | Matrix-remodeling-associated protein 8 | MXRA8 |
| 1831 | 45 kDa calcium-binding protein | SDF4 |
| 1832 | Migration and invasion enhancer 1 | MIEN1 |
| 1833 | Endoplasmic reticulum resident protein 44 | ERP44 |
| 1834 | Protease-associated domain-containing protein 1 | PRADC1 |
| 1835 | Synaptotagmin-11 | SYT11 |
| 1836 | Ubiquitin-related modifier 1 | URM1 |
| 1837 | Plasma alpha-L-fucosidase | FUCA2 |
| 1838 | Chordin-like protein 1 | CHRDL1 |
| 1839 | Spondin-2 | SPON2 |
| 1840 | Protein crumbs homolog 3 | CRB3 |
| 1841 | Protein MENT | MENT |
| 1842 | 3-hydroxybutyrate dehydrogenase type 2 | BDH2 |
| 1843 | Vesicle-associated membrane protein 8 | VAMP8 |
| 1844 | Gamma-glutamylaminecyclotransferase | GGACT |
| 1845 | Specifically androgen-regulated gene protein | SARG |
| 1846 | Junctional adhesion molecule C | JAM3 |
| 1847 | TBC1 domain family member 10A | TBC1D10A |
| 1848 | C-type lectin domain family 7 member A | CLEC7A |

**Supp. Table 5. All urinary proteins measured**

| Protein No. | Protein names | Gene names |
|---|---|---|
| 1849 | Pappalysin-2 | PAPPA2 |
| 1850 | Complement factor H-related protein 5 | CFHR5 |
| 1851 | EMILIN-2 | EMILIN2 |
| 1852 | Charged multivesicular body protein 4a | CHMP4A |
| 1853 | Cell adhesion molecule 1 | CADM1 |
| 1854 | Cadherin-related family member 2 | CDHR2 |
| 1855 | Angiotensin-converting enzyme 2 | ACE2 |
| 1856 | Fibroblast growth factor-binding protein 2 | FGFBP2 |
| 1857 | Ly-6/neurotoxin-like protein 1 | LYNX1 |
| 1858 | Sialoadhesin | SIGLEC1 |
| 1859 | Tubulointerstitial nephritis antigen-like | TINAGL1 |
| 1860 | Twisted gastrulation protein homolog 1 | TWSG1 |
| 1861 | Toll-interacting protein | TOLLIP |
| 1862 | Protein ITFG3 | ITFG3 |
| 1863 | Cysteine-rich and transmembrane domain-containing protein 1 | CYSTM1 |
| 1864 | Multiple epidermal growth factor-like domains protein 9 | MEGF9 |
| 1865 | EH domain-containing protein 4 | EHD4 |
| 1866 | Caspase recruitment domain-containing protein 9 | CARD9 |
| 1867 | C-X-C motif chemokine 16 | CXCL16 |
| 1868 | STE20-like serine/threonine-protein kinase | SLK |
| 1869 | Probable serine carboxypeptidase CPVL | CPVL |
| 1870 | Charged multivesicular body protein 4b | CHMP4B |
| 1871 | Frizzled-8 | FZD8 |
| 1872 | Rab GTPase-binding effector protein 2 | RABEP2 |
| 1873 | CXADR-like membrane protein | CLMP |
| 1874 | Epidermal growth factor receptor kinase substrate 8-like protein 2 | EPS8L2 |
| 1875 | Anthrax toxin receptor 1 | ANTXR1 |
| 1876 | UPF0454 protein C12orf49 | C12orf49 |
| 1877 | Leucine-rich repeat-containing protein 19 | LRRC19 |
| 1878 | MANSC domain-containing protein 1 | MANSC1 |
| 1879 | Multimerin-2 | MMRN2 |
| 1880 | Chondrolectin | CHODL |
| 1881 | Sialate O-acetylesterase | SIAE |
| 1882 | Tumor necrosis factor receptor superfamily member 27 | EDA2R |
| 1883 | Retinoid-inducible serine carboxypeptidase | SCPEP1 |
| 1884 | Cadherin-related family member 5 | CDHR5 |
| 1885 | Collectrin | TMEM27 |
| 1886 | Putative sodium-coupled neutral amino acid transporter 10 | SLC38A10 |
| 1887 | Cadherin-20 | CDH20 |

**Supp. Table 5. All urinary proteins measured**

| Protein No. | Protein names | Gene names |
|---|---|---|
| 1888 | EGF, latrophilin and seven transmembrane domain-containing protein 1 | ELTD1 |
| 1889 | Otoferlin | OTOF |
| 1890 | Mucin-5B | MUC5B |
| 1891 | Copine-5 | CPNE5 |
| 1892 | UPF0606 protein KIAA1549 | KIAA1549 |
| 1893 | Platelet glycoprotein VI | GP6 |
| 1894 | Endosialin | CD248 |
| 1895 | Cadherin EGF LAG seven-pass G-type receptor 2 | CELSR2 |
| 1896 | Resistin | RETN |
| 1897 | Tumor necrosis factor receptor superfamily member 12A | TNFRSF12A |
| 1898 | CD320 antigen | CD320 |
| 1899 | Protocadherin-12 | PCDH12 |
| 1900 | Interleukin-1 receptor accessory protein | IL1RAP |
| 1901 | Complement component C1q receptor | CD93 |
| 1902 | Signal peptide, CUB and EGF-like domain-containing protein 2 | SCUBE2 |
| 1903 | Serine protease inhibitor Kazal-type 5 | SPINK5 |
| 1904 | Cartilage acidic protein 1 | CRTAC1 |
| 1905 | Omega-amidase NIT2 | NIT2 |
| 1906 | Nectin-3 | PVRL3 |
| 1907 | Baculoviral IAP repeat-containing protein 6 | BIRC6 |
| 1908 | Scavenger receptor cysteine-rich type 1 protein M160 | CD163L1 |
| 1909 | Matrix-remodeling-associated protein 5 | MXRA5 |
| 1910 | Cytokine-like protein 1 | CYTL1 |
| 1911 | 14 kDa phosphohistidine phosphatase | PHPT1 |
| 1912 | Tumor necrosis factor receptor superfamily member 19 | TNFRSF19 |
| 1913 | Gastrokine-1 | GKN1 |
| 1914 | EMILIN-3 | EMILIN3 |
| 1915 | Glycoprotein integral membrane protein 1 | GINM1 |
| 1916 | Transmembrane protein 106B | TMEM106B |
| 1917 | Protein FAM49B | FAM49B |
| 1918 | UPF0587 protein C1orf123 | C1orf123 |
| 1919 | Stabilin-1 | STAB1 |
| 1920 | N-acetyllactosaminide beta-1,3-N-acetylglucosaminyltransferase 2 | B3GNT2 |
| 1921 | Cadherin EGF LAG seven-pass G-type receptor 1 | CELSR1 |
| 1922 | Podocalyxin-like protein 2 | PODXL2 |
| 1923 | Alpha-hemoglobin-stabilizing protein | AHSP |
| 1924 | G-protein coupled receptor family C group 5 member B | GPRC5B |
| 1925 | Eukaryotic translation initiation factor 2-alpha kinase 3 | EIF2AK3 |
| 1926 | Methionine adenosyltransferase 2 subunit beta | MAT2B |

**Supp. Table 5. All urinary proteins measured**

| Protein No. | Protein names | Gene names |
|---|---|---|
| 1927 | Interleukin-1 receptor accessory protein-like 1 | IL1RAPL1 |
| 1928 | Complement C1r subcomponent-like protein | C1RL |
| 1929 | Calmodulin-like protein 5 | CALML5 |
| 1930 | Leucine-rich repeat transmembrane protein FLRT3 | FLRT3 |
| 1931 | Cysteine-rich motor neuron 1 protein | CRIM1 |
| 1932 | Charged multivesicular body protein 5 | CHMP5 |
| 1933 | Neurotrimin | NTM |
| 1934 | Costars family protein ABRACL | ABRACL |
| 1935 | Prostaglandin F2 receptor negative regulator | PTGFRN |
| 1936 | Small proline-rich protein 3 | SPRR3 |
| 1937 | Ammonium transporter Rh type C | RHCG |
| 1938 | C-type mannose receptor 2 | MRC2 |
| 1939 | Cornulin | CRNN |
| 1940 | Guanine nucleotide-binding protein G(I)/G(S)/G(O) subunit gamma-12 | GNG12 |
| 1941 | Glyoxylate reductase/hydroxypyruvate reductase | GRHPR |
| 1942 | Cathepsin Z | CTSZ |
| 1943 | Peflin | PEF1 |
| 1944 | Cathepsin F | CTSF |
| 1945 | Kallikrein-11 | KLK11 |
| 1946 | DnaJ homolog subfamily B member 4 | DNAJB4 |
| 1947 | Klotho | KL |
| 1948 | Myelin protein zero-like protein 1 | MPZL1 |
| 1949 | Neuroplastin | DKFZp566H1924 |
| 1950 | CGG triplet repeat-binding protein 1 | CGGBP1 |
| 1951 | Deleted in malignant brain tumors 1 protein | DMBT1 |
| 1952 | Fetuin-B | FETUB |
| 1953 | CMRF35-like molecule 8 | CD300A |
| 1954 | Sushi domain-containing protein 2 | SUSD2 |
| 1955 | Niemann-Pick C1-like protein 1 | NPC1L1 |
| 1956 | Tachykinin-3 | TAC3 |
| 1957 | ProSAAS | PCSK1N |
| 1958 | Prenylcysteine oxidase 1 | PCYOX1 |
| 1959 | A disintegrin and metalloproteinase with thrombospondin motifs 1 | ADAMTS1 |
| 1960 | Dipeptidyl peptidase 2 | DPP7 |
| 1961 | Brain-specific angiogenesis inhibitor 1-associated protein 2-like protein 1 | BAIAP2L1 |
| 1962 | Enolase-phosphatase E1 | ENOPH1 |
| 1963 | SLAM family member 5 | CD84 |
| 1964 | Leucyl-cystinyl aminopeptidase | LNPEP |
| 1965 | Serpin B13 | SERPINB13 |

**Supp. Table 5. All urinary proteins measured**

| Protein No. | Protein names | Gene names |
|---|---|---|
| 1966 | N-acetyl-D-glucosamine kinase | NAGK |
| 1967 | Annexin A10 | ANXA10 |
| 1968 | N-acetylglucosamine-1-phosphotransferase subunit gamma | GNPTG |
| 1969 | N-acetylglucosamine-1-phosphodiester alpha-N-acetylglucosaminidase | NAGPA |
| 1970 | Vacuolar protein sorting-associated protein 28 homolog | VPS28 |
| 1971 | Kallikrein-13 | KLK13 |
| 1972 | Protein kinase C and casein kinase substrate in neurons protein 3 | PACSIN3 |
| 1973 | Thyrotropin-releasing hormone-degrading ectoenzyme | TRHDE |
| 1974 | Angiopoietin-related protein 2 | ANGPTL2 |
| 1975 | Prion-like protein doppel | PRND |
| 1976 | Endomucin | EMCN |
| 1977 | Protein HEG homolog 1 | HEG1 |
| 1978 | RING finger protein 150 | RNF150 |
| 1979 | Unconventional myosin-Vb | MYO5B |
| 1980 | Frizzled-4 | FZD4 |
| 1981 | Coronin-1C | CORO1C |
| 1982 | Apoptosis-associated speck-like protein containing a CARD | PYCARD |
| 1983 | Mammalian ependymin-related protein 1 | EPDR1 |
| 1984 | Neurogenic locus notch homolog protein 3 | NOTCH3 |
| 1985 | Unconventional myosin-VI | MYO6 |
| 1986 | Lysosomal thioesterase PPT2 | PPT2 |
| 1987 | Neudesin | NENF |
| 1988 | Vacuolar protein sorting-associated protein 4A | VPS4A |
| 1989 | Protocadherin beta-8 | PCDHB8 |
| 1990 | Protocadherin gamma-C3 | PCDHGC3 |
| 1991 | Protocadherin alpha-6 | PCDHA6 |
| 1992 | Protein kinase C and casein kinase substrate in neurons protein 2 | PACSIN2 |
| 1993 | Endothelial protein C receptor | PROCR |
| 1994 | Multiple inositol polyphosphate phosphatase 1 | MINPP1 |
| 1995 | NSFL1 cofactor p47 | NSFL1C |
| 1996 | Frizzled-1 | FZD1 |
| 1997 | | |
| 1998 | V-set and immunoglobulin domain-containing protein 4 | VSIG4 |
| 1999 | Integral membrane protein 2B | ITM2B |
| 2000 | Band 4.1-like protein 3 | EPB41L3 |
| 2001 | Lambda-crystallin homolog | CRYL1 |
| 2002 | von Willebrand factor A domain-containing protein 7 | VWA7 |
| 2003 | Calcium-binding protein 39 | CAB39 |
| 2004 | Talin-1 | TLN1 |

**Supp. Table 5. All urinary proteins measured**

| Protein No. | Protein names | Gene names |
|---|---|---|
| 2005 | Neurexin-3 | NRXN3 |
| 2006 | Plexin-D1 | PLXND1 |
| 2007 | Low-density lipoprotein receptor-related protein 12 | LRP12 |
| 2008 | Angiopoietin-related protein 3 | ANGPTL3 |
| 2009 | Protocadherin gamma-C5 | PCDHGC5 |
| 2010 | Protocadherin alpha-3 | PCDHA3 |
| 2011 | Suppressor of tumorigenicity 14 protein | ST14 |
| 2012 | Lymphatic vessel endothelial hyaluronic acid receptor 1 | LYVE1 |
| 2013 | Heme-binding protein 2 | HEBP2 |
| 2014 | Phosphoserine aminotransferase | PSAT1 |
| 2015 | Junctional adhesion molecule A | F11R |
| 2016 | Carboxypeptidase Q | CPQ |
| 2017 | G-protein coupled receptor 56 | GPR56 |
| 2018 | Chloride intracellular channel protein 4 | CLIC4 |
| 2019 | EMILIN-1 | EMILIN1 |
| 2020 | Tumor necrosis factor receptor superfamily member 11A | TNFRSF11A |
| 2021 | Adseverin | SCIN |
| 2022 | Calpain-7 | CAPN7 |
| 2023 | Bis(5-adenosyl)-triphosphatase ENPP4 | ENPP4 |
| 2024 | Protein S100 | S100A6 |
| 2025 | DENN domain-containing protein 4C | DENND4C |
| 2026 | Fatty acid-binding protein, heart | FABP3 |
| 2027 | Lipolysis-stimulated lipoprotein receptor | LSR |
| 2028 | | AMBP |
| 2029 | Calpain small subunit 1 | CAPNS1 |
| 2030 | OX-2 membrane glycoprotein | CD200 |
| 2031 | Histone H2B | HIST1H2BN |
| 2032 | Low-density lipoprotein receptor-related protein 3 | LRP3 |
| 2033 | | APOC2 |
| 2034 | Apolipoprotein A-II | APOA2 |
| 2035 | Signaling lymphocytic activation molecule | SLAMF1 |
| 2036 | Carboxypeptidase | CTSA |
| 2037 | Carboxylic ester hydrolase | CEL |
| 2038 | Uromodulin | UMOD |
| 2039 | Complement C1q subcomponent subunit A | C1QA |

**Supp. Table 6. Molecular pathways**

| Pathway Number | Pathway Name |
|---|---|
| 1 | ESKD - Coagulation |
| 2 | ESKD - Acute Phase Response |
| 3 | ESKD - Intrinsic Prothrombin Activation |
| 4 | ESKD - GP6 |
| 5 | Control - Acute Phase Response Signaling |
| 6 | Control - Cardiac Hypertrophy Signaling |
| 7 | Control - Gα12/13 Signaling |
| 8 | Control - Glioma Signaling |
| 9 | Control - IL-15 Signaling |
| 10 | Control - LXR-RXR Activation |
| 11 | Control - NF-κB Signaling |
| 12 | Control - PTEN Signaling |
| 13 | Control - Regulation of the Epithelial-Mesenchymal Transition by Growth Factors Pathway |
| 14 | Control - Stat3 Pathway |
| 15 | Control - Tumor Microenvironment Pathway |
| 16 | Control T-Cell Exhaustion Signaling Pathway |
| 17 | Control - Complement System |
| 18 | Control - Neuroprotective Role of THOP1 in Alzheimer's Disease |
| 19 | Control - Coagulation System |
| 20 | Control - Intrinsic Prothrombin Activation Pathway |
| 21 | Control - Production of Nitric Oxide and Reactive Oxygen Species |
